# Diverging trends in health at older ages in England, 2004–2024: evidence from the English Longitudinal Study of Ageing

**DOI:** 10.64898/2026.07.13.26357914

**Authors:** Jiawei Wu, Karen Glaser, Debora Price, Giorgio Di Gessa

## Abstract

**Background:** Given uncertainty about whether later-life health at similar ages is improving over time, we examined trends across multiple health domains.

**Methods:** We analysed data from community-dwelling adults aged 50 and older in the English Longitudinal Study of Ageing in 2004/05, 2012/13, and 2023/24 (main survey: N=8389, 8549, and 6090, respectively). Outcomes included self-rated health, limiting long-standing illness, pain, mobility limitations, cardiometabolic and chronic conditions, obesity, inflammation, mental health, quality of life, and memory. Weighted pooled modified Poisson and linear regressions compared outcomes over time, overall, and by age group and education, with additional adjustment for sex and wealth.

**Results:** Adjusted estimates showed divergent trends. Fair/poor self-rated health increased from 27% to 34%, and any pain from 37% to 47%, whereas mobility impairments declined from 58% to 52%. Self-reported high cholesterol increased from 19% to 39%, while biomarker-defined high cholesterol declined from 78% to 54%; diabetes increased on both measures. Psychiatric problems increased from 6% to 10%, quality of life declined, and memory improved. However, trends differed by age and education, particularly for limiting long-standing illness, mobility limitations, cholesterol biomarkers, and mental health, indicating that aggregate trends masked unevenly distributed changes.

**Conclusion:** Later-life health in England has not improved uniformly. Gains in functioning, biomarkers, and cognition coexist with rising pain and poorer mental health. Trends were also socially and age patterned, producing increasingly multidimensional and socially patterned health outcomes. Multidomain health monitoring is essential for interpreting population health trends and planning healthy ageing, prevention, long-term care, and work policies.

**Key points:**

- Health at older ages in England changed in divergent rather than uniformly favourable directions between 2004/05 and 2023/24.
- Improvements in mobility, selected biomarkers and memory coexisted with rising pain, diabetes, poorer mental health and lower quality of life.
- Self-reported and biomarker-defined cardiometabolic trends diverged, highlighting the importance of how health is measured.
- Trends differed by age and education, showing that population averages can conceal persistent or worsening disadvantage.
- Monitoring and planning for ageing populations should use multidomain health measures rather than chronological age or single indicators alone.

## Introduction

Whether health at similar ages has improved or deteriorated over time remains an unresolved question in population health research [1]. This is an issue central to understanding the future implications of population ageing, since chronological age is often treated as a proxy for need, capacity, and risk across health, social care, and labour market policy. Policy responses will need to change if the health profile associated with a given age is itself changing over time. Understanding this question has become increasingly urgent as life expectancy gains have slowed in several high-income countries, including the UK, since the early 2010s, alongside declines in healthy life expectancy in the UK and some other countries [2], at a time when policymakers are pressing for extensions to working lives and tighter eligibility thresholds for state support [3].

A growing body of research has examined cohort and temporal changes in later-life health, but findings are mixed. Some studies have reported increases in prevalence of cardiometabolic risks (e.g., diabetes, hypertension) and diseases (e.g., stroke, heart disease) [1, 4–11], whereas others find stable or improving trends in indicators such as blood pressure and high cholesterol, heart disease and stroke [12–16], and in measures of functional limitations (e.g., activities of daily living (ADLs), instrumental activities of daily living (IADLs), and mobility limitations) [4, 9, 10, 17–20]. While memory has shown improvements over time [21–24], mental health [1, 4, 25–28] and quality of life [29] appear to have worsened among adults more recently entering mid-life.

Interpretation is further complicated because trends may differ according to whether health is measured through self-report, functioning, or biomarkers, and whether they refer to ever-or current health status. Improved diagnosis and treatment may increase reported prevalence while underlying physiological risk declines [4, 5]. Moreover, evidence on whether these contrasting trends are socially patterned remains limited [6, 17, 28]. For example, Choi et al. (2022) found declining disability among those aged 75+ in England and, to a lesser extent, the US between 2002 and 2016. Among those aged 55–75, however, trends were less favourable and increasingly socially patterned, contributing to widening health inequalities [17].

Most existing studies have focused on specific conditions or single health domains, used broad age groupings (e.g. working age, adults aged 65+), and paid limited attention to inequalities. Consequently, it remains unclear whether adults observed at similar ages in more recent periods have experienced consistent improvements in health, or whether gains in some domains coexist with deterioration in others. This uncertainty is compounded by the limited integration of self-reported and biomarker-based measures, which can show contrasting trends, and by insufficient evidence on whether changes in later-life health differ across sociodemographic groups. A multidomain assessment of health at comparable ages over time, within a consistent analytical framework, is therefore needed to characterise the changing profile of health at older ages and its inequalities.

Using two decades of nationally representative data from the English Longitudinal Study of Ageing (ELSA), we compared adults aged 50 years and older at similar ages across three time points (2004/05, 2012/13, and 2023/24). We examined trends across multiple domains of health, including self-rated health and pain; I/ADLs and mobility; cardiometabolic and chronic conditions; mental health and quality of life; and cognition. By further examining these trends within age (50–59, 60– 69, 70-79, and 80+) and educational groups, we assessed whether changes in health were consistent across domains and socially patterned, providing new evidence on the changing nature of health in later life in England. Understanding how different domains of health are changing at comparable ages is important for population health surveillance, because chronological age alone may no longer capture the diversity of need, capacity, and risk in ageing populations.

## Methods

### Study design and population

We used data from the English Longitudinal Study of Ageing (ELSA), an ongoing longitudinal survey representative of individuals aged 50 and older who live in private households in England [30]. ELSA has collected information biennially since 2002/03, with regular refreshment samples to maintain its representativeness of older adults [31], and nurse visits in selected waves to obtain blood samples and objective assessments of physical function (see eligibility criteria in Supplementary File 1, Table S1). All ELSA waves received ethical approval, and all participants gave informed consent. Data can be accessed through the UK Data Service (SN 5050). We analysed cross-sectional, non-proxy core members from Wave 2 (2004/05, N=8,389), Wave 6 (2012/13, N=8,549), and Wave 11 (2023/24, N=6,090). These waves were selected because they are approximately 10 years apart and include nurse-visit data, allowing comparison of self-reported and observer-measured health outcomes. Analytical samples varied by outcome, according to questionnaire module, nurse visit, and blood availability.

### Health outcomes

We examined outcomes across three broad domains. (I) <u>Self-rated health, pain, and functional limitations</u>: self-rated health (SRH, dichotomised as ‘fair or poor’ versus ‘better’ health); presence of limiting long-standing illness (LLI); being often troubled with pain and having moderate or severe intensity of pain; any difficulties with activities of daily living (ADLs, e.g., eating, bathing); any difficulty with instrumental activities of daily living (IADLs, e.g., preparing a hot meal, shopping for groceries); inability to perform mobility and motor coordination tasks (e.g., walking 100 yards, picking up a 5p coin from a flat surface). (II) <u>Cardiometabolic and chronic conditions, obesity and inflammation</u>: we considered the *current prevalence* of both self-reported and observer-measured outcomes. Specifically, participants were asked if they currently had hypertension, high cholesterol, diabetes, arthritis, heart disease, or lung disease that had been diagnosed by a doctor.

We also considered whether respondents reported having recently had a stroke. Moreover, we considered the corresponding observer-measured biomarkers: hypertension (if respondents’ valid mean systolic blood pressure ≥140 mmHg or their valid mean diastolic blood pressure ≥ 90 mmHg), high cholesterol (if the blood total cholesterol level ≥ 5 mmol/L), and diabetes (if glycated haemoglobin level ≥ 48 mmol/mol, which is equivalent to ≥ 6.5%) [32]. Additionally, we considered obesity (defined as body mass index (BMI) ≥ 30, with height and weight measured during the nurse visit) and elevated high-sensitivity C-reactive protein (CRP, ≥ 3 mg/L) [33]. (III) <u>Mental health and cognition</u>: self-reported doctor-diagnosed psychiatric problems; continuous and categorical (≥4 symptoms) depressive symptoms using the 8-item Centre for Epidemiologic Studies Depression (CES-D) Scale [34] which includes 8 binary questions, such as feeling sad or having restless sleep, in the week prior to the interview; Quality of life (QoL) was assessed using the 19-item Control, Autonomy, Self-realisation and Pleasure (CASP-19) scale [35], specifically designed to assess subjective QoL among older individuals (with scores ranging from 0 to 57); and memory scores were measured by summing the number of 10 words that respondents correctly recalled, both immediately and after a delay (totalling 20 points). Full question wording, harmonisation decisions, and biomarker thresholds are provided in Supplementary File 1, Table S2. Syntax for reproducing the variables is available in Supplementary File 2.

### Key demographic and socioeconomic variables and covariates

We examined health inequalities by age groups and education. Age was grouped as 50–59, 60–69, 70–79, or 80 years and older. Education was categorised into low (up to lower secondary), medium (upper secondary), and high (university or above) following the International Standard Classification of Education (http://www.uis.unesco.org/). Models adjusted for sex and wealth quintiles.

### Analytical strategy

First, we provided weighted cross-sectional descriptives at each wave for each outcome and variable of interest. Then, we fitted pooled modified Poisson or linear regression models (depending on outcome type), first overall and then including interaction terms between wave and age group or education. All analyses employed relevant cross-sectional weights and survey settings for the main interview, self-completion questionnaire, nurse visit, and blood sample, to account for the differential probability of participation in each. Robust standard errors accounted for repeated observations among participants contributing to more than one wave. Results are presented as marginal predicted probabilities or means over time, and by age group and education. Full model estimates are available in Supplementary File 1, Tables S3-S8. All analyses were conducted using Stata 19.

## Results

**Table 1** summarises the weighted sociodemographic characteristics and health characteristics of adults aged 50 years and older in ELSA waves 2 (2004/05), 6 (2012/13), and 11 (2023/24). The age and sex composition was similar across waves, while the share of the population with higher education rose from 11.4% in 2004/05 to 28.0% in 2023/24. Health indicators suggested divergent unadjusted trends across domains. LLI was broadly stable at 36%, 34%, and 35%, respectively, whereas any pain became more common, rising from 38% in 2004/05 to 45% in 2023/24. Cardiometabolic indicators showed marked divergence between self-reported and biomarker-based measures. For example, the prevalence of self-reported hypertension increased from 35% (2004/05) to 38% (2023/24), whereas biomarker-defined hypertension declined from 38% to 29%. Prevalence of psychiatric problems increased from 7% to 10% by 2023–24, whereas mean memory scores increased from 9.74 in 2004/05 to 10.83 in 2023/24.

**Table 1.** Weighted Proportion of ELSA Respondent Characteristics in Waves 2 (2004/05), 6 (2012/13), and 11 (2023/24)

|  | Wave 2 (2004/05) | Wave 6 (2012/13) | Wave 11 (2023/24) |
| --- | --- | --- | --- |
| Unweighted main survey N | 8,389 | 8,549 | 6,090 |
| Unweighted self-completion N | 7,545 | 7,721 | 5,630 |
| Unweighted nurse visit N | 7,401 | 7,519 | 4,827 |
| Unweighted blood sample N | 6,011 | 6,032 | 3,705 |
| ***Socio-demographics*** |  |  |  |
| Age group |  |  |  |
| 50-59 | 0.32 (0.31, 0.33) | 0.35 (0.34, 0.37) | 0.34 (0.33, 0.36) |
| 60-69 | 0.31 (0.30, 0.32) | 0.32 (0.31, 0.33) | 0.30 (0.28, 0.31) |
| 70-79 | 0.24 (0.23, 0.25) | 0.20 (0.19, 0.21) | 0.23 (0.22, 0.24) |
| 80+ | 0.13 (0.12, 0.14) | 0.12 (0.11, 0.13) | 0.13 (0.12, 0.14) |
| Women | 0.54 (0.53, 0.55) | 0.53 (0.52, 0.54) | 0.53 (0.52, 0.54) |
| Wealth quintile (selected) |  |  |  |
| 1st (poorest) | 0.20 (0.19, 0.21) | 0.21 (0.19, 0.21) | 0.20 (0.19, 0.22) |
| 5th (wealthiest) | 0.20 (0.19, 0.21) | 0.20 (0.19, 0.21) | 0.20 (0.19, 0.21) |
| Education |  |  |  |
| High | 0.11 (0.10, 0.12) | 0.17 (0.16, 0.18) | 0.28 (0.27, 0.30) |
| Medium | 0.47 (0.46, 0.48) | 0.57 (0.56, 0.58) | 0.58 (0.56, 0.59) |
| Low | 0.42 (0.41, 0.43) | 0.26 (0.25, 0.27) | 0.14 (0.13, 0.16) |
| ***Health outcomes panel A. Self-rated health, pain, and functional limitations*** |  |  |  |
| Poor/fair self-rated health | 0.29 (0.28, 0.30) | 0.27 (0.26, 0.29) | 0.31 (0.30, 0.33) |
| Limiting long-standing illness | 0.36 (0.35, 0.37) | 0.34 (0.33, 0.35) | 0.35 (0.34, 0.37) |
| Any pain | 0.38 (0.37, 0.39) | 0.42 (0.41, 0.43) | 0.45 (0.43, 0.47) |
| Moderate/severe pain | 0.28 (0.27, 0.29) | 0.29 (0.28, 0.30) | 0.30 (0.29, 0.32) |
| If reported ADL limitations | 0.21 (0.20, 0.22) | 0.17 (0.16, 0.18) | 0.18 (0.17, 0.20) |
| If reported IADL limitations | 0.23 (0.22, 0.24) | 0.19 (0.18, 0.20) | 0.19 (0.18, 0.21) |
| If reported mobility impairments | 0.59 (0.58, 0.61) | 0.52 (0.51, 0.53) | 0.50 (0.48, 0.52) |
| ***Health outcomes panel B. Cardiometabolic and chronic conditions, obesity, and inflammation*** |  |  |  |
| Self-reported hypertension | 0.35 (0.34, 0.36) | 0.38 (0.37, 0.39) | 0.38 (0.36, 0.39) |
| Hypertension biomarker | 0.38 (0.37, 0.40) | 0.31 (0.29, 0.32) | 0.29 (0.27, 0.31) |
| Self-reported high cholesterol | 0.19 (0.18, 0.20) | 0.33 (0.32, 0.34) | 0.36 (0.35, 0.38) |
| High cholesterol biomarker | 0.77 (0.76, 0.79) | 0.67 (0.66, 0.69) | 0.55 (0.53, 0.57) |
| Self-reported diabetes | 0.09 (0.08, 0.09) | 0.12 (0.11, 0.13) | 0.12 (0.11, 0.13) |
| Diabetes biomarker | 0.07 (0.06, 0.08) | 0.11 (0.10, 0.12) | 0.10 (0.08, 0.11) |
| Self-reported arthritis | 0.36 (0.35, 0.37) | 0.40 (0.39, 0.41) | 0.34 (0.33, 0.36) |
| Self-reported stroke | 0.02 (0.01, 0.02) | 0.02 (0.01, 0.02) | 0.01 (0.01, 0.02) |
| Self-reported heart diseases | 0.15 (0.14, 0.16) | 0.16 (0.15, 0.17) | 0.17 (0.15, 0.18) |
| Self-reported lung disease | 0.05 (0.05, 0.06) | 0.05 (0.05, 0.06) | 0.05 (0.05, 0.06) |
| Obesity | 0.29 (0.28, 0.30) | 0.32 (0.31, 0.34) | 0.33 (0.31, 0.35) |
| Elevated C-reactive protein | 0.37 (0.36, 0.38) | 0.29 (0.27, 0.30) | 0.24 (0.22, 0.26) |
| ***Health outcomes panel C. Mental health and cognition*** |  |  |  |
| Psychiatric problems | 0.07 (0.06, 0.07) | 0.07 (0.06, 0.08) | 0.10 (0.09, 0.11) |
| Depressed (CES-D-8 ≥4) | 0.16 (0.15, 0.17) | 0.15 (0.14, 0.16) | 0.17 (0.16, 0.18) |
| Mean depressive symptoms (0-8) | 1.61 (1.57, 1.65) | 1.45 (1.40, 1.51) | 1.63 (1.56, 1.70) |
| Mean CASP-19 score (0-57) | 40.86 (40.61, 41.10) | 39.45 (39.18, 39.72) | 39.55 (39.17, 39.93) |
| Mean memory score (0-20) | 9.74 (9.66, 9.82) | 10.60 (10.51, 10.69) | 10.83 (10.72, 10.95) |

Notes: Data sources: ELSA Waves 2, 6 and 11. Unless specified, we present proportions with 95% Confidence Intervals in brackets. Wave- and outcome-specific sampling (interview, self-completion, nurse, and venous) weights were applied. The syntax used to derive each outcome in each dataset (and a description of how outcomes were derived when discrepancies across waves were present) is available in Supplementary Files 1 and 2.

**Figure 1** shows adjusted predicted probabilities of self-rated health, pain, and functional limitations across survey waves, overall and by age group and education, with mixed trends (full model estimates for all Figures are in Supplementary File 1, Tables S3–S8). The adjusted predicted probability of fair/poor SRH increased from 27% to 34% (+7 percentage points [pp], 95% CI 5 to 9), LLI from 35% to 37% (+3 pp, 1 to 4), and moderate or severe pain from 27% to 33% (+6 pp, 4 to 8). Between W2 and W11, I/ADL limitations changed little, whereas mobility impairments declined from 58% to 52% (–6 pp, –8 to –4). Age-stratified analyses indicated significant heterogeneity for fair/poor SRH, LLI, and mobility impairments. Fair or poor SRH increased by 12 pp among those 80+ compared to 5 pp among adults aged 50–69; LLI changed little among 50-79 but increased from 46% to 55% among people aged 80 and older. Although pain increased across all age groups, trends did not differ significantly by age group, whereas mobility impairments declined among adults aged 50–79 years but not among those aged 80+. Heterogeneity of trends was observed for education. LLI and IADLs, for example, declined among respondents with higher education (–5 pp) but increased among those with lower education (+5 and +4 pp, respectively). By contrast, trends in SRH, any pain, and ADL limitations did not differ significantly by education.

**Figure 1.**
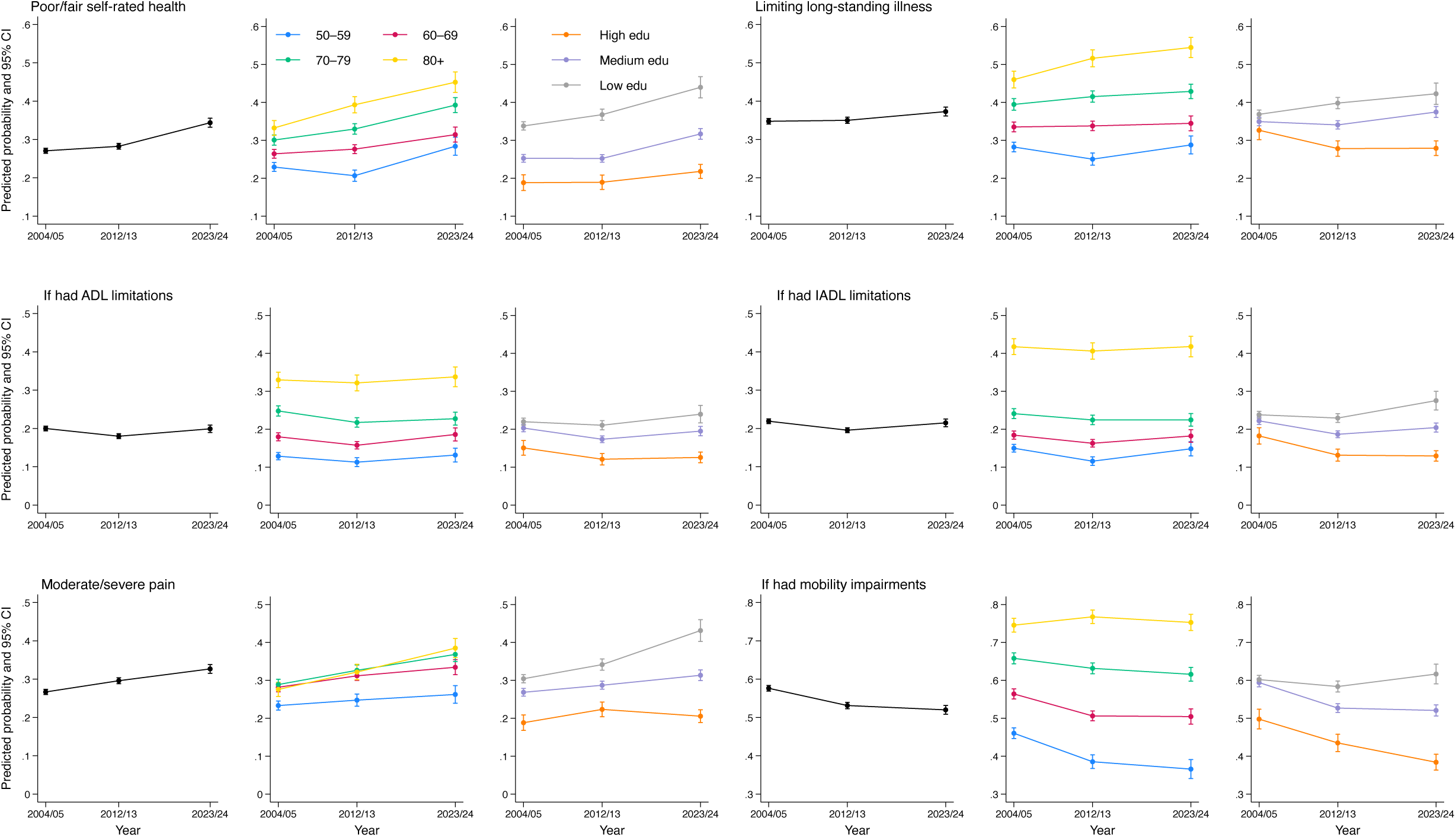
Marginal predicted probabilities of self-rated health, pain, and functional limitations across time points (overall, by age groups, and education) Source: English Longitudinal Study of Ageing (ELSA) Waves 2 (2004/05), 6 (2012/13), and 11 (2023/24). Notes: Weighted and adjusted models (adjusted for age group, sex, educational attainment, and wealth quintiles. Predicted probabilities are shown with 84% confidence intervals to facilitate visual comparisons; statistical inference was based on model contrasts and interaction tests (at the 5% level). Graphs across domains are not directly comparable because the y-axis values represent different prevalence levels.

**Figure 2** shows adjusted predicted probabilities of self-reported and biomarker-defined cardiometabolic conditions, obesity, and inflammation (overall, by age group, and by educational attainment). Overall, trends in self-reported and biomarker-defined measures diverged markedly for hypertension and high cholesterol. For instance, self-reported hypertension increased from 35% to 40% (+5 pp, 95% CI 3 to 7), whereas biomarker-defined hypertension declined from 38% to 30% (– 8 pp, –10 to –6). Prevalence of diabetes, however, increased in both self-reported (from 8% to 13%) and biomarker-defined (from 7% to 11%) measures. Obesity also increased, from 28% to 34% (+6 pp, 4 to 9), whereas elevated C-reactive protein declined from 37% to 24% (–12 pp, –15 to –10). Age-stratified analyses indicated significant heterogeneity for self-reported hypertension, self-reported and biomarker-defined high cholesterol, and elevated C-reactive protein, but not for biomarker-defined hypertension or either diabetes measure. For example, self-reported hypertension increased little among adults aged 50–59 but rose by 19 pp among those aged 80 and older, while biomarker-defined high cholesterol declined across all age groups, with larger reductions among adults aged 60 and older. Educational heterogeneity was evident for self-reported diabetes and biomarker-defined high cholesterol. Self-reported diabetes remained stable at 9% among highly educated respondents but increased from 9% to 15% (p<0.001) among low-educated adults. Biomarker-defined high cholesterol declined in all educational groups, with the largest reduction among those with low education (–32 pp vs –16 pp in the high-education group).

**Figure 2.**
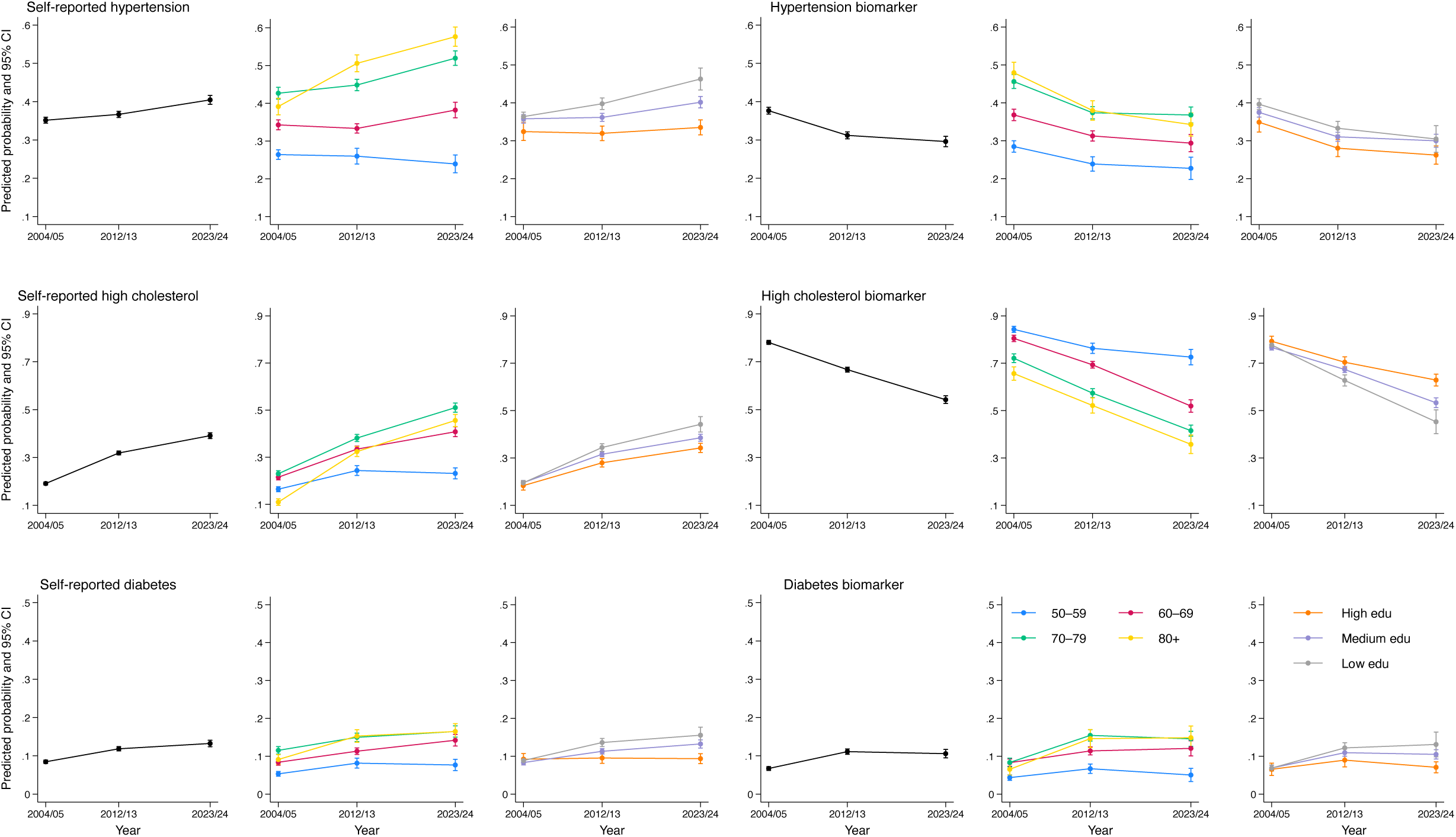
Marginal predicted probabilities of hypertension, high cholesterol, and diabetes across time points (overall, by age groups, and education) Source: English Longitudinal Study of Ageing (ELSA) Waves 2 (2004/05), 6 (2012/13), and 11 (2023/24). Notes: Weighted and adjusted models (adjusted for age group, sex, educational attainment, and wealth quintiles). Predicted probabilities are shown with 84% confidence intervals to facilitate visual comparisons; statistical inference was based on model contrasts and interaction tests (at the 5% level). Graphs across domains are not directly comparable because the y-axis values represent different prevalence levels.

**Figure 3** shows adjusted estimates for mental health, quality of life, and memory across ELSA waves 2, 6, and 11, overall and by age group and educational attainment. Overall, self-reported psychiatric problems increased from 6% to 10% (+4 pp, 95% CI 2 to 5); mean depressive symptom score increased by 0.13 (95% CI: 0.05 to 0.21, with scores going from 1.56 to 1.69); mean CASP-19 scores declined from 41.14 to 39.12 (–2.04, –2.49 to –1.58), whereas memory scores improved by +0.58 (95% CI: 0.44 to 0.72). Age-stratified analyses indicated significant heterogeneity for all health outcomes in this domain. Deterioration in depressive symptoms and quality of life was concentrated among adults aged 50–69, while the largest memory gains were observed among adults aged 70 and older. Finally, although depression, poorer quality of life, and lower memory were more common among adults with lower education, we found little evidence of differential trends by education.

**Figure 3.**
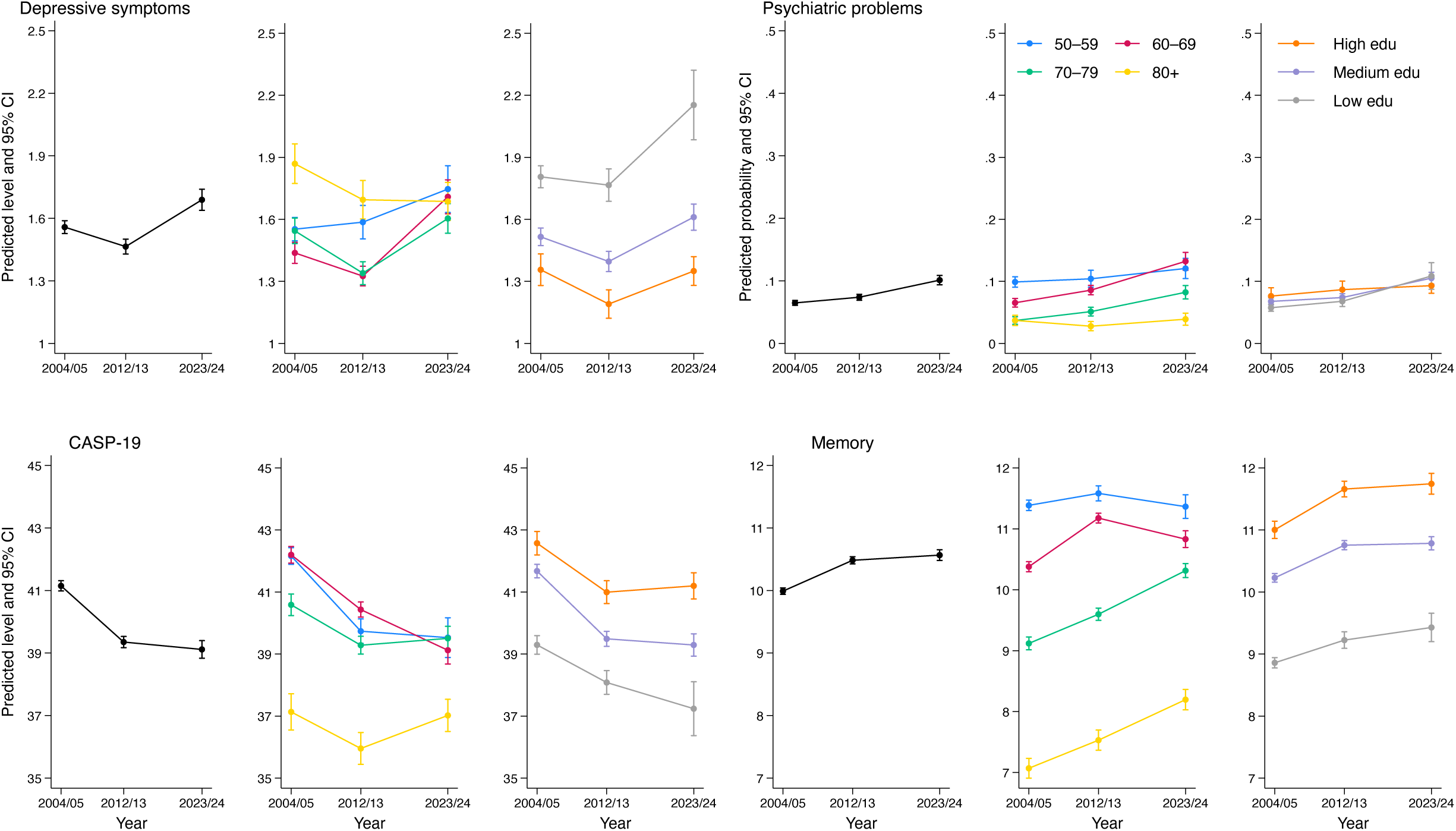
Marginal predicted probabilities and mean values of mental health and memory across time points (overall, by age groups, and education) Source: English Longitudinal Study of Ageing (ELSA) Waves 2 (2004/05), 6 (2012/13), and 11 (2023/24). Notes: Weighted and adjusted models (adjusted for age group, sex, educational attainment, and wealth quintiles). Predicted probabilities are shown with 84% confidence intervals to facilitate visual comparisons; statistical inference was based on model contrasts and interaction tests (at the 5% level). Graphs across domains are not directly comparable because the y-axis values represent different prevalence levels.

## Discussion

In this nationally representative study of adults aged 50 years and older in England, health at comparable ages changed in divergent rather than uniformly favourable directions between 2004/05 and 2023/24. A key strength is that contrasting trends were observed within a single national dataset, using comparable self-reported and biomarker measures of current, rather than ever-reported, health status. We found that trends in self-reported and biomarker-based measures of cardiometabolic health differed substantially, indicating that conclusions about population health change depend partly on how health is measured. We also found that favourable trends in IADLs and mobility limitations were concentrated among adults with higher educational attainment, whereas adults with lower educational attainment experienced worsening trends. Worsening of mental health and quality of life was most evident among adults aged 50-69. Taken together, these findings show that population averages or single health indicators alone can misrepresent changes in health at older ages, underscoring the need for multidomain approaches to population health surveillance in ageing societies.

We found that self-reported hypertension and high cholesterol increased over time, whereas biomarker-based measures declined. For example, in adjusted estimates, biomarker-defined high cholesterol prevalence fell from 78% to 54% while self-reports of high cholesterol doubled from 19% to 39%. This divergence was evident across age and education groups, particularly for high cholesterol. Improvements in biomarker measures were most pronounced among older and less educated groups. These patterns are consistent with improvements in diagnosis, monitoring, and treatment, increasing the proportion of adults aware of or treated for cardiometabolic conditions while reducing measured physiological risk [4, 5]. However, improvements in selected biomarkers should not be interpreted as evidence of uniformly improving cardiometabolic health. For example, prevalence of diabetes increased on both self-reported and biomarker-defined measures, from 8% to 13% and from 7% to 11%, respectively, indicating that favourable trends in some cardiometabolic indicators coexist with deterioration in others. These findings also show how reliance on self-reported conditions or biomarkers alone may lead to different conclusions about the direction and magnitude of population health change.

Functional and self-rated health trends were also socially patterned. LLI increased while mobility impairments declined overall, but improvements were concentrated among adults with higher educational attainment, whereas they worsened among those with lower educational attainment. For example, LLI declined among highly educated adults from 33% to 28% but increased among those with low education from 37% to 43%. Age differences were also evident: fair or poor SRH increased by 12 pp among adults aged 80 and older, compared with 5 pp among those aged 50–69, while mobility impairments improved among adults aged 50–79 but not among those aged 80 and older. Such evidence may suggest the presence of survival effects, reflecting improved survival following the onset of chronic disease(s) [36]. Previous evidence on changes in functional limitations had been mixed [4, 9, 10, 17], with some studies reporting improvements in population averages but less favourable trends among socioeconomically disadvantaged groups [17]. Our findings, therefore, suggest that aggregate improvements in selected functional outcomes can conceal persistent or widening inequalities among groups already experiencing poorer health.

Mental health, quality of life, and cognition also followed different patterns. Depressive symptoms and psychiatric problems increased, and quality of life declined, with less favourable changes particularly evident among adults aged 50–69 years. These patterns are consistent with previous British evidence of worsening psychological distress among adults entering midlife and older age [1, 4, 27]. By contrast, memory scores improved over time, especially among adults aged 70 years and older, consistent with studies reporting cognitive advantages in more recent populations reaching older ages [21, 22]. The coexistence of memory improvements alongside worsening mental health and quality of life reinforces the central interpretation of our findings: later-life health cannot be adequately characterised by a single indicator or by assuming uniform changes across population subgroups.

These findings have important implications for population health surveillance and policy. The health of adults in their 50s and 60s is central to policies intended to support longer working lives, yet this is also the age range in which less favourable trends in mental health and quality of life are most evident. Increasing pain, poorer mental well-being, and rising cardiometabolic risk factors may affect the ability of some adults to remain in work, particularly among socioeconomically disadvantaged groups. Although employment consequences were not assessed in this study, our findings might be relevant to understanding the recent rise in economic inactivity in the United Kingdom in this age group [37]. These patterns are also relevant for healthy ageing and long-term care policies, as gains in longevity may not translate into healthier lives [9]. More broadly, the diverging trends, combined with educational and age inequalities, indicate that monitoring health in ageing populations requires multidomain measures spanning physical functioning, cardiometabolic health, biomarkers, mental health, and cognition rather than relying on chronological age or any single health outcome.

Our study has several strengths. ELSA provides nationally representative data on adults aged 50 and older, collected with broadly comparable measures across three waves spanning two decades. The inclusion of self-reported and biomarker-based repeated measures of health allowed us to compare trends across multiple health domains and by age and education. Several limitations should also be noted. First, comparisons between adults observed at similar ages across survey periods cannot isolate birth-cohort effects from period effects, including the possible effects following the 2008/09 financial crisis and the COVID-19 pandemic. Second, ELSA does not represent adults living in residential or nursing care, among whom health limitations are likely to be greatest. Third, even when questions have not changed over time, trends in self-reported conditions might be affected by and reflect changes in diagnosis, treatment, awareness, expectations, or reporting. Finally, differential participation in nurse visits and blood sampling could affect biomarker comparisons despite the use of weights.

Overall, health at comparable ages in England has changed in divergent and socially patterned ways over the past two decades, suggesting that chronological age alone may be an increasingly incomplete proxy for need, capacity, and risk in later life. Improvements in some functional, cognitive, and biomarker measures coexist with worsening pain, mental well-being, and diabetes, with differential trends by age and education. Epidemiological monitoring of ageing populations should therefore use multidomain health profiles, rather than chronological age or single indicators alone, to inform healthy-ageing, prevention, work, and care policies.

## Statements and Declarations

### Author contributions

Giorgio Di Gessa had the initial idea for the paper; Karen Glaser and Debora Price further contributed to conceptualising the study. Giorgio Di Gessa designed the analysis and supervised its implementation. Jiawei Wu ran all the analyses. All authors contributed to reviewing and editing the manuscript. All authors read and approved the final manuscript. Giorgio Di Gessa is the guarantor and accepts full responsibility for the work and the conduct of the study.

### Ethics approval

All ELSA waves received ethical approval. Information on the ethical approval received for each wave of ELSA is available at https://www.elsa-project.ac.uk/ethical-approval. No additional ethical approval was required for this secondary analysis of de-identified data.

### Consent

All ELSA participants gave informed consent.

### Data Availability

All data can be accessed through the UK Data Service (SN 5050). More information on how to access data is available at https://www.elsa-project.ac.uk/accessing-elsa-data

### Code availability

Stata code to reproduce the main variables of interest used in this study is provided in Supplementary File 2.

## References

1. Gimeno, L., et al., The generational health drift: A systematic review of evidence from the British Birth Cohort Studies. Population Studies, 2026: p.1–19. DOI:10.1080/00324728.2026.2652038

2. Welsh, C.E., et al., Trends in life expectancy and healthy life years at birth and age 65 in the UK, 2008–2016, and other countries of the EU28: An observational cross-sectional study. The Lancet Regional Health–Europe, 2021. 2. DOI:10.1016/j.lanepe.2020.100023

3. Foster, L., Pensions and the extending working lives agenda in the UK: the impact on women. Journal of Population Ageing, 2023. 16(2): p.319–342. DOI: 10.1007/s12062-021-09354-2

4. Geiger, B.B., Has working-age morbidity been declining? Changes over time in survey measures of general health, chronic diseases, symptoms and biomarkers in England 1994–2014. BMJ open, 2020. 10(3): p.e032378. DOI: 10.1136/bmjopen-2019-032378

5. Jivraj, S., et al., Living longer but not necessarily healthier: The joint progress of health and mortality in the working-age population of England. Population Studies, 2020. 74(3): p.399–414. DOI: 10.1080/00324728.2020.1767297

6. Hudomiet, P., et al., Trends in health in midlife and late life. Journal of human capital, 2022. 16(1): p.133–156. DOI: 10.1086/717542

7. GBD 2021 Adult BMI Collaborators, Global, regional, and national prevalence of adult overweight and obesity, 1990–2021, with forecasts to 2050: a forecasting study for the Global Burden of Disease Study 2021. The Lancet, 2025. 405(10481): p.813–838. DOI: 10.1016/S0140-6736(25)00355-1

8. Koopman, C., et al., Trends in risk factors for coronary heart disease in the Netherlands. BMC Public Health, 2016. 16(1): p.835. DOI: 10.1186/s12889-016-3526-7

9. Gondek, D., et al., Post-war (1946-2017) population health change in the United Kingdom: A systematic review. PLOS ONE, 2019. 14(7): p.e0218991. DOI: 10.1371/journal.pone.0218991

10. Gimeno, L., et al., Cohort Differences in Physical Health and Disability in the United States and Europe. The Journals of Gerontology: Series B, 2024. 79(8). DOI: 10.1093/geronb/gbae113

11. Ong, K.L., et al., Global, regional, and national burden of diabetes from 1990 to 2021, with projections of prevalence to 2050: a systematic analysis for the Global Burden of Disease Study 2021. The Lancet, 2023. 402(10397): p.203–234. DOI: 10.1016/S0140-6736(23)01301-6

12. Guan, C., et al., Global, regional, and national burden of ischaemic heart disease and its trends, 1990–2019. Public health, 2023. 223: p.57–66. DOI: 10.1016/j.puhe.2023.07.010

13. Liu, C., et al., Global, regional, and national burden of ischaemic heart disease from 1990 to 2021: a comprehensive analysis based on the Global Burden of Disease study 2021. Journal of Global Health 2025. 15: p.04291. DOI:10.7189/jogh.15.04291

14. NCD Risk Factor Collaboration (NCD-RisC), Contributions of mean and shape of blood pressure distribution to worldwide trends and variations in raised blood pressure: a pooled analysis of 1018 population-based measurement studies with 88.6 million participants. International journal of epidemiology, 2018. 47(3): p.872–883i. DOI: 10.1093/ije/dyy016

15. NCD Risk Factor Collaboration (NCD-RisC), National trends in total cholesterol obscure heterogeneous changes in HDL and non-HDL cholesterol and total-to-HDL cholesterol ratio: a pooled analysis of 458 population-based studies in Asian and Western countries. International Journal of Epidemiology, 2020. 49(1): p.173–192. DOI: 10.1093/ije/dyz099

16. Beltrán-Sánchez, H., et al., Assessing morbidity compression in two cohorts from the Health and Retirement Study. J Epidemiol Community Health, 2016. 70(10): p.1011–1016. DOI: 10.1136/jech-2015-206722

17. Choi, H., et al., Differential Trends in Disability Among Rich and Poor Adults in the United States and England From 2002 to 2016. Journals of Gerontology Series B-Psychological Sciences & Social Sciences, 2022. 77(Suppl_2): p.S189–S198. DOI: 10.1093/geronb/gbac029

18. van Oostrom, S.H., et al., Time trends in prevalence of chronic diseases and multimorbidity not only due to aging: data from general practices and health surveys. PloS one, 2016. 11(8): p.e0160264. DOI: 10.1371/journal.pone.0160264

19. Fors, S., et al., Cohort-specific disability trajectories among older women and men in Europe 2004–2017. European Journal of Ageing, 2022. 19(4): p.1111–1119. DOI: 10.1007/s10433-022-00684-4

20. Zimmer, Z., et al., Persistent, consistent, and extensive: The trend of increasing pain prevalence in older Americans. The Journals of Gerontology: Series B, 2020. 75(2): p.436–447. DOI: 10.1093/geronb/gbx162

21. Brailean, A., et al., Cohort Differences in Cognitive Aging in the Longitudinal Aging Study Amsterdam. Journals of Gerontology Series B-Psychological Sciences & Social Sciences, 2018. 73(7): p.1214–1223. DOI: 10.1093/geronb/gbw129

22. Jagger, C., et al., A comparison of health expectancies over two decades in England: results of the Cognitive Function and Ageing Study I and II. The Lancet, 2016. 387(10020): p.779–786. DOI: 10.1016/S0140-6736(15)00947-2

23. Dodge, H.H., et al., Cohort effects in verbal memory function and practice effects: a population-based study. International Psychogeriatrics, 2017. 29(1): p.137–148. DOI: 10.1017/S1041610216001551

24. Thorvaldsson, V., et al., Better Cognition in New Birth Cohorts of 70 Year Olds, But Greater Decline Thereafter. Journals of Gerontology Series B-Psychological Sciences & Social Sciences, 2017. 72(1): p.16–24. DOI: 10.1093/geronb/gbw125

25. GBD Mental Disorders Collaborators, Global, regional, and national burden of 12 mental disorders in 204 countries and territories, 1990–2019: a systematic analysis for the Global Burden of Disease Study 2019. The Lancet Psychiatry, 2022. 9(2): p.137–150. DOI: 10.1016/S2215-0366(21)00395-3

26. Zhang, Y., et al., Depression trajectories from mid to late life (50-89 Years): The roles of cohort, multimorbidity status, and national contexts across nine European countries. Social Science & Medicine, 2025. 385: p.118605. DOI: 10.1016/j.socscimed.2025.118605

27. Gondek, D., et al., Psychological distress from early adulthood to early old age: evidence from the 1946, 1958 and 1970 British birth cohorts. Psychological Medicine, 2022. 52(8): p.1471–1480. DOI: 10.1017/S003329172000327X

28. Abrams, L.R., et al., Changes in depressive symptoms over age among older Americans: Differences by gender, race/ethnicity, education, and birth cohort. SSM - Population Health, 2019. 7: p.100399. DOI:10.1016/j.ssmph.2019.100399

29. Clause-Verdreau, A.C., et al., Contrasted trends in health-related quality of life across gender, age categories and work status in France, 1995-2016: repeated population-based cross-sectional surveys using the SF-36. Journal of Epidemiology & Community Health, 2019. 73(1): p.65–72. DOI: 10.1136/jech-2018-210941

30. Banks, J., et al., English Longitudinal Study of Ageing: Waves 0-11, 1998-2024. [data collection]. 50th Edition. UK Data Service. SN: 5050 doi: 10.5255/UKDA-SN-5050-37. 2025 DOI: 10.5255/UKDA-SN-5050-37

31. Steptoe, A., et al., Cohort Profile: The English Longitudinal Study of Ageing. International Journal of Epidemiology, 2012. 42(6): p.1640–1648. DOI:10.1093/ije/dys168

32. de Oliveira, C., et al., Mortality risk attributable to smoking, hypertension and diabetes among English and Brazilian older adults (The ELSA and Bambui cohort ageing studies). The European Journal of Public Health, 2016. 26(5): p.831–835. DOI:10.1093/eurpub/ckv225

33. Pearson, T.A., et al., Markers of inflammation and cardiovascular disease: application to clinical and public health practice: a statement for healthcare professionals from the Centers for Disease Control and Prevention and the American Heart Association. Circulation, 2003. 107(3): p.499–511. DOI:10.1161/01.cir.0000052939.59093.45

34. Radloff, L.S., The CES-D scale: A self-report depression scale for research in the general population. Applied Psych Measurement, 1977. 1(3): p.385–401. DOI:10.1177/014662167700100306

35. Hyde, M., et al., A measure of quality of life in early old age: the theory, development and properties of a needs satisfaction model (CASP-19). Aging Ment Health, 2003. 7(3): p.186–94. DOI:10.1080/1360786031000101157

36. Callaway, J., et al., Ageing populations: new challenges in longevity. BMC public health, 2025. 25(1): p.4395. DOI:10.1186/s12889-025-25531-w

37. Gimeno, L., et al., Chronic health conditions and health-related economic inactivity in midlife: Evidence from the 1958 and 1970 British birth cohorts. SSM - Population Health, 2026. 35. DOI:10.1016/j.ssmph.2026.101940

