## Supplementary File 1 for "Diverging trends in health at older ages in England, 2004–2024: evidence from the English Longitudinal Study of Ageing"

### Supplementary materials for manuscript "Diverging trends in health at older ages in England, 2004–2024: evidence from the English Longitudinal Study of Ageing "

### Table S1. Eligibility Criteria for nurse visit and blood sample collection.

<table>
<tr><td>Nurse Visit</td><td>Full eligibility criteria:<br><ul><li>Only core members who completed a main interview in person and were marked as eligible for a nurse visit at each wave were offered a nurse visit at the end of their interview.</li><li>No ELSA partners were eligible for nurse visits.</li><li>However, a small number of partners and non-eligible core members were given a nurse visit if someone else in their household was completing a nurse interview, they specifically requested it, and it was believed it would assist with their future participation in the survey.</li><li>Individuals who completed an interview by proxy were not eligible for a nurse visit.</li><li>There were specific eligibility criteria for each measure conducted by the nurse. These are briefly outlined below and in more detail in the ELSA Nurse User Guide (available on the UK Data Service website).<ol><li>BMI was calculated for all those respondents for whom both a valid height and weight measurement were recorded</li><li>All respondents were eligible for the blood pressure module, except those who were pregnant. The respondent was asked not to eat, smoke, drink alcohol or take vigorous exercise in the 30 minutes preceding the blood pressure measurement</li></ol></li></ul></td></tr>
<tr><td>Blood Sample</td><td>Full eligibility criteria:<br><ul><li>Blood samples were taken from willing ELSA core members</li><li>Exceptions were respondents who<ol><li>had a clotting or bleeding disorder (e.g. haemophilia and low platelets)</li><li>had ever had a fit</li><li>were not willing to give their consent in writing</li><li>were currently on anticoagulant drugs (e.g. warfarin therapy)</li></ol></li><li>Fasting blood samples were taken whenever possible, but respondents over 80 years, those known to be diabetic and on treatment, those with a clotting or bleeding disorder or on anticoagulant drugs (e.g., warfarin), those who had ever had fits, and those who seemed frail were not asked to fast. Subjects were considered to have fasted if they had not had food or drink, except water, for at least 5 hours prior to the blood test.</li></ul>All the blood samples were analysed at the Royal Victoria Infirmary laboratory in Newcastle.</td></tr>
</table>

#### Table S2. Harmonisation of health outcomes in ELSA waves 2 (2004/05), 6 (2012/13), and 11 (2023/24)

| Outcome | Wave | (variable name) Question | Answer | Harmonisation |
|---|---|---|---|---|
| Self-rated health | 2, 6, 11 | (hehelf) Would you say your health is... | 1.Excellent<br>2.Very Good<br>3.Good<br>4.Fair<br>5.Poor | 1-3:At least good;<br>4/5:Fair/Poor |
| Limiting long-standing illness | 2, 6, 11 | (heill) Do you have any long-standing illness, disability or infirmity?<br>(helim) (Does this/Do these) illness(es) or disability(ies) limit your activities in any way? | 1.Yes<br>2.No | heill=2:No illness;<br>heill=1&helim=1:Illness not limiting;<br>heill=1&helim=2: Limiting illness |
| Any pain | 2, 6, 11 | (hepain) Are you often troubled with pain? | 1.Yes<br>2.No | 1.Yes<br>2.No |
| Moderate or severe pain | 2, 6, 11 | (hepaa) How bad is the pain most of the time? Is it… | 1.Mild<br>2.Moderate<br>3.Severe | hepain=1&hepaa=2/3: Severe/moderate pain;<br>hepain=0 or (hepain=1 & hepaa=1): no pain or mild pain |
| ADL limitations | 2, 6, 11 | (headlb) Because of a health or memory problem, do you have difficulty doing any of the activities? | 1.Dressing<br>2.Walking across a room<br>3.Bathing<br>4.Eating<br>5.Getting in/out of bed<br>6.Using the toilet | If the respondent reported 1 or more ADL limitations, they were classified as having ADL limitations |
| IADL limitations | 2, 6, 11 | (headlb) Because of a health or memory problem, do you have difficulty doing any of the activities? | 7.Using a map<br>8.Preparing meals<br>9.Shopping<br>10.Making phone calls<br>11.Taking medications<br>12.Doing work around the house/garden<br>13.Managing money | If the respondent reported 1 or more ADL limitations, they were classified as having IADL limitations |
| Mobility impairments | 2, 6, 11 | (headla) Do you have any difficulty doing each of these everyday activities? | 1.Walking 100 yards<br>2. Sitting for about 2hs<br>3.Getting up from a chair after sitting for long periods<br>4.Climbing several flights of stairs without resting<br>5.Climbing one flight of stairs without resting<br>6.Stooping, kneeling, or crouching<br>7.Reaching or extending your arms above shoulder level<br>8.Pulling or pushing large objects like a living room chair<br>9.Lifting or carrying weights over 10 pounds like a heavy bag of groceries<br>10.Picking up a 5p coin from a table | If the respondent reported 1 or more mobility impairments = yes |
| Self-reported lung disease | 2 | (hediad1) Whether confirms diagnosis of lung diseases recorded in wave 1?<br>(hediam1) Reason disputed chronic lung disease diagnosis from wave 1.<br>(hedids1) Do you still have lung diseases?<br>(hedib01-hedib04) Has a doctor ever told you that you have the following conditions (1st to 4th mention)? | (hediad1) 1.Yes 2.No -1.Not Applicable<br><br>(hediam1) 1.Never had 2.No longer has 3.Did not have previously, but has now -1.N.A. -8.Don't know<br>(hedids1) 1.Yes 2.No<br>(hedib01-hedib04) 1.Chronic lung disease | Respondents were considered as currently having lung disease if they (a) reported lung disease in wave 1, did not dispute this record in wave 2, and reported still having it in wave 2 [hediad1=1 & hedids1=1], or (2) did not have it previously but reported it in wave 2 [hediad1 !=1 & hediam1=3], or (3) did not report the condition in wave 1 but reported doctor diagnosis of chronic lung disease in wave 2 [hediad1 = -1 & (hedib01=1 \| hedib02=1 \| hedib03=1 \| hedib04=1)] |

| Outcome | Wave | (variable name) Question | Answer | Harmonisation |
|---|---|---|---|---|
| | | | | *Only 1 respondent had the issue of feedforward error on lung disease in wave 2 so we ignored it |
| | 6 | (hedbdlu) Whether confirms diagnosis of lung diseases recorded in previous wave(s)?<br>(hedbmlu) Reason disputed lung disease diagnosis fed forward.<br>(hedblu) Do you still have lung diseases?<br>(hediblu) Has a doctor ever told you that you have lung diseases? | (hedbdlu) 1.Yes 2.No -1.Not Applicable<br>(hedbmlu) 1.Never diagnosed 2.No longer has 3.Did not have previously but has now 4.Misdiagnosed -1. Item not applicable<br>(hedblu) 1.Yes 2.No<br>(hediblu) 1.Yes 2.No | Respondents were considered as currently having lung disease if they (a) reported lung disease in a prior wave, did not dispute this record in wave 6, and reported still having it in wave 6 [hedbdlu=1 & hedblu=1], or (2) did not report it previously but reported it in wave 6 [hedbmlu=3], or (3) reported doctor diagnosis of lung disease since last interview [hediblu=1]<br>* In wave 6, the prevalence of lung disease could only be calculated among Rs who were not in the refreshment sample (see Stata .do file for additional details). |
| | 11 | (hehavecl) Do you still have lung diseases? | (hehavecl) 1.Still have chronic lung disease but do not take medication or treatment for it. 2.Still have high chronic lung disease and take medication or treatment for it 3.No longer have chronic lung disease -1.Not applicable | Respondents were considered as currently having lung disease if hehavecl=(1 or 2) |
| Self-reported current hypertension | 2 | (hediac1) Whether confirms diagnosis of high blood pressure (BP) recorded in wave 1?<br>(hedian1) Reason R disputed high blood pressure diagnosis from wave 1<br>(hedias1) Do you still have high blood pressure?<br>(hedim01-08) Has a doctor ever told you that you have the following cardiovascular conditions (1st to 8th mention, merged variable)? | (hediac1) 1.Yes 2.No -1.Not applicable<br>(hedian1) 1. Never had 2. No longer has 3. Did not have previously but has now -1. Not applicable<br>(hedias1) 1.Yes 2.No<br>(hedim01-08) 1.High blood pressure | Respondents were considered as currently having hypertension if they (1) reported hypertension in wave 1, did not dispute this record in wave 2, and reported still having it in wave 2 [hediac1=1 & hedias1=1], or (2) did not have it previously but reported it in wave 2 [hediac1!=1 & hedian1=3], or (3) did not report the condition in wave 1 but reported doctor diagnosis of hypertension in wave 2 [hediac1= -1 & (hedim01 to hedim08=1)].<br>* Respondents with an issue with feedforward on this condition (n = 1,786) were randomly assigned based on observed prevalence (see Stata .do file for additional details) |
| | 6 | (hedacbp) Whether confirms diagnosis of high BP recorded previous wave(s)?<br>(hedanbp) Reason R disputed high blood pressure diagnosis fed forward<br>(hedasbp) Do you still have high BP?<br>(hedimbp) Newly diagnosed high blood pressure? | (hedacbp) 1.Yes 2.No -1.Not applicable<br>(hedanbp) 1.Never had 2.No longer has 3.Did not have previously but has now 4.Misdiagnosed -1.Not applicable -8. Don't know<br>(hedasbp) 1.Yes 2.No<br>(hedimbp) 1.Yes 2.No | Respondents were considered as currently having hypertension if they (1) reported hypertension in a prior wave, did not dispute this record in wave 6, and reported still having it in wave 6 [hedacbp=1 & hedasbp=1], or (2) did not report it previously but reported it in wave 6 [hedanbp=3], or (3) reported doctor diagnosis of hypertension since last interview [hedimbp=1]<br>* In wave 6, the prevalence of hypertension could only be calculated among Rs who were not in the refreshment sample (see Stata .do file for additional details). |
| | 11 | (hehavebp) Do you still have high BP? | (hehavebp) 1.Still have high blood pressure but do not take medication or treatment for it 2.Still have high blood pressure and take medication or treatment for it 3.No longer have high blood pressure -1. Not applicable -8. Don't know | Respondents were considered as currently having hypertension if hehavebp=(1 or 2) |
| Hypertension biomarker | 2, 6, 11 | (sysval) (Derived) valid mean systolic blood pressure | Valid values.<br>-1 = invalid or incomplete set of blood pressure readings | Hypertension if<br>140 <= sysval<= 300 or 90 <= diaval <= 150 |

| Outcome | Wave | (variable name) Question | Answer | Harmonisation |
|---|---|---|---|---|
| | | (diaval) (Derived) valid mean diastolic blood pressure | | |
| Self-reported current diabetes | 2 | (hediac7) Whether confirms diagnosis of diabetes recorded in wave 1?<br>(hedian7) Reason disputed diabetes or high blood sugar diagnosis fed forward<br>(hedias7) Do you still have diabetes?<br>(hedia0*) Has a doctor ever told you that you have diabetes?<br>(hedim01-08) Has a doctor ever told you that you have the following cardiovascular conditions (1st to 8th mention, merged variable)?<br>(heacd) Has a doctor ever told you that you have diabetes? | (hediac7) 1.Yes 2.No -1.Not Applicable<br>(hedian7) 1.Never had 2. No longer has 3. Did not have previously but has now -1. Not applicable<br>(hedias7) 1.Yes 2.No<br>(hedia0) 1.Yes 2.No<br>(hedim01-08) 7.Diabetes<br>(heacd) 1.Yes 2.No -1.Not Applicable | Respondents were considered as currently having diabetes if they (1) reported diabetes in wave 1, did not dispute this record in wave 2, and reported still having it in wave 2 [hediac7=1 & hedias7=1], or (2) did not have previously but reported it in wave 2 [hediac7!=1 & hedian7=3], or (3) did not report diabetes in wave 1 but reported doctor diagnosis of diabetes in wave 2 [hediac7= -1 & (hedim01 to hedim08=7)].<br>* Respondents with an issue with feedforward on this condition (n = 355) were randomly assigned based on observed prevalence (see Stata .do file for additional details). |
| | 6 | (hedacdi) Whether confirms diagnosis of diabetes recorded in previous wave(s)?<br>(hedandi) Reason R disputed diabetes diagnosis fed forward<br>(hedimdi) Newly diagnosed diabetes | (hedacdi) 1.Yes 2.No -1.Not Applicable<br>(hedandi) 1. Never had 2. No longer has 3. Did not have previously but has now 4. Misdiagnosed -1. Not applicable -8. Don't know<br>(hedimdi) 1.Yes 2.No | Respondents were considered as currently having diabetes if they (1) reported not having diabetes previously but have it in wave6 [hedandi=3], or (2) reported doctor diagnosis of diabetes since last interview [hedimdi=1].<br>* In wave 6, the prevalence of hypertension could only be calculated among Rs who were not in the refreshment sample (see Stata .do file for additional details). |
| | 11 | (hehavedi) Do you still have diabetes? | (hehavedi) 1.Still have diabetes but do not take medication or treatment for it 2.Still have diabetes and take medication or treatment for it 3.No longer have diabetes -1.N.A. -8.Don't know | Respondents were considered as currently having diabetes if hehavedi= (1 or 2) |
| Diabetes biomarker | 2, 6, 11 | (hba1c) Blood glycated haemoglobin level | Only valid values were considered<br>[(% in w2, mmol/mol in w6 and w11)] | Diabetes if<br>6.5 <= hba1c <= 120 [in wave 2, unit is %]<br>48 <= hba1c <= 130 [waves 6 and 11, unit is mmol/mol] |
| Self-reported current high total cholesterol | 2 | (hedim01-08) Has a doctor ever told you that you have the following cardiovascular conditions (1st to 8th mention, merged variable)? | (hedim01-08) 9.High cholesterol | Respondents were considered to currently have high total cholesterol if they reported ever being diagnosed by a doctor with high cholesterol (hedim01 to hedim08=9). In wave 2, Rs were not asked to confirm previous diagnosis or report if still had it. |
| | 6 | (hedacch) Whether confirms diagnosis of HC recorded in previous wave(s)?<br>(hedanch) Reason R disputed HC diagnosis fed forward<br>(hedasch) Do you still have abnormal HC?<br>(hedimch) Newly diagnosed high cholesterol? | (hedacch) 1.Yes 2.No -1.Not Applicable<br>(hedanch) 1. Never had 2. No longer has 3. Did not have previously but has it now 4. Misdiagnosed -1. Not applicable<br>(hedasch) 1.Yes 2.No<br>(hedimch) 1.Yes 2.No | Respondents were considered to currently have high total cholesterol if they (1) confirmed previous doctor diagnosis of high cholesterol and still had high cholesterol [hedacch=1 & hedasch=1], or (2) did not report it previously but reported it in wave 6 [hedanch=3], or (3) reported doctor diagnosis of hypertension since last interview [hedimch=1].<br>*In wave 6, the prevalence of hypertension could only be calculated among Rs who were not in the refreshment sample (see Stata .do file for additional details). |
| | 11 | (hehavehc) Do you still have high cholesterol? | (hehavehc) 1.Still have high cholesterol but do not take medication or treatment for it 2.Still have high cholesterol and take medication or treatment | Respondents were considered as currently having high total cholesterol if hehavehc=(1 or 2) |

| Outcome | Wave | (variable name) Question | Answer | Harmonisation |
|---|---|---|---|---|
| | | | for it 3.No longer have high cholesterol -1.Not applicable -8. Don't know | |
| High total cholesterol biomarker | 2, 6, 11 | (chol) Blood total cholesterol level (mmol/L) | Valid values.<br>-11:blood sample not taken; -8: cannot measure parameter reliably as TRIG value > 13mmol/L; -7:sample unusable for other reason; -6: sample took more than 5 days to reach the lab; -3: sample received but was haemolysed so not suitable for analysis; -2: sample received but insufficient blood for analysis; -1: sample not received | High total cholesterol if<br>chol >= 5 mmol/L |
| Self-reported current stroke | 2 | (hediac8) Whether confirms diagnosis of stroke recorded in wave 1?<br>(hedian8) Reason R disputed stroke diagnosis from wave 1<br>(hedim01-08) Has a doctor ever told you that you have the following cardiovascular conditions (1st to 8th mention, merged variable)?<br>(henmst) Number of strokes since last nurse visit? | (hediac8) 1.Yes 2.No -1.Not Applicable<br>(hedian8) 1. Never had 2. No longer has 3. Did not have previously but has it now -1. Not applicable<br>(hedim01-08) 8.A stroke<br>(henmst) Valid values | Respondents were considered as having recently had a stroke if they (1) did not report stroke in wave 1 but reported doctor diagnosis of stroke in wave 2 [hediac8!=1 & hedian8=3], or (2) did not have previously but reported it in wave 2 [hediac8 = -1 & (hedim01 to hedim08=8)], or (3) had >=1 stroke since last visit [henmst>=1]<br>* Respondents with an issue with feedforward on this condition (n = 183) could not be randomly assigned because there is no question "Do you still have stroke?" to calculate the prevalence for those not subject to this issue. |
| | 6 | (hedacst) Whether confirms diagnosis of stroke recorded in previous wave(s)?<br>(hedanst) Reason R disputed stroke diagnosis fedforward<br>(hediast) Has a doctor ever told you that you have strokes?<br>(hedimst) Newly diagnosed stroke?<br>(henmst) Number of strokes in the past 2 years | (hedacst) 1.Yes 2.No -1.Not Applicable<br>(hedanst) 1. Never had 2.No longer has 3.Did not have previously but has it now 4.Misdiagnosed -1.Not applicable<br>(hediast) 1.Yes 2.No<br>(hedimst) 1.Yes 2.No -9.Refusal -8.Don't know<br>Valid values | Respondents were considered as having recently had a stroke if they (1) did not report it previously but have it in wave 6 [hedanst=3], or (2) report doctor diagnosis of stroke since last interview [hedimst=1], or (3) has had >=1 stroke in the past two years [henmst>=1].<br>*In wave 6, the prevalence of hypertension could only be calculated among Rs who were not in the refreshment sample (see Stata .do file for additional details). |
| | 11 | (hediacst) Whether confirms previous diagnosis of stroke?<br>(hediia7) [Since we last saw you] has a doctor or other health professional [ever] told you that you have stroke?<br>(heothst) Since the last stroke you told us about, has a doctor or other health professional told you that you have had another stroke? | (hediacst) 1.Yes 2.No -1.Not Applicable<br>(hediia7) 1.Yes 2.No<br>(heothst) 1.Yes 2.No | Respondents were considered as having recently had a stroke if they (1) confirm previous diagnosis of stroke and has had another stroke since last diagnosis [hediacst=1 & heothst=1], or (2) did not report diagnosis of stroke previously but reports diagnosis of stroke in wave 11 [hediacst=-1 & hediia7=1]. |
| Self-reported current heart failure | 2 | (hediac4) Whether confirms diagnosis of heart failure recorded in wave 1?<br>(hedian4) Reason disputed diagnosis of heart failure from wave 1<br>(hedias4) Do you still have heart failure?<br>(hedim01-08) Has a doctor ever told you that you have the following cardiovascular conditions (1st to 8th mention, merged variable)? | (hediac4) 1.Yes 2.No -1.Not applicable<br>(hedian4) 1. Never had 2. No longer has 3. Did not have previously but has now -1. Not applicable<br>(hedias4) 1.Yes 2.No<br>(hedim01-08) 4.Congestive heart failure | Respondents were considered as currently having heart failure if they (1) reported heart failure in wave 1, did not dispute this record in wave 2, and reported still having it in wave 2 [hediac4=1 & hedias4=1], or (2) did not have it previously but reported it in wave 2 [hediac4!=1 & hedian4=3], or (3) did not report the condition in wave 1 but reported doctor diagnosis of heart failure in wave 2 [hediac4 = -1 & (hedim01 to hedim08=4)].<br>* Respondents with an issue with feedforward on this condition (n = 34) were randomly assigned based on |

| Outcome | Wave | (variable name) Question | Answer | Harmonisation |
|---|---|---|---|---|
| | | | | observed prevalence (see Stata .do file for additional details) |
| | 6 | (hedachf) Whether confirms diagnosis of heart failure recorded in previous wave(s)?<br>(hedanhf) Reason disputed heart failure diagnosis fed forward?<br>(hedashf) Do you still have heart failure?<br>(hedimhf) Heart failure diagnosis newly reported? | (hedachf) 1.Yes 2.No -1.Not Applicable<br>(hedanhf) 1.Never had 2.No longer has 3.Did not have previously but has now -1.N.A. -8.Don't know<br>(hedashf) 1. Yes 2.No<br>(hedimhf) 0.Not mentioned 1.Mentioned -9.Refusal -8.Don't know | Respondents were considered as currently having heart failure if they (a) confirmed previous doctor diagnosis of heart failure and still had heart failure [hedachf=1 & hedashf=1], or (2) did not report it previously but reported it in wave 6 [hedanhf=3], or (3) reported doctor diagnosis of heart failure since last interview [hedimhf=1]<br>* In wave 6, the prevalence of heart failure could only be calculated among Rs who were not in the refreshment sample (see Stata .do file for additional details). |
| | 11 | (hehavehf) | (hehavehf) 1.Still have heart failure but do not take medication or treatment for it 2.Still have heart failure and take medication or treatment for it 3. No longer have heart failure -1.Not applicable -8.Don't know | Respondents were considered as having heart failure if hehavehf=(1 or 2) |
| Self-reported current heart murmur | 2 | (hediac5) Whether confirms diagnosis of heart murmur recorded in wave 1?<br>(hedian5) Reason disputed diagnosis of heart murmur from wave 1<br>(hedias5) Do you still have heart murmur?<br>(hedim01-08) Has a doctor ever told you that you have the following cardiovascular conditions (1st to 8th mention, merged variable)? | (hediac5) 1.Yes 2.No -1.Not applicable<br>(hedian5) 1. Never had 2. No longer has 3. Did not have previously but has now -1. Not applicable<br>(hedias5) 1.Yes 2.No<br>(hedim01-08) 5.A heart murmur | Respondents were considered as currently having heart murmur if they (1) reported heart murmur in wave 1, did not dispute this record in wave 2, and reported still having it in wave 2 [hediac5=1 & hedias5=1], or (2) did not have it previously but reported it in wave 2 [hediac5!=1 & hedian5=3], or (3) did not report the condition in wave 1 but reported doctor diagnosis of heart murmur in wave 2 [hediac5 = -1 & (hedim01 to hedim08=5)].<br>* Respondents with an issue with feedforward on this condition (n = 191) were randomly assigned based on observed prevalence (see Stata .do file for additional details) |
| | 6 | (hedachm) Whether confirms diagnosis of heart murmur recorded in previous wave(s)?<br>(hedanhm) Reason disputed heart murmur diagnosis fed forward?<br>(hedashm) Do you still have heart murmur?<br>(hedimhm) Heart murmur diagnosis newly reported? | (hedachm) 1.Yes 2.No -1.Not Applicable<br>(hedanhm) 1.Never had 2.No longer has 3.Did not have previously but has now -1.N.A. -8.Don't know<br>(hedashm) 1. Yes 2.No<br>(hedimhm) 0.Not mentioned 1.Mentioned -9.Refusal -8.Don't know | Respondents were considered as currently having heart murmur if they (a) confirmed a previous doctor diagnosis of heart murmur and still had heart murmur [hedachm=1 & hedashm=1], or (2) did not report it previously but reported it in wave 6 [hedanhm=3], or (3) reported doctor diagnosis of heart murmur since last interview [hedimhm=1]<br>* In wave 6, the prevalence of heart murmur could only be calculated among Rs who were not in the refreshment sample (see Stata .do file for additional details). |
| | 11 | (hehavehm) Whether still has heart murmur? | (hehavehm) 1.Still have heart murmur but do not take medication or treatment for it 2.Still have heart murmur and take medication or treatment for it 3. No longer have heart murmur -1.Not applicable -8.Don't know | Respondents were considered as having heart murmur if hehavehm=(1 or 2) |
| Self-reported current abnormal heart rhythm | 2 | (hediac6) Whether confirms diagnosis of abnormal heart rhythm recorded in wave 1? | (hediac6) 1.Yes 2.No -1.Not applicable | Respondents were considered as currently having abnormal heart rhythm if they (1) reported abnormal heart rhythm in wave 1, did not dispute this record in wave 2, and reported still having it in wave 2 |

| Outcome | Wave | (variable name) Question | Answer | Harmonisation |
| --- | --- | --- | --- | --- |
| | | (hedian6) Reason disputed diagnosis of abnormal heart rhythm from wave 1<br>(hedias6) Do you still have abnormal heart rhythm?<br>(hedim01-08) Has a doctor ever told you that you have the following cardiovascular conditions (1st to 8th mention, merged variable)? | (hedian6) 1. Never had 2. No longer has 3. Did not have previously but has now -1. Not applicable<br>(hedias6) 1.Yes 2.No<br>(hedim01-08) 6.An abnormal heart rhythm | [hediac6=1 & hedias6=1], or (2) did not have it previously but reported it in wave 2 [hediac6!=1 & hedian6=3], or (3) did not report the condition in wave 1 but reported doctor diagnosis of abnormal heart rhythm in wave 2 [hediac6 = -1 & (hedim01/08=6)]. |
| | 6 | (hedacar) Whether confirms diagnosis of abnormal heart rhythm recorded in previous wave(s)?<br>(hedanar) Reason disputed abnormal heart rhythm diagnosis fed forward?<br>(hedasar) Do you still have abnormal heart rhythm?<br>(hedimar) Abnormal heart rhythm diagnosis newly reported? | (hedacar) 1.Yes 2.No -1.Not Applicable<br>(hedanar) 1.Never had 2.No longer has 3.Did not have previously but has now -1.N.A. -8.Don't know<br>(hedasar) 1. Yes 2.No<br>(hedimar) 0.Not mentioned 1.Mentioned -9.Refusal -8.Don't know | Respondents were considered as currently having abnormal heart rhythm if they (a) confirmed previous doctor diagnosis of abnormal heart rhythm and still had abnormal heart rhythm [hedacar=1 & hedasar=1], or (2) did not report it previously but reported it in wave 6 [hedanar=3], or (3) reported doctor diagnosis of abnormal heart rhythm since last interview [hedimar=1]<br>* In wave 6, the prevalence of abormal heart rhythm could only be calculated among Rs who were not in the refreshment sample (see Stata .do file for additional details). |
| | 11 | (hehaveah) Do you still have abnormal heart rhythm? | (hehaveah) 1.Still have abnormal heart rhythm but do not take medication or treatment for it 2.Still have abnormal heart rhythm and take medication or treatment for it 3. No longer have abnormal heart rhythm -1.N.A. -8.Don't know | Respondents were considered as having abnormal heart rhythm if hehaveah=(1 or 2) |
| Self-reported current angina | 2 | (hediac2) Whether confirms diagnosis of angina recorded in wave 1?<br>(hedian2) Reason disputed diagnosis of angina from wave 1<br>(hedias2) Do you still have angina?<br>(hedim01-08) Has a doctor ever told you that you have the following cardiovascular conditions (1st to 8th mention, merged variable)? | (hediac2) 1.Yes 2.No -1.Not applicable<br>(hedian2) 1. Never had 2. No longer has 3. Did not have previously but has now -1. Not applicable<br>(hedias2) 1.Yes 2.No<br>(hedim01-08) 2.Angina | Respondents were considered as currently having angina if they (1) reported angina in wave 1, did not dispute this record in wave 2, and reported still having it in wave 2 [hediac2=1 & hedias2=1], or (2) did not have it previously but reported it in wave 2 [hediac2!=1 & hedian2=3], or (3) did not report the condition in wave 1 but reported doctor diagnosis of angina in wave 2 [hediac2 = -1 & (hedim01/08=2)].<br>* Respondents with an issue with feedforward on this condition (n = 481) were randomly assigned based on observed prevalence (see Stata .do file for additional details) |
| | 6 | (hedacan) Whether confirms diagnosis of angina recorded in previous wave(s)?<br>(hedanan) Reason disputed angina diagnosis fed forward?<br>(hedasan) Do you still have angina?<br>(hediman) Angina diagnosis newly reported? | (hedacan) 1.Yes 2.No -1.Not Applicable<br>(hedanan) 1.Never had 2.No longer has 3.Did not have previously but has now -1.N.A. -8.Don't know<br>(hedasan) 1. Yes 2.No<br>(hediman) 0.Not mentioned 1.Mentioned -9.Refusal -8.Don't know | Respondents were considered as currently having angina if they (a) confirmed previous doctor diagnosis of angina and still had angina [hedacan=1 & hedasan=1], or (2) did not report it previously but reported it in wave 6 [hedanan=3], or (3) reported doctor diagnosis of angina since last interview [hediman=1]<br>* In wave 6, the prevalence of angina could only be calculated among Rs who were not in the refreshment sample (see Stata .do file for additional details). |
| | 11 | (hehavean) Whether still has angina? | (hehavean) 1.Still have heart murmur but do not take medication or treatment for it 2.Still have heart murmur and take medication or treatment for it | Respondents were considered as having angina if hehavean=(1 or 2) |

| Outcome | Wave | (variable name) Question | Answer | Harmonisation |
|---|---|---|---|---|
| | | | 3. No longer have heart murmur -1.Not applicable -8.Don't know | |
| Self-reported current heart attack | 2 | (hediac3) Whether confirms diagnosis of heart attack recorded in wave 1?<br>(hedian3) Reason disputed diagnosis of heart attack from wave 1<br>(hedim01-08) Has a doctor ever told you that you have the following cardiovascular conditions (1st to 8th mention, merged variable)?<br>(henmmi) Number of heart attacks since last visit? | (hediac3) 1.Yes 2.No -1.Not applicable<br>(hedian3) 1. Never had 2. No longer has 3. Did not have previously but has now -1. Not applicable<br>(hedim01-08) 3.A heart attack<br>Valid number | Respondents were considered as having recently had a heart attack if they (1) did not report in wave 1 but reported doctor diagnosis of a heart attack in wave 2 [hediac3!=1 & hedian3=3], or (2) did not have previously but reported it in wave 2 [hediac3 = -1 & (hedim01/08=3)], or (3) had >=1 heart attack since last visit [henmmi>=1]<br>*Rs with an issue with feedforward (n=260) could not be randomly assigned because there is no question "Do you still have a heart attack?" to calculate the prevalence among those not affected by this issue. |
| | 6 | (hedanmi) Reason disputed heart attack diagnosis fed forward?<br>(hedimmi) Heart attack diagnosis newly reported?<br>(henmmi) Number of heart attacks since last visit? | (hedanmi) 1.Never had 2.No longer has 3.Did not have previously but has now -1.N.A. -8.Don't know<br>(hedimmi) 0.Not mentioned 1.Mentioned -9.Refusal -8.Don't know<br>Valid number | Respondents were considered as having recently had a heart attack if they (1) did not have it previously but had it now [hedanmi=3], or (2) (for non-refreshment sample) mentioned diagnosis of a heart attack since last interview [hedimmi = 1], or (3) had >=1 heart attack in past 2 years [henmmi>=1] |
| | 11 | (hediacmi) Whether confirms previous diagnosis of a heart attack?<br>(heothha) Whether had another heart attack since last diagnosis?<br>(hediia3) New cardiac condition diagnosis: A heart attack (including myocardial infarction)<br>(hemedha) Whether taking any prescribed medication for heart attack? | 1.Yes 2.No, respondent has never had condition -1.Not applicable -8.Don't know<br>1.Yes 2.No -1.Not applicable -8.Don't know<br>0.Not mentioned 1.Mentioned -9.Refusal -8.Don't know<br>1.Yes 2.No -1.Not applicable -8.Don't know | Respondents were considered as having recently had a heart attack if they (1) did not dispute previous diagnosis of a heart attack and had another heart attack since last diagnosis [hediacmi=1 & heothha =1], or (2) did not report diagnosis of a heart attack previously but reported a diagnosis of heart attack in wave 11 [hediacmi=1 & hediia3 = 1] |
| Obesity | 2, 6, 11 | (bmiobe) (Derived) Valid BMI grouped according to WHO definitions (kg/m2) | 1.Under 18.5, underweight<br>2.18.5 or over but less than 25, normal range<br>3. 25 or over but less than 30, pre-obese<br>4. 30 or over but less than 35, obese class I<br>5. 35 or over but less than 40, obese class II<br>6. 40 or over, obese class III<br>-1. Not applicable | Obese if valid bmiobe >=4 |
| Elevated high-sensitivity C-reactive protein (hsCRP) | 2, 6, 11 | (hscrp) Blood CRP level (mg/L) | Valid values | Elevated C-reactive protein if hscrp >=3 |
| Self-reported arthritis | 2 | (hediad3) Whether confirms diagnosis of arthritis recorded in wave 1?<br>(hediam3) Reason disputed arthritis diagnosis from wave 1.<br>(hedids3) Do you still have arthritis?<br>(hedib01-hedib04) Has a doctor ever told you that you have the following conditions (1st to 4th mention)? | (hediad3) 1.Yes 2.No -1.Not Applicable<br>(hediam3) 1.Never had 2.No longer has 3.Did not have previously, but has now -1.N.A. -8.Don't know.<br>(hedids3) 1.Yes 2.No<br>(hedib01-hedib04) 3.Arthritis | Respondents were considered as currently having arthritis if they (a) reported arthritis in wave 1, did not dispute this record in wave 2, and reported still having it in wave 2 [hediad3=1 & hedids3=1], or (2) did not have it previously but reported it in wave 2 [hediad3 !=1 & hediam3=3], or (3) did not report the condition in wave 1 but reported doctor diagnosis of arthritis in wave 2 [hediad3 = -1 & (hedib01=3 \| hedib02=3 \| hedib03=3 \| hedib04=3)]<br>[Note: Only 3 Rs had the issue of feedforward error on arthritis in wave 2 so we ignored it]. |

| | | | | |
|---|---|---|---|---|
| | 6 | (hedbdar) Whether confirms diagnosis of arthritis recorded in previous wave(s)?<br>(hedbmar) Reason disputed arthritis diagnosis fed forward.<br>(hedbsar) Do you still have arthritis?<br>(hedibar) Has a doctor ever told you that you have arthritis? | (hedbdar) 1.Yes 2.No -1.Not Applicable<br>(hedbmar) 1.Never diagnosed 2.No longer has 3.Did not have previously but has now 4.Misdiagnosed -1. N/A<br>(hedbsar) 1.Yes 2.No<br>(hedibar) 1.Yes 2.No | Respondents were considered as currently having arthritis if they (a) reported arthritis in a prior wave, did not dispute this record in wave 6, and reported still having it in wave 6 [hedbdar=1 & hedbsar=1], or (2) did not report it previously but reported it in wave 6 [hedbmar=3], or (3) reported doctor diagnosis of arthritis since last interview [hedibar=1]<br>* In wave 6, the prevalence of arthritis could only be calculated among Rs who were not in the refreshment sample (see Stata .do file for additional details). |
| | 11 | (hehavear) Do you still have arthritis? | (hehavear) 1.Still have arthritis but do not take medication or treatment for it 2.Still have high arthritis and take medication or treatment for it 3.No longer have arthritis -1.Not applicable | Respondents were considered as currently having arthritis if hehavear=(1 or 2) |
| Self-reported current psychiatric problems | 2 | (hediad7) Whether confirms diagnosis of psychiatric problems recorded in wave 1?<br>(hediam7) Reason R disputed psychiatric problems diagnosis from wave 1<br>(hedids7) Do you still have psychiatric problems?<br>(hedib01-hedib04) Has a doctor ever told you that you have the following conditions (1st to 4th mention)?<br>(heyrc) Whether experienced any psychiatric problems in last 2 years? | (hediad7) 1.Yes 2.No -1.Not Applicable<br>(hediam7) 1.Never had 2.No longer has 3.Did not have previously but has now -1.Not applicable -8.Don't know<br>(hedids7) 1.Yes 2.No<br>(hedib01-hedib04) 7.Psychiatric problems<br>(heyrc) 1.Yes 2.No | Respondents were considered as having psychiatric problems if they (1) confirm previous diagnosis of psychiatric problems and still have psychiatric problems [hediad7=1 & hedids7=1], or (2) did not have them previously but have them in wave 2 [ediad7!=1 & hediam7=3], or (3) did not report diagnosis of psychiatric problems previously but reported diagnosis of psychiatric problems in wave 2 [hediad7=-1 & (hedib01 to hedib07=7)], or (4) experienced psychiatric problems in last 2 years [heyrc=1] |
| | 6 | (hedbdps) Whether confirms diagnosis of psychiatric problems recorded previously<br>(hedbmps) Reason R disputed psychiatric condition diagnosis fed forward<br>(hedibps) Has a doctor told you that you have psychiatric problems since last interview?<br>(heyrc) Whether experienced any psychiatric problems in last 2 years? | (hedbdps) 1.Yes 2.No -1.Not Applicable<br>(hedbmps) 1. Never had 2. No longer has 3.Did not have previously but has now 4. Misdiagnosed -1.Not applicable<br>(hedibps) 1.Yes 2.No<br>(heyrc) 1.Yes 2.No | Respondents were considered as having psychiatric problems if they (1) confirm previous diagnosis of psychiatric problems and experienced psychiatric problems in last 2 years [hedbdps=1 & heyrc=1], or (2) did not repot them previously but have them in wave 6 [hedbmps=3], or (3) reported doctor diagnosis of psychiatric problems since last interview [hedibps=1]<br>* In wave 6, the prevalence of hypertension could only be calculated among Rs who were not in the refreshment sample (see Stata .do file for additional details). |
| | 11 | (hehaveps) Do you still have psychiatric problems? | (hehaveps) 1.Still have psychiatric problems but do not take medication or treatment for it 2.Still have psychiatric problems and take medication or treatment for it 3. No longer have psychiatric problems -1.Not applicable -8.Don't know | Respondents were considered as having psychiatric problems if hehaveps=(1 or 2) |

| | | | | |
|---|---|---|---|---|
| Depressive symptoms (CES-D-8) and depression | 2, 6, 11 | Much of the time durin the past week<br>(psceda) you felt depressed?<br>(pscedb) you felt everything you did was an effort?<br>(pscedc) your sleep was restless?<br>(pscedd) you were happy?<br>(pscede) you felt lonely?<br>(pscedf) you enjoyed life?<br>(pscedg) you felt sad?<br>(pscedh) you could not get going? | 1.Yes<br>2.No<br>-9.Refusal<br>-8.Don't know<br>-1.Not applicable | We recoded pscedd and pscedf: 0=yes, 1=no.<br>The other six variables were coded: 0=no, 1=yes.<br><br>Depressive symptoms (CES-D-8) is the sum of all valid values from these 8 variables.<br>Depression is established if CES-D-8 >=4. |
| CASP-19 (Control, Autonomy, Self-Realisation, and Pleasure) | 2, 6, 11 | How often do you fell<br>(scqola) My age prevents me from doing the things I would like to<br>(scqolb) I feel that what happens to me is out of my control<br>(scqolc) I feel free to plan for the future<br>(scqold) I feel left out of things<br>(scqole) I can do the things that I want to do<br>(scqolf) Family responsibilities prevent me from doing what I want to do<br>(scqolg) I feel I can please myself with what I do<br>(scqolh) My health stops me from doing the things I would like to<br>(scqoli) Shortage of money stops me from doing things I want to do<br>(scqolj) I look forward to each day<br>(scqolk) I feel that my life has meaning<br>(scqoll) I enjoy the things that I do<br>(scqolm) I enjoy being in the company of others<br>(scqoln) On balance, I look back on my life with a sense of happiness<br>(scqolo) I feel full of energy these days<br>(scqolp) I choose to do things that I have never done before<br>(scqolq) I feel satisfied with the way my life has turned out<br>(scqolr) I feel that life is full of opportunities<br>(scqols) I feel the future looks good for me | 1.Often<br>2.Sometimes<br>3.Not often<br>4.Never<br>-9.Not answered<br>-1.Not applicable | Answers for scqola scqolb scqold scqolf scqolh scqoli were recoded as 4=3, 3=2, 2=1, 1=0.<br>Answers for scqolc scqole scqolg scqolj scqolk scqoll scqolm scqoln scqolo scqolp scqolq scqolr scqols were recoded as 1=3, 3=1, 4=0.<br><br>CASP-19 is the sum of all valid values from these 19 variables (range:0-57). |
| Memory | 2, 6, 11 | (cflisen) number of words recalled immediately (out of 10)<br>(cflisd) number of words recalled after delay (out of 10) | (cflisen) Valid values 0-10<br><br>(cflisd) Valid values 0-10 | Memory is the sum of immediate recall score and delayed recall score (cflisen+cflisd) |

Note: Data source: English Longitudinal Study of Ageing (ELSA) waves 2 (2004/05), 6 (2012/13), and 11 (2023/24).

### Table S3. Weighted results from Poisson and linear regression models testing the statistical effect of time (wave) on health outcomes

***Panel A. Self-rated health, pain, and functional limitations***

| | Poor/fair self-rated health | | | Limiting long-standing illness | | | Any pain | | | Moderate/severe pain | | | Any ADL limitations | | | Any IADL limitations | | | Any mobility impairments | | |
|---|---|---|---|---|---|---|---|---|---|---|---|---|---|---|---|---|---|---|---|---|---|
| | RR | 95%CI | p | RR | 95%CI | p | RR | 95%CI | p | RR | 95%CI | p | RR | 95%CI | p | RR | 95%CI | p | RR | 95%CI | p |
| Wave (ref:w2) | | | <0.001 | | | 0.024 | | | <0.001 | | | <0.001 | | | 0.001 | | | <0.001 | | | <0.001 |
| Wave 6 | 1.04 | [1.00,1.09] | 0.071 | 1.01 | [0.97,1.05] | 0.721 | 1.14*** | [1.10,1.19] | <0.001 | 1.11*** | [1.06,1.16] | <0.001 | 0.90*** | [0.85,0.96] | <0.001 | 0.89*** | [0.85,0.96] | <0.001 | 0.92*** | [0.90,0.95] | <0.001 |
| Wave 11 | 1.27*** | [1.20,1.35] | <0.001 | 1.07** | [1.02,1.13] | 0.009 | 1.27*** | [1.21,1.33] | <0.001 | 1.23*** | [1.15,1.30] | <0.001 | 1.00 | [0.92,1.08] | 0.917 | 0.98 | [0.92,1.08] | 0.612 | 0.90*** | [0.87,0.94] | <0.001 |
| Age (ref: 50-59) | | | <0.001 | | | <0.001 | | | <0.001 | | | <0.001 | | | <0.001 | | | <0.001 | | | <0.001 |
| 60-69 | 1.21*** | [1.13,1.29] | <0.001 | 1.25*** | [1.18,1.33] | <0.001 | 1.11*** | [1.06,1.17] | <0.001 | 1.24*** | [1.17,1.32] | <0.001 | 1.40*** | [1.27,1.54] | <0.001 | 1.29*** | [1.18,1.41] | <0.001 | 1.29*** | [1.24,1.34] | <0.001 |
| 70-79 | 1.43*** | [1.34,1.52] | <0.001 | 1.51*** | [1.43,1.61] | <0.001 | 1.13*** | [1.07,1.19] | <0.001 | 1.31*** | [1.23,1.40] | <0.001 | 1.88*** | [1.71,2.06] | <0.001 | 1.69*** | [1.55,1.85] | <0.001 | 1.56*** | [1.50,1.62] | <0.001 |
| 80+ | 1.64*** | [1.52,1.77] | <0.001 | 1.85*** | [1.73,1.97] | <0.001 | 1.12*** | [1.05,1.19] | <0.001 | 1.30*** | [1.20,1.40] | <0.001 | 2.65*** | [2.40,2.93] | <0.001 | 3.02*** | [2.76,3.31] | <0.001 | 1.85*** | [1.77,1.93] | <0.001 |
| Women | 0.94* | [0.89, 0.98] | 0.010 | 1.06* | [1.01,1.10] | 0.013 | 1.20*** | [1.15,1.25] | <0.001 | 1.32*** | [1.25,1.39] | <0.001 | 1.09* | [1.02,1.16] | 0.011 | 1.31*** | [1.23,1.39] | <0.001 | 1.28*** | [1.24,1.32] | <0.001 |
| Education (ref:High) | | | <0.001 | | | <0.001 | | | <0.001 | | | <0.001 | | | <0.001 | | | <0.001 | | | <0.001 |
| Medium | 1.39*** | [1.26,1.52] | <0.001 | 1.23*** | [1.14,1.32] | <0.001 | 1.25*** | [1.17,1.33] | <0.001 | 1.41*** | [1.29,1.55] | <0.001 | 1.46*** | [1.30,1.65] | <0.001 | 1.43*** | [1.28,1.60] | <0.001 | 1.26*** | [1.20,1.33] | <0.001 |
| Low | 1.93*** | [1.75,2.13] | <0.001 | 1.36*** | [1.25,1.47] | <0.001 | 1.39*** | [1.29,1.49] | <0.001 | 1.70*** | [1.54,1.87] | <0.001 | 1.69*** | [1.48,1.92] | <0.001 | 1.68*** | [1.49,1.90] | <0.001 | 1.35*** | [1.28,1.43] | <0.001 |
| Wealth (ref:Pooresr) | | | <0.001 | | | <0.001 | | | <0.001 | | | <0.001 | | | <0.001 | | | <0.001 | | | <0.001 |
| 2nd | 0.77*** | [0.72,0.81] | <0.001 | 0.79*** | [0.75,0.84] | <0.001 | 0.86*** | [0.82,0.91] | <0.001 | 0.80*** | [0.75,0.85] | <0.001 | 0.72*** | [0.67,0.79] | <0.001 | 0.69*** | [0.64,0.74] | <0.001 | 0.85*** | [0.82,0.88] | <0.001 |
| 3rd | 0.58*** | [0.55,0.62] | <0.001 | 0.63*** | [0.59,0.67] | <0.001 | 0.78*** | [0.74,0.83] | <0.001 | 0.68*** | [0.64,0.73] | <0.001 | 0.57*** | [0.52,0.62] | <0.001 | 0.54*** | [0.50,0.58] | <0.001 | 0.78*** | [0.75,0.81] | <0.001 |
| 4th | 0.48*** | [0.44,0.52] | <0.001 | 0.56*** | [0.53,0.60] | <0.001 | 0.69*** | [0.65,0.73] | <0.001 | 0.57*** | [0.53,0.61] | <0.001 | 0.47*** | [0.43,0.52] | <0.001 | 0.44*** | [0.40,0.48] | <0.001 | 0.74*** | [0.71,0.77] | <0.001 |
| 5th, wealthiest | 0.36*** | [0.33,0.39] | <0.001 | 0.52*** | [0.48,0.55] | <0.001 | 0.62*** | [0.58,0.66] | <0.001 | 0.46*** | [0.42,0.50] | <0.001 | 0.35*** | [0.31,0.39] | <0.001 | 0.33*** | [0.30,0.37] | <0.001 | 0.65*** | [0.61,0.68] | <0.001 |
| N | 22,115 | | | 22,120 | | | 22,124 | | | 22,117 | | | 22,125 | | | 22,125 | | | 22,126 | | |

***Panel B. Chronic conditions, obesity, and inflammation***

| | Self-reported hypertension | | | Hypertension biomarker | | | Self-reported diabetes | | | Diabetes biomarkers | | | Self-reported high cholesterol | | | High cholesterol biomarker | | | Self-reported lung disease | | |
|---|---|---|---|---|---|---|---|---|---|---|---|---|---|---|---|---|---|---|---|---|---|
| | RR | 95%CI | p | RR | 95%CI | p | RR | 95%CI | p | RR | 95%CI | p | RR | 95%CI | p | RR | 95%CI | p | RR | 95%CI | p |
| Wave (ref:w2) | | | <0.001 | | | <0.001 | | | <0.001 | | | <0.001 | | | 0.001 | | | <0.001 | | | 0.011 |
| Wave 6 | 1.04* | [1.01,1.08] | 0.025 | 0.83*** | [0.79,0.87] | <0.001 | 1.40*** | [1.29,1.52] | <0.001 | 1.66*** | [1.46,1.88] | <0.001 | 1.67*** | [1.58,1.76] | <0.001 | 0.85*** | [0.83,0.88] | <0.001 | 0.98 | [0.87,1.11] | 0.791 |
| Wave 11 | 1.15*** | [1.09,1.21] | <0.001 | 0.79*** | [0.73,0.85] | <0.001 | 1.56*** | [1.39,1.76] | <0.001 | 1.58*** | [1.32,1.89] | <0.001 | 2.04*** | [1.92,2.18] | <0.001 | 0.69*** | [0.66,0.73] | <0.001 | 1.25* | [1.06,1.49] | 0.010 |
| Age (ref: 50-59) | | | <0.001 | | | <0.001 | | | <0.001 | | | <0.001 | | | <0.001 | | | <0.001 | | | <0.001 |
| 60-69 | 1.37*** | [1.28,1.46] | <0.001 | 1.30*** | [1.21,1.40] | <0.001 | 1.60*** | [1.39,1.84] | <0.001 | 1.90*** | [1.57,2.31] | <0.001 | 1.48*** | [1.37,1.60] | <0.001 | 0.89*** | [0.86,0.91] | <0.001 | 1.70*** | [1.39,2.09] | <0.001 |
| 70-79 | 1.80*** | [1.68,1.91] | <0.001 | 1.59*** | [1.48,1.72] | <0.001 | 2.06*** | [1.79,2.38] | <0.001 | 2.30*** | [1.87,2.82] | <0.001 | 1.72*** | [1.59,1.85] | <0.001 | 0.76*** | [0.73,0.78] | <0.001 | 2.06*** | [1.67,2.54] | <0.001 |
| 80+ | 1.88*** | [1.75,2.02] | <0.001 | 1.61*** | [1.48,1.76] | <0.001 | 1.94*** | [1.64,2.29] | <0.001 | 2.12*** | [1.65,2.72] | <0.001 | 1.32*** | [1.21,1.45] | <0.001 | 0.68*** | [0.64,0.72] | <0.001 | 1.84*** | [1.44,2.35] | <0.001 |
| Women | 0.92*** | [0.88,0.96] | <0.001 | 0.94* | [0.90,0.99] | 0.027 | 0.71*** | [0.64,0.79] | <0.001 | 0.73*** | [0.64,0.84] | <0.001 | 1.00 | [0.95,1.05] | 0.857 | 1.31*** | [1.27,1.34] | <0.001 | 0.75*** | [0.65,0.86] | <0.001 |
| Education (ref:High) | | | <0.001 | | | 0.002 | | | 0.002 | | | 0.034 | | | <0.001 | | | <0.001 | | | <0.001 |
| Medium | 1.16*** | [1.08,1.24] | <0.001 | 1.11* | [1.02,1.20] | 0.013 | 1.21* | [1.03,1.43] | 0.019 | 1.28* | [1.03,1.59] | 0.028 | 1.12** | [1.04,1.20] | 0.002 | 0.92*** | [0.89,0.96] | <0.001 | 1.84*** | [1.43,2.37] | <0.001 |
| Low | 1.24*** | [1.15,1.33] | <0.001 | 1.17*** | [1.07,1.28] | <0.001 | 1.37*** | [1.15,1.64] | <0.001 | 1.40** | [1.08,1.80] | 0.010 | 1.20*** | [1.10,1.31] | <0.001 | 0.90*** | [0.86,0.94] | <0.001 | 2.30*** | [1.74,3.03] | <0.001 |
| Wealth (ref:Pooresr) | | | <0.001 | | | 0.003 | | | <0.001 | | | <0.001 | | | 0.002 | | | <0.001 | | | <0.001 |
| 2nd | 0.96 | [0.90,1.02] | 0.207 | 1.00 | [0.92,1.08] | 0.936 | 0.70*** | [0.62,0.80] | <0.001 | 0.71*** | [0.59,0.86] | <0.001 | 1.02 | [0.94,1.10] | 0.650 | 1.06* | [1.01,1.11] | 0.019 | 0.68*** | [0.58,0.81] | <0.001 |
| 3rd | 0.89*** | [0.84,0.95] | <0.001 | 0.90* | [0.83,0.98] | 0.018 | 0.60*** | [0.52,0.68] | <0.001 | 0.58*** | [0.48,0.69] | <0.001 | 0.92* | [0.85,1.00] | 0.041 | 1.06* | [1.01,1.11] | 0.014 | 0.48*** | [0.39,0.58] | <0.001 |
| 4th | 0.87*** | [0.81,0.93] | <0.001 | 0.97 | [0.89,1.05] | 0.479 | 0.49*** | [0.42,0.57] | <0.001 | 0.42*** | [0.35,0.51] | <0.001 | 0.93 | [0.86,1.01] | 0.084 | 1.10*** | [1.06,1.15] | <0.001 | 0.37*** | [0.30,0.46] | <0.001 |
| 5th, wealthiest | 0.78*** | [0.73,0.84] | <0.001 | 0.88** | [0.81,0.97] | 0.006 | 0.40*** | [0.34,0.47] | <0.001 | 0.37*** | [0.29,0.46] | <0.001 | 0.89** | [0.82,0.96] | 0.004 | 1.12*** | [1.07,1.17] | <0.001 | 0.28*** | [0.22,0.37] | <0.001 |
| N | 21,311 | | | 17,213 | | | 21,360 | | | 14,271 | | | 21,256 | | | 14,473 | | | 21,364 | | |

| | Self-reported stroke | | | Self-reported other heart diseases | | | Obesity biomarker | | | Elevated C-reactive protein | | | Self-reported Arthritis | | |
|---|---|---|---|---|---|---|---|---|---|---|---|---|---|---|---|
| | RR | 95%CI | p | RR | 95%CI | p | RR | 95%CI | p | RR | 95%CI | p | RR | 95%CI | p |
| Wave (ref:w2) | | | 0.561 | | | 0.004 | | | <0.001 | | | <0.001 | | | <0.001 |
| Wave 6 | 1.03 | [0.78,1.35] | 0.856 | 1.03 | [0.96,1.10] | 0.426 | 1.15*** | [1.09,1.20] | <0.001 | 0.80*** | [0.76,0.84] | <0.001 | 1.08*** | [1.04,1.12] | <0.001 |
| Wave 11 | 0.87 | [0.65,1.17] | 0.356 | 1.16** | [1.06,1.27] | 0.001 | 1.22*** | [1.14,1.31] | <0.001 | 0.66*** | [0.61,0.73] | <0.001 | 1.06* | [1.01,1.11] | 0.025 |
| Age (ref: 50-59) | | | <0.001 | | | <0.001 | | | <0.001 | | | <0.001 | | | <0.001 |
| 60-69 | 1.39 | [0.86,2.23] | 0.178 | 1.57*** | [1.39,1.78] | <0.001 | 1.04 | [0.98,1.11] | 0.157 | 1.14*** | [1.06,1.22] | <0.001 | 1.58*** | [1.48,1.68] | <0.001 |
| 70-79 | 2.43*** | [1.56,3.80] | <0.001 | 2.39*** | [2.11,2.70] | <0.001 | 0.93* | [0.87,0.99] | 0.029 | 1.26*** | [1.16,1.36] | <0.001 | 1.91*** | [1.79,2.03] | <0.001 |
| 80+ | 4.63*** | [2.94,7.29] | <0.001 | 3.60*** | [3.16,4.09] | <0.001 | 0.63*** | [0.56,0.70] | <0.001 | 1.25*** | [1.13,1.38] | <0.001 | 2.03*** | [1.89,2.18] | <0.001 |
| Women | 0.91 | [0.73,1.15] | 0.438 | 0.84*** | [0.78,0.90] | <0.001 | 1.11*** | [1.05,1.18] | <0.001 | 1.19*** | [1.12,1.26] | <0.001 | 1.45*** | [1.38,1.52] | <0.001 |
| Education (ref:High) | | | 0.608 | | | 0.257 | | | <0.001 | | | <0.001 | | | <0.001 |
| Medium | 1.10 | [0.77,1.56] | 0.608 | 1.05 | [0.94,1.17] | 0.376 | 1.24*** | [1.13,1.36] | <0.001 | 1.21*** | [1.10,1.34] | <0.001 | 1.18*** | [1.10,1.26] | <0.001 |
| Low | 1.21 | [0.82,1.79] | 0.346 | 0.98 | [0.87,1.11] | 0.765 | 1.47*** | [1.33,1.64] | <0.001 | 1.40*** | [1.25,1.57] | <0.001 | 1.23*** | [1.14,1.33] | <0.001 |
| Wealth (ref:Pooresr) | | | <0.001 | | | <0.001 | | | <0.001 | | | <0.001 | | | <0.001 |
| 2nd | 0.64** | [0.46,0.89] | 0.007 | 0.87** | [0.78,0.97] | 0.009 | 0.85*** | [0.79,0.92] | <0.001 | 0.78*** | [0.72,0.85] | <0.001 | 0.85*** | [0.81,0.90] | <0.001 |
| 3rd | 0.65* | [0.47,0.92] | 0.014 | 0.75*** | [0.67,0.84] | <0.001 | 0.78*** | [0.72,0.85] | <0.001 | 0.71*** | [0.65,0.77] | <0.001 | 0.76*** | [0.72,0.81] | <0.001 |
| 4th | 0.48*** | [0.33,0.70] | <0.001 | 0.69*** | [0.62,0.78] | <0.001 | 0.68*** | [0.63,0.75] | <0.001 | 0.68*** | [0.62,0.74] | <0.001 | 0.74*** | [0.69,0.79] | <0.001 |
| 5th, wealthiest | 0.48*** | [0.32,0.71] | <0.001 | 0.59*** | [0.52,0.67] | <0.001 | 0.60*** | [0.55,0.66] | <0.001 | 0.57*** | [0.51,0.63] | <0.001 | 0.64*** | [0.60,0.69] | <0.001 |
| N | 21,351 | | | 21,372 | | | 18,160 | | | 14,468 | | | 21,348 | | |

***Panel C. Mental health and cognition***

| | Self-reported psychiatric problems | | | Depression | | | Depressive symptoms | | | CASP-19 | | | Memory | | |
|---|---|---|---|---|---|---|---|---|---|---|---|---|---|---|---|
| | RR | 95%CI | p | RR | 95%CI | p | Beta | 95%CI | p | Beta | 95%CI | p | Beta | 95%CI | p |
| Wave (ref:w2) | | | <0.001 | | | <0.001 | | | <0.001 | | | <0.001 | | | <0.001 |
| Wave 6 | 1.14* | [1.02,1.27] | 0.021 | 1.00 | [0.92,1.07] | 0.915 | -0.09** | [-0.15,-0.03] | 0.003 | -1.80*** | [-2.11,-1.49] | <0.001 | 0.49*** | [0.39,0.59] | <0.001 |
| Wave 11 | 1.56*** | [1.36,1.79] | <0.001 | 1.19*** | [1.08,1.31] | <0.001 | 0.13** | [0.05,0.21] | 0.002 | -2.03*** | [-2.49,-1.58] | <0.001 | 0.58*** | [0.44,0.72] | <0.001 |
| Age (ref: 50-59) | | | <0.001 | | | <0.001 | | | <0.001 | | | <0.001 | | | <0.001 |
| 60-69 | 0.85** | [0.76,0.96] | 0.009 | 0.84*** | [0.77,0.92] | <0.001 | -0.16*** | [-0.23,-0.08] | <0.001 | 0.20 | [-0.18,0.58] | 0.305 | -0.66*** | [-0.78,-0.53] | <0.001 |
| 70-79 | 0.51*** | [0.44,0.59] | <0.001 | 0.85*** | [0.77,0.93] | <0.001 | -0.13** | [-0.21,-0.05] | 0.002 | -0.75*** | [-1.19,-0.32] | <0.001 | -1.85*** | [-1.98,-1.71] | <0.001 |
| 80+ | 0.33*** | [0.26,0.41] | <0.001 | 0.98 | [0.88,1.10] | 0.726 | 0.14* | [0.03,0.24] | 0.010 | -3.87*** | [-4.45,-3.29] | <0.001 | -3.92*** | [-4.10,-3.75] | <0.001 |
| Women | 1.59*** | [1.40,1.80] | <0.001 | 1.44*** | [1.33,1.56] | <0.001 | 0.40*** | [0.34,0.47] | <0.001 | 0.28 | [-0.08,0.63] | 0.129 | 0.87*** | [0.76,0.98] | <0.001 |
| Education (ref:High) | | | 0.409 | | | 0.409 | | | <0.001 | | | <0.001 | | | <0.001 |
| Medium | 1.00 | [0.85,1.17] | 0.956 | 1.30*** | [1.15,1.48] | <0.001 | 0.22*** | [0.14,0.30] | <0.001 | -1.48*** | [-1.93,-1.04] | <0.001 | -0.89*** | [-1.04,-0.74] | <0.001 |
| Low | 0.90 | [0.74,1.10] | 0.317 | 1.77*** | [1.54,2.04] | <0.001 | 0.58*** | [0.47,0.68] | <0.001 | -3.42*** | [-3.99,-2.85] | <0.001 | -2.31*** | [-2.50,-2.13] | <0.001 |
| Wealth (ref:Pooresr) | | | <0.001 | | | <0.001 | | | <0.001 | | | <0.001 | | | <0.001 |
| 2nd | 0.58*** | [0.50,0.67] | <0.001 | 0.64*** | [0.58,0.71] | <0.001 | -0.65*** | [-0.77,-0.53] | <0.001 | 3.60*** | [2.98,4.22] | <0.001 | 0.50*** | [0.33,0.67] | <0.001 |
| 3rd | 0.44*** | [0.37,0.52] | <0.001 | 0.52*** | [0.47,0.58] | <0.001 | -0.87*** | [-0.99,-0.76] | <0.001 | 5.68*** | [5.08,6.28] | <0.001 | 0.89*** | [0.71,1.07] | <0.001 |
| 4th | 0.39*** | [0.33,0.46] | <0.001 | 0.43*** | [0.38,0.48] | <0.001 | -1.07*** | [-1.18,-0.95] | <0.001 | 6.83*** | [6.23,7.43] | <0.001 | 1.29*** | [1.12,1.47] | <0.001 |
| 5th, wealthiest | 0.31*** | [0.26,0.37] | <0.001 | 0.33*** | [0.29,0.38] | <0.001 | -1.20*** | [-1.31,-1.09] | <0.001 | 8.47*** | [7.89,9.05] | <0.001 | 1.55*** | [1.37,1.72] | <0.001 |
| N | 21,350 | | | 21,992 | | | 21,992 | | | 20,055 | | | 21,929 | | |

Data source: English Longitudinal Study of Ageing waves 2, 6 and 11. Notes: RR: risk ratio; CI: confidence interval. P-values for overall wave, age group, education, and wealth correspond to the omnibus test. The outcome "other heart diseases" was established if the respondent reported one of the following: heart failure, heart murmur, heart attack, abnormal heart rhythm, or angina. Depending on health outcomes, different sampling weights were applied (cross-sectional weight, self-completion weight, nurse-visit weight, or blood-sample weight). P values: ***<0.001; **<0.01; *<0.05.

#### Table S4. Weighted results from Poisson or linear regression models testing the statistical effect of time (wave), age group, and their interaction on health outcomes

***Panel A. Self-rated health, pain, and physical functioning***

| | Poor/fair self-rated health | | | Limiting long-standing illness | | | Any pain | | | Moderate/severe pain | | | Any ADL limitations | | | Any IADL limitations | | | Any mobility impairments | | |
|---|---|---|---|---|---|---|---|---|---|---|---|---|---|---|---|---|---|---|---|---|---|
| | RR | 95%CI | p | RR | 95%CI | p | RR | 95%CI | p | RR | 95%CI | p | RR | 95%CI | p | RR | 95%CI | p | RR | 95%CI | p |
| Wave (ref: w2) | | | <0.001 | | | 0.061 | | | <0.001 | | | 0.233 | | | 0.291 | | | 0.007 | | | <0.001 |
| Wave 6 | 0.90 | [0.80,1.02] | 0.091 | 0.89* | [0.80,0.99] | 0.030 | 1.12* | [1.03,1.22] | 0.011 | 1.06 | [0.95,1.19] | 0.313 | 0.88 | [0.74,1.05] | 0.150 | 0.77** | [0.66,0.91] | 0.002 | 0.84*** | [0.77,0.90] | <0.001 |
| Wave 11 | 1.24** | [1.08,1.42] | 0.002 | 1.02 | [0.89,1.16] | 0.774 | 1.26*** | [1.14,1.39] | <0.001 | 1.13 | [0.98,1.30] | 0.103 | 1.02 | [0.82,1.27] | 0.838 | 0.99 | [0.81,1.21] | 0.925 | 0.79*** | [0.72,0.88] | <0.001 |
| Age (ref: 50-59) | | | <0.001 | | | <0.001 | | | 0.027 | | | <0.001 | | | <0.001 | | | <0.001 | | | <0.001 |
| 60-69 | 1.15** | [1.05,1.26] | 0.003 | 1.19*** | [1.09,1.29] | <0.001 | 1.11** | [1.03,1.19] | 0.006 | 1.21*** | [1.10,1.32] | <0.001 | 1.40*** | [1.23,1.59] | <0.001 | 1.23*** | [1.09,1.39] | <0.001 | 1.23*** | [1.16,1.29] | <0.001 |
| 70-79 | 1.31*** | [1.19,1.44] | <0.001 | 1.40*** | [1.29,1.52] | <0.001 | 1.11* | [1.02,1.19] | 0.010 | 1.24*** | [1.12,1.36] | <0.001 | 1.92*** | [1.69,2.19] | <0.001 | 1.61*** | [1.43,1.81] | <0.001 | 1.43*** | [1.36,1.51] | <0.001 |
| 80+ | 1.45*** | [1.30,1.61] | <0.001 | 1.63*** | [1.49,1.79] | <0.001 | 1.08 | [0.98,1.19] | 0.107 | 1.18** | [1.05,1.33] | 0.006 | 2.56*** | [2.24,2.93] | <0.001 | 2.78*** | [2.48,3.13] | <0.001 | 1.62*** | [1.53,1.71] | <0.001 |
| Wave (ref:w2) x Age (ref: 50-59) | | | 0.040 | | | 0.035 | | | 0.714 | | | 0.432 | | | 0.691 | | | 0.246 | | | <0.001 |
| W6 x 60-69 | 1.16 | [1.00,1.36] | 0.055 | 1.14 | [0.99,1.30] | 0.065 | 1.03 | [0.93,1.15] | 0.558 | 1.04 | [0.90,1.21] | 0.565 | 1.00 | [0.80,1.24] | 0.983 | 1.14 | [0.93,1.41] | 0.207 | 1.07 | [0.98,1.18] | 0.150 |
| W6 x 70-79 | 1.22** | [1.05,1.41] | 0.008 | 1.19** | [1.04,1.35] | 0.010 | 1.03 | [0.92,1.15] | 0.570 | 1.06 | [0.92,1.23] | 0.393 | 1.00 | [0.81,1.23] | 0.990 | 1.20 | [0.99,1.46] | 0.062 | 1.15** | [1.05,1.25] | 0.002 |
| W6 x 80+ | 1.32*** | [1.12,1.54] | <0.001 | 1.27*** | [1.10,1.45] | <0.001 | 1.03 | [0.90,1.17] | 0.653 | 1.10 | [0.93,1.30] | 0.267 | 1.11 | [0.90,1.37] | 0.330 | 1.26* | [1.04,1.52] | 0.018 | 1.23*** | [1.13,1.34] | <0.001 |
| W11 x 60-69 | 0.96 | [0.81,1.14] | 0.661 | 1.01 | [0.86,1.18] | 0.914 | 0.97 | [0.86,1.10] | 0.667 | 1.05 | [0.89,1.25] | 0.546 | 1.01 | [0.78,1.31] | 0.938 | 1.00 | [0.78,1.28] | 0.984 | 1.13 | [1.00,1.27] | 0.058 |
| W11 x 70-79 | 1.06 | [0.89,1.25] | 0.532 | 1.07 | [0.91,1.25] | 0.420 | 1.03 | [0.91,1.17] | 0.668 | 1.13 | [0.95,1.35] | 0.160 | 0.90 | [0.70,1.16] | 0.402 | 0.94 | [0.74,1.19] | 0.616 | 1.18** | [1.05,1.32] | 0.006 |
| W11 x 80+ | 1.10 | [0.92,1.31] | 0.292 | 1.16 | [0.99,1.36] | 0.066 | 1.08 | [0.94,1.25] | 0.297 | 1.24* | [1.02,1.50] | 0.028 | 1.00 | [0.78,1.29] | 0.992 | 1.01 | [0.80,1.27] | 0.929 | 1.27*** | [1.13,1.43] | <0.001 |
| N | 22,115 | | | 22,120 | | | 22,124 | | | 22,117 | | | 22,125 | | | 22,125 | | | 22,126 | | |

***Panel B. Chronic conditions, obesity, and inflammation***

| | Self-reported hypertension | | | Hypertension biomarker | | | Self-reported diabetes | | | Diabetes biomarkers | | | Self-reported high cholesterol | | | High cholesterol biomarker | | | Self-reported lung disease | | |
|---|---|---|---|---|---|---|---|---|---|---|---|---|---|---|---|---|---|---|---|---|---|
| | RR | 95%CI | p | RR | 95%CI | p | RR | 95%CI | p | RR | 95%CI | p | RR | 95%CI | p | RR | 95%CI | p | RR | 95%CI | p |
| Wave (ref: w2) | | | 0.454 | | | 0.007 | | | 0.004 | | | 0.052 | | | <0.001 | | | <0.001 | | | 0.425 |
| Wave 6 | 0.98 | [0.86,1.12] | 0.807 | 0.84** | [0.73,0.96] | 0.010 | 1.53** | [1.16,2.02] | 0.003 | 1.53* | [1.08,2.17] | 0.016 | 1.48*** | [1.28,1.73] | <0.001 | 0.91*** | [0.87,0.95] | <0.001 | 0.77 | [0.51,1.18] | 0.231 |
| Wave 11 | 0.91 | [0.78,1.06] | 0.209 | 0.80* | [0.66,0.97] | 0.024 | 1.44* | [1.05,1.98] | 0.025 | 1.16 | [0.68,1.96] | 0.593 | 1.41*** | [1.19,1.67] | <0.001 | 0.86*** | [0.81,0.92] | <0.001 | 1.05 | [0.67,1.66] | 0.821 |
| Age (ref: 50-59) | | | <0.001 | | | <0.001 | | | <0.001 | | | <0.001 | | | <0.001 | | | <0.001 | | | <0.001 |
| 60-69 | 1.30*** | [1.19,1.41] | <0.001 | 1.29*** | [1.18,1.42] | <0.001 | 1.57*** | [1.28,1.94] | <0.001 | 1.90*** | [1.43,2.52] | <0.001 | 1.31*** | [1.17,1.47] | <0.001 | 0.95** | [0.92,0.99] | 0.004 | 1.57*** | [1.20,2.05] | <0.001 |
| 70-79 | 1.62*** | [1.48,1.76] | <0.001 | 1.60*** | [1.46,1.76] | <0.001 | 2.16*** | [1.76,2.65] | <0.001 | 1.91*** | [1.41,2.58] | <0.001 | 1.40*** | [1.24,1.58] | <0.001 | 0.86*** | [0.82,0.89] | <0.001 | 1.67*** | [1.28,2.20] | <0.001 |
| 80+ | 1.48*** | [1.34,1.64] | <0.001 | 1.68*** | [1.51,1.88] | <0.001 | 1.71*** | [1.32,2.20] | <0.001 | 1.50* | [1.00,2.24] | 0.049 | 0.67*** | [0.55,0.81] | <0.001 | 0.78*** | [0.73,0.83] | <0.001 | 1.68** | [1.22,2.31] | 0.002 |
| Wave (ref:w2) x Age (ref: 50-59) | | | <0.001 | | | 0.863 | | | 0.480 | | | 0.198 | | | <0.001 | | | <0.001 | | | 0.652 |
| W6 x 60-69 | 0.99 | [0.85,1.15] | 0.883 | 1.01 | [0.86,1.19] | 0.874 | 0.88 | [0.63,1.24] | 0.463 | 0.90 | [0.59,1.35] | 0.601 | 1.05 | [0.88,1.25] | 0.611 | 0.95 | [0.90,1.01] | 0.097 | 1.27 | [0.78,2.07] | 0.338 |
| W6 x 70-79 | 1.07 | [0.92,1.24] | 0.375 | 0.98 | [0.84,1.14] | 0.774 | 0.85 | [0.62,1.16] | 0.309 | 1.21 | [0.80,1.83] | 0.370 | 1.12 | [0.94,1.34] | 0.213 | 0.88*** | [0.82,0.95] | <0.001 | 1.47 | [0.91,2.37] | 0.114 |
| W6 x 80+ | 1.31*** | [1.12,1.54] | <0.001 | 0.94 | [0.79,1.13] | 0.534 | 1.10 | [0.77,1.57] | 0.615 | 1.46 | [0.86,2.45] | 0.159 | 2.00*** | [1.57,2.55] | <0.001 | 0.88* | [0.78,0.98] | 0.023 | 1.19 | [0.69,2.03] | 0.532 |
| W11 x 60-69 | 1.23* | [1.03,1.47] | 0.022 | 1.00 | [0.79,1.26] | 0.997 | 1.17 | [0.81,1.70] | 0.395 | 1.26 | [0.70,2.27] | 0.446 | 1.35** | [1.11,1.64] | 0.003 | 0.75*** | [0.68,0.83] | <0.001 | 1.09 | [0.63,1.86] | 0.765 |
| W11 x 70-79 | 1.34*** | [1.13,1.59] | <0.001 | 1.01 | [0.81,1.26] | 0.923 | 1.00 | [0.69,1.43] | 0.981 | 1.51 | [0.84,2.71] | 0.168 | 1.58*** | [1.30,1.91] | <0.001 | 0.67*** | [0.60,0.75] | <0.001 | 1.44 | [0.85,2.43] | 0.170 |
| W11 x 80+ | 1.62*** | [1.35,1.95] | <0.001 | 0.90 | [0.70,1.14] | 0.379 | 1.26 | [0.83,1.89] | 0.275 | 1.97 | [1.00,3.87] | 0.051 | 2.96*** | [2.30,3.82] | <0.001 | 0.63*** | [0.53,0.75] | <0.001 | 1.23 | [0.69,2.19] | 0.494 |
| N | 21,311 | | | 17,213 | | | 21,360 | | | 14,271 | | | 21,256 | | | 14,473 | | | 21,364 | | |

| | Self-reported stroke | | | Self-reported other heart diseases | | | Obesity biomarker | | | Elevated C-reactive protein | | | Self-reported Arthritis | | |
|---|---|---|---|---|---|---|---|---|---|---|---|---|---|---|---|
| | RR | 95%CI | p | RR | 95%CI | p | RR | 95%CI | p | RR | 95%CI | p | RR | 95%CI | p |
| Wave (ref: w2) | | | <0.001 | | | 0.224 | | | 0.294 | | | <0.001 | | | <0.001 |
| Wave 6 | 4.97*** | [2.08,11.87] | <0.001 | 1.17 | [0.90,1.52] | 0.252 | 1.08 | [0.97,1.20] | 0.167 | 0.75*** | [0.66,0.86] | <0.001 | 1.06 | [0.93,1.20] | 0.388 |
| Wave 11 | 1.31** | [0.47,3.64] | 0.606 | 1.24 | [0.95,1.63] | 0.119 | 1.08 | [0.94,1.26] | 0.276 | 0.71*** | [0.58,0.87] | <0.001 | 0.63*** | [0.53,0.74] | <0.001 |
| Age (ref: 50-59) | | | <0.001 | | | <0.001 | | | <0.001 | | | <0.001 | | | <0.001 |
| 60-69 | 3.17** | [1.55,6.46] | 0.002 | 1.87*** | [1.58,2.21] | <0.001 | 0.95 | [0.87,1.04] | 0.307 | 1.10 | [1.00,1.20] | 0.053 | 1.36*** | [1.25,1.48] | <0.001 |
| 70-79 | 5.70*** | [2.82,11.37] | <0.001 | 2.55*** | [2.16,3.01] | <0.001 | 0.87** | [0.79,0.97] | 0.009 | 1.30*** | [1.19,1.43] | <0.001 | 1.52*** | [1.40,1.66] | <0.001 |
| 80+ | 9.48*** | [4.70,19.10] | <0.001 | 3.40*** | [2.85,4.06] | <0.001 | 0.56*** | [0.47,0.66] | <0.001 | 1.16* | [1.03,1.32] | 0.017 | 1.66*** | [1.51,1.82] | <0.001 |
| Wave (ref:w2) x Age (ref: 50-59) | | | 0.006 | | | 0.003 | | | 0.074 | | | 0.009 | | | <0.001 |
| W6 x 60-69 | 0.13*** | [0.05,0.37] | <0.001 | 0.69* | [0.51,0.94] | 0.020 | 1.10 | [0.96,1.27] | 0.171 | 1.12 | [0.95,1.32] | 0.179 | 1.02 | [0.88,1.18] | 0.820 |
| W6 x 70-79 | 0.16*** | [0.06,0.41] | <0.001 | 0.92 | [0.69,1.23] | 0.564 | 1.03 | [0.89,1.20] | 0.651 | 0.95 | [0.81,1.12] | 0.541 | 1.08 | [0.94,1.24] | 0.287 |
| W6 x 80+ | 0.19*** | [0.07,0.51] | <0.001 | 1.03 | [0.76,1.39] | 0.855 | 1.26* | [1.01,1.57] | 0.043 | 1.31** | [1.07,1.59] | 0.008 | 1.06 | [0.91,1.23] | 0.454 |
| W11 x 60-69 | 0.74 | [0.23,2.36] | 0.608 | 0.82 | [0.60,1.14] | 0.238 | 1.23* | [1.03,1.48] | 0.022 | 0.97 | [0.76,1.25] | 0.819 | 1.73*** | [1.43,2.09] | <0.001 |
| W11 x 70-79 | 0.51 | [0.17,1.55] | 0.233 | 0.86 | [0.63,1.16] | 0.323 | 1.19 | [0.99,1.44] | 0.071 | 0.87 | [0.68,1.11] | 0.272 | 2.07*** | [1.72,2.50] | <0.001 |
| W11 x 80+ | 0.71 | [0.23,2.17] | 0.541 | 1.12 | [0.82,1.52] | 0.468 | 1.08 | [0.82,1.43] | 0.565 | 0.83 | [0.62,1.13] | 0.237 | 1.97*** | [1.62,2.40] | <0.001 |
| N | 21,351 | | | 21,372 | | | 18,160 | | | 14,468 | | | | | |

***Panel C. Mental health and cognition***

| | Self-reported psychiatric problems | | | Depression | | | Depressive symptoms | | | CASP-19 | | | Memory | | |
|---|---|---|---|---|---|---|---|---|---|---|---|---|---|---|---|
| | RR | 95%CI | p | RR | 95%CI | p | Beta | 95%CI | p | Beta | 95%CI | p | Beta | 95%CI | p |
| Wave (ref: w2) | | | 0.216 | | | 0.016 | | | 0.100 | | | <0.001 | | | 0.155 |
| Wave 6 | 1.05 | [0.85,1.31] | 0.646 | 1.18* | [1.02,1.37] | 0.024 | 0.03 | [-0.10,0.17] | 0.633 | -2.43*** | [-3.09,-1.76] | <0.001 | 0.20 | [-0.01,0.41] | 0.066 |
| Wave 11 | 1.22 | [0.98,1.52] | 0.080 | 1.27* | [1.05,1.52] | 0.014 | 0.19* | [0.02,0.37] | 0.032 | -2.63*** | [-3.61,-1.66] | <0.001 | -0.02 | [-0.32,0.28] | 0.890 |
| Age (ref: 50-59) | | | <0.001 | | | 0.009 | | | <0.001 | | | <0.001 | | | <0.001 |
| 60-69 | 0.66*** | [0.55,0.80] | <0.001 | 0.90 | [0.78,1.02] | 0.108 | -0.11* | [-0.22,-0.01] | 0.034 | 0.04 | [-0.49,0.57] | 0.888 | -1.01*** | [-1.17,-0.84] | <0.001 |
| 70-79 | 0.37*** | [0.29,0.48] | <0.001 | 0.99 | [0.86,1.14] | 0.875 | -0.01 | [-0.13,0.11] | 0.890 | -1.58*** | [-2.19,-0.96] | <0.001 | -2.27*** | [-2.46,-2.08] | <0.001 |
| 80+ | 0.37*** | [0.27,0.52] | <0.001 | 1.17* | [1.00,1.37] | 0.045 | 0.32*** | [0.16,0.47] | <0.001 | -5.03*** | [-5.93,-4.13] | <0.001 | -4.32*** | [-4.57,-4.06] | <0.001 |
| Wave (ref:w2) x Age (ref: 50-59) | | | 0.004 | | | <0.001 | | | 0.001 | | | <0.001 | | | <0.001 |
| W6 x 60-69 | 1.25 | [0.92,1.69] | 0.155 | 0.79* | [0.65,0.98] | 0.029 | -0.15 | [-0.32,0.03] | 0.099 | 0.66 | [-0.21,1.53] | 0.139 | 0.60*** | [0.33,0.87] | <0.001 |
| W6 x 70-79 | 1.32 | [0.92,1.90] | 0.132 | 0.72** | [0.59,0.89] | 0.002 | -0.24** | [-0.42,-0.06] | 0.009 | 1.12* | [0.22,2.03] | 0.015 | 0.28 | [-0.00,0.57] | 0.053 |
| W6 x 80+ | 0.71 | [0.42,1.20] | 0.199 | 0.78* | [0.62,0.98] | 0.031 | -0.21 | [-0.43,0.01] | 0.067 | 1.25* | [0.01,2.49] | 0.049 | 0.26 | [-0.12,0.65] | 0.175 |
| W11 x 60-69 | 1.66*** | [1.23,2.23] | <0.001 | 1.12 | [0.88,1.42] | 0.359 | 0.08 | [-0.14,0.30] | 0.485 | -0.44 | [-1.64,0.75] | 0.466 | 0.47* | [0.10,0.84] | 0.012 |
| W11 x 70-79 | 1.83** | [1.27,2.65] | 0.001 | 0.87 | [0.68,1.12] | 0.281 | -0.13 | [-0.36,0.09] | 0.237 | 1.55* | [0.33,2.78] | 0.013 | 1.22*** | [0.84,1.59] | <0.001 |
| W11 x 80+ | 0.86 | [0.52,1.44] | 0.568 | 0.70* | [0.53,0.92] | 0.012 | -0.38** | [-0.63,-0.12] | 0.004 | 2.52*** | [1.07,3.97] | <0.001 | 1.15*** | [0.71,1.59] | <0.001 |
| N | 21,350 | | | 21,992 | | | 21,992 | | | 20,055 | | | 21,929 | | |

Data source: English Longitudinal Study of Ageing waves 2, 6 and 11. Notes: RR = Rate ratio; CI: confidence interval. P-values for overall wave, age group, and wave x age group correspond to the omnibus test. The outcome "other heart diseases" was established if the respondent reported one of the following: heart failure, heart murmur, heart attack, abnormal heart rhythm, or angina. All analyses also controlled for gender, education, and wealth. Depending on health outcomes, different sampling weights were applied (cross-sectional weight, self-completion weight, nurse visit weight, or blood sample weight). P values: ***<0.001; **<0.01; *<0.05.

#### Table S5. Weighted results from Poisson or linear regression models testing the statistical effect of time (wave), education, and their interaction on health outcomes

***Panel A. Self-rated health, pain, and physical functioning***

| | Poor/fair self-rated health | | | Limiting long-standing illness | | | Any pain | | | Moderate/severe pain | | | Any ADL limitations | | | Any IADL limitations | | | Any mobility impairments | | |
|---|---|---|---|---|---|---|---|---|---|---|---|---|---|---|---|---|---|---|---|---|---|
| | RR | 95%CI | p | RR | 95%CI | p | RR | 95%CI | p | RR | 95%CI | p | RR | 95%CI | p | RR | 95%CI | p | RR | 95%CI | p |
| Wave (ref:w2) | | | 0.180 | | | 0.034 | | | <0.001 | | | 0.163 | | | 0.146 | | | 0.003 | | | <0.001 |
| Wave 6 | 1.01 | [0.83,1.22] | 0.950 | 0.85* | [0.74,0.98] | 0.021 | 1.19** | [1.04,1.35] | 0.010 | 1.19 | [0.99,1.42] | 0.061 | 0.80 | [0.64,1.01] | 0.062 | 0.72** | [0.58,0.90] | 0.004 | 0.87** | [0.79,0.96] | 0.006 |
| Wave 11 | 1.16 | [0.96,1.40] | 0.127 | 0.85* | [0.75,0.98] | 0.025 | 1.33*** | [1.16,1.52] | <0.001 | 1.09 | [0.91,1.31] | 0.350 | 0.83 | [0.66,1.05] | 0.117 | 0.71** | [0.57,0.88] | 0.002 | 0.77*** | [0.70,0.86] | <0.001 |
| Edu (ref:High) | | | <0.001 | | | 0.066 | | | <0.001 | | | <0.001 | | | <0.001 | | | 0.008 | | | <0.001 |
| Medium | 1.34*** | [1.14,1.58] | <0.001 | 1.07 | [0.95,1.20] | 0.250 | 1.31*** | [1.17,1.47] | <0.001 | 1.43*** | [1.22,1.67] | <0.001 | 1.35** | [1.11,1.63] | 0.002 | 1.22* | [1.02,1.45] | 0.028 | 1.19*** | [1.10,1.29] | <0.001 |
| Low | 1.79*** | [1.52,2.11] | <0.001 | 1.13* | [1.00,1.27] | 0.042 | 1.42*** | [1.26,1.60] | <0.001 | 1.62*** | [1.37,1.90] | <0.001 | 1.46*** | [1.20,1.76] | <0.001 | 1.31** | [1.09,1.56] | 0.003 | 1.21*** | [1.12,1.31] | <0.001 |
| Wave (ref:w2) x Edu (ref:High) | | | 0.385 | | | <0.001 | | | 0.135 | | | 0.004 | | | 0.174 | | | <0.001 | | | <0.001 |
| W6 x Medium | 0.99 | [0.81,1.22] | 0.949 | 1.14 | [0.99,1.33] | 0.075 | 0.95 | [0.83,1.10] | 0.521 | 0.90 | [0.74,1.09] | 0.289 | 1.07 | [0.83,1.37] | 0.616 | 1.16 | [0.92,1.48] | 0.213 | 1.01 | [0.92,1.12] | 0.777 |
| W6 x Low | 1.08 | [0.88,1.32] | 0.449 | 1.27** | [1.09,1.47] | 0.002 | 0.95 | [0.83,1.10] | 0.513 | 0.95 | [0.78,1.15] | 0.574 | 1.19 | [0.93,1.53] | 0.161 | 1.33* | [1.05,1.69] | 0.017 | 1.11* | [1.00,1.23] | 0.047 |
| W11 xMedium | 1.08 | [0.88,1.33] | 0.442 | 1.25** | [1.08,1.46] | 0.004 | 0.92 | [0.79,1.06] | 0.247 | 1.07 | [0.88,1.31] | 0.509 | 1.15 | [0.90,1.48] | 0.270 | 1.29* | [1.02,1.64] | 0.032 | 1.14* | [1.01,1.27] | 0.027 |
| W11 x Low | 1.12 | [0.91,1.39] | 0.279 | 1.34*** | [1.13,1.59] | <0.001 | 1.03 | [0.89,1.21] | 0.665 | 1.30* | [1.05,1.60] | 0.014 | 1.31* | [1.00,1.71] | 0.050 | 1.63*** | [1.27,2.09] | <0.001 | 1.33*** | [1.18,1.50] | <0.001 |
| N | 22,115 | | | 22,120 | | | 22,124 | | | 22,117 | | | 22,125 | | | 22,125 | | | 22,126 | | |

***Panel B. Chronic conditions, obesity, and inflammation***

| | Self-reported hypertension | | | Hypertension biomarker | | | Self-reported diabetes | | | Diabetes biomarkers | | | Self-reported high cholesterol | | | High cholesterol biomarker | | | Self-reported lung disease | | |
|---|---|---|---|---|---|---|---|---|---|---|---|---|---|---|---|---|---|---|---|---|---|
| | RR | 95%CI | p | RR | 95%CI | p | RR | 95%CI | p | RR | 95%CI | p | RR | 95%CI | p | RR | 95%CI | p | RR | 95%CI | p |
| Wave (ref:w2) | | | 0.680 | | | <0.001 | | | 0.965 | | | 0.213 | | | <0.001 | | | <0.001 | | | 0.182 |
| Wave 6 | 0.99 | [0.88,1.10] | 0.797 | 0.80** | [0.70,0.93] | 0.003 | 1.03 | [0.80,1.33] | 0.795 | 1.37 | [0.92,2.04] | 0.122 | 1.53*** | [1.31,1.77] | <0.001 | 0.89*** | [0.84,0.94] | <0.001 | 0.82 | [0.50,1.34] | 0.431 |
| Wave 11 | 1.03 | [0.91,1.17] | 0.606 | 0.75*** | [0.64,0.88] | <0.001 | 1.01 | [0.76,1.35] | 0.921 | 1.08 | [0.70,1.67] | 0.726 | 1.87*** | [1.59,2.19] | <0.001 | 0.79*** | [0.74,0.85] | <0.001 | 1.24 | [0.74,2.07] | 0.418 |
| Edu (ref:High) | | | 0.133 | | | 0.075 | | | 0.620 | | | 0.966 | | | 0.706 | | | 0.270 | | | 0.006 |
| Medium | 1.10 | [0.99,1.23] | 0.079 | 1.07 | [0.96,1.20] | 0.214 | 0.91 | [0.71,1.16] | 0.433 | 1.05 | [0.72,1.52] | 0.799 | 1.07 | [0.91,1.25] | 0.411 | 0.97 | [0.93,1.01] | 0.119 | 1.83** | [1.16,2.87] | 0.009 |
| Low | 1.12* | [1.00,1.26] | 0.045 | 1.14* | [1.01,1.28] | 0.035 | 0.96 | [0.75,1.24] | 0.767 | 1.05 | [0.72,1.54] | 0.802 | 1.06 | [0.91,1.25] | 0.453 | 0.98 | [0.93,1.03] | 0.410 | 2.09** | [1.32,3.30] | 0.002 |
| Wave (ref:w2) x Edu (ref:High) | | | 0.055 | | | 0.951 | | | 0.017 | | | 0.401 | | | 0.224 | | | <0.001 | | | 0.684 |
| W6 x Medium | 1.03 | [0.91,1.16] | 0.688 | 1.03 | [0.88,1.21] | 0.719 | 1.31 | [0.99,1.73] | 0.058 | 1.16 | [0.75,1.80] | 0.501 | 1.06 | [0.89,1.25] | 0.514 | 0.99 | [0.93,1.05] | 0.733 | 1.14 | [0.68,1.92] | 0.615 |
| W6 x Low | 1.11 | [0.98,1.26] | 0.102 | 1.04 | [0.88,1.24] | 0.609 | 1.48** | [1.12,1.97] | 0.006 | 1.29 | [0.83,2.02] | 0.255 | 1.16 | [0.97,1.37] | 0.096 | 0.91* | [0.84,0.98] | 0.018 | 1.27 | [0.75,2.13] | 0.375 |
| W11 xMedium | 1.09 | [0.94,1.25] | 0.258 | 1.06 | [0.88,1.28] | 0.513 | 1.56** | [1.13,2.17] | 0.007 | 1.41 | [0.86,2.30] | 0.168 | 1.05 | [0.88,1.26] | 0.564 | 0.88** | [0.80,0.96] | 0.003 | 0.92 | [0.53,1.62] | 0.782 |
| W11 x Low | 1.23** | [1.05,1.44] | 0.010 | 1.02 | [0.81,1.29] | 0.851 | 1.73** | [1.21,2.47] | 0.003 | 1.76 | [0.99,3.14] | 0.056 | 1.21 | [1.00,1.48] | 0.055 | 0.73*** | [0.62,0.87] | <0.001 | 1.15 | [0.63,2.09] | 0.646 |
| N | 21,311 | | | 17,213 | | | 21,360 | | | 14,271 | | | 21,256 | | | 14,473 | | | 21,364 | | |

| | Self-reported stroke | | | Self-reported other heart diseases | | | Obesity biomarker | | | Elevated C-reactive protein | | | Self-reported Arthritis | | |
|---|---|---|---|---|---|---|---|---|---|---|---|---|---|---|---|
| | RR | 95%CI | p | RR | 95%CI | p | RR | 95%CI | p | RR | 95%CI | p | RR | 95%CI | p |
| Wave (ref:w2) | | | 0.190 | | | 0.540 | | | 0.055 | | | 0.008 | | | 0.015 |
| Wave 6 | 1.51 | [0.68,3.37] | 0.312 | 1.11 | [0.92,1.33] | 0.285 | 1.14 | [0.96,1.34] | 0.130 | 0.91 | [0.75,1.10] | 0.310 | 1.18** | [1.05,1.33] | 0.006 |
| Wave 11 | 0.80 | [0.34,1.85] | 0.598 | 1.05 | [0.85,1.30] | 0.650 | 1.24* | [1.04,1.47] | 0.016 | 0.71** | [0.57,0.88] | 0.002 | 1.08 | [0.94,1.23] | 0.269 |
| Edu (ref:High) | | | 0.549 | | | 0.200 | | | <0.001 | | | <0.001 | | | <0.001 |
| Medium | 1.22 | [0.60,2.48] | 0.585 | 1.05 | [0.87,1.27] | 0.616 | 1.23** | [1.06,1.43] | 0.005 | 1.34*** | [1.15,1.55] | <0.001 | 1.25*** | [1.11,1.41] | <0.001 |
| Low | 1.41 | [0.69,2.88] | 0.349 | 0.95 | [0.78,1.15] | 0.578 | 1.49*** | [1.28,1.73] | <0.001 | 1.49*** | [1.27,1.73] | <0.001 | 1.26*** | [1.12,1.43] | <0.001 |
| Wave (ref:w2) x Edu (ref:High) | | | 0.610 | | | 0.245 | | | 0.980 | | | 0.426 | | | 0.207 |
| W6 x Medium | 0.65 | [0.27,1.59] | 0.349 | 0.89 | [0.72,1.10] | 0.277 | 1.02 | [0.86,1.22] | 0.798 | 0.84 | [0.68,1.03] | 0.091 | 0.88 | [0.78,1.00] | 0.055 |
| W6 x Low | 0.65 | [0.27,1.56] | 0.334 | 0.95 | [0.77,1.18] | 0.639 | 1.00 | [0.83,1.20] | 0.984 | 0.91 | [0.74,1.12] | 0.379 | 0.94 | [0.82,1.06] | 0.309 |
| W11 xMedium | 1.21 | [0.48,3.07] | 0.681 | 1.09 | [0.86,1.39] | 0.475 | 1.00 | [0.82,1.21] | 0.969 | 0.91 | [0.71,1.16] | 0.444 | 0.96 | [0.83,1.12] | 0.599 |
| W11 x Low | 0.98 | [0.35,2.72] | 0.970 | 1.20 | [0.91,1.60] | 0.196 | 0.96 | [0.76,1.21] | 0.701 | 0.96 | [0.71,1.31] | 0.817 | 1.02 | [0.86,1.20] | 0.837 |
| N | 21,351 | | | 21,372 | | | 18,160 | | | 14,468 | | | | | |

***Panel C. Mental health and cognition***

| | Self-reported psychiatric problems | | | Depression | | | Depressive symptoms | | | CASP-19 | | | Memory | | |
|---|---|---|---|---|---|---|---|---|---|---|---|---|---|---|---|
| | RR | 95%CI | p | RR | 95%CI | p | Beta | 95%CI | p | Beta | 95%CI | p | Beta | 95%CI | p |
| Wave (ref:w2) | | | 0.414 | | | 0.429 | | | 0.013 | | | <0.001 | | | <0.001 |
| Wave 6 | 1.14 | [0.85,1.52] | 0.391 | 0.90 | [0.69,1.18] | 0.464 | -0.17* | [-0.29,-0.04] | 0.011 | -1.58*** | [-2.21,-0.94] | <0.001 | 0.66*** | [0.42,0.89] | <0.001 |
| Wave 11 | 1.22 | [0.91,1.64] | 0.187 | 1.06 | [0.82,1.38] | 0.643 | -0.01 | [-0.14,0.13] | 0.927 | -1.38*** | [-2.12,-0.63] | <0.001 | 0.74*** | [0.45,1.03] | <0.001 |
| Edu (ref:High) | | | 0.090 | | | <0.001 | | | <0.001 | | | <0.001 | | | <0.001 |
| Medium | 0.89 | [0.68,1.16] | 0.386 | 1.23 | [0.98,1.53] | 0.072 | 0.16** | [0.04,0.28] | 0.008 | -0.90** | [-1.49,-0.31] | <0.001 | -0.77*** | [-0.99,-0.56] | <0.001 |
| Low | 0.75 | [0.57,1.01] | 0.055 | 1.59*** | [1.27,1.99] | <0.001 | 0.45*** | [0.32,0.58] | <0.001 | -3.28*** | [-3.97,-2.59] | <0.001 | -2.14*** | [-2.38,-1.91] | <0.001 |
| Wave (ref:w2) x Edu (ref:High) | | | 0.228 | | | 0.689 | | | 0.139 | | | 0.037 | | | 0.395 |
| W6 x Medium | 0.96 | [0.69,1.33] | 0.808 | 1.06 | [0.79,1.43] | 0.682 | 0.05 | [-0.11,0.20] | 0.554 | -0.61 | [-1.36,0.15] | 0.344 | -0.13 | [-0.40,0.14] | 0.344 |
| W6 x Low | 1.04 | [0.73,1.47] | 0.839 | 1.15 | [0.86,1.54] | 0.339 | 0.13 | [-0.05,0.30] | 0.160 | 0.37 | [-0.52,1.26] | 0.066 | -0.29 | [-0.60,0.02] | 0.066 |
| W11 xMedium | 1.27 | [0.91,1.79] | 0.166 | 1.09 | [0.81,1.45] | 0.577 | 0.10 | [-0.07,0.27] | 0.244 | -1.01* | [-1.94,-0.07] | 0.281 | -0.19 | [-0.53,0.15] | 0.281 |
| W11 x Low | 1.55* | [1.01,2.36] | 0.044 | 1.20 | [0.88,1.63] | 0.250 | 0.35* | [0.08,0.63] | 0.012 | -0.68 | [-2.14,0.78] | 0.444 | -0.17 | [-0.62,0.27] | 0.444 |
| N | 21,350 | | | 21,992 | | | 21,992 | | | 20,055 | | | 21,929 | | |

Data source: English Longitudinal Study of Ageing waves 2, 6 and 11. Notes: RR = Rate ratio; CI: confidence interval. P-values for overall wave, education, and wave x education correspond to the omnibus test. The outcome "other heart diseases" was established if the respondent reported one of the following: heart failure, heart murmur, heart attack, abnormal heart rhythm, or angina. All analyses also controlled for gender, age group, and wealth. Depending on health outcomes, different sampling weights were applied (cross-sectional weight, self-completion weight, nurse visit weight, or blood sample weight). P values: ***<0.001; **<0.01; *<0.05.

#### Table S6. Overall adjusted predicted probabilities and absolute percentage-point changes in health outcomes, 2004/05 to 2023/24

| Health outcomes | Predicted Probability 2004–05 | Predicted Probability 2023–24 | Absolute change [95% CI] | p-values for trend |
|---|---|---|---|---|
| ***Panel A. Self-rated health, pain, and functional limitations*** | | | | |
| Poor/fair self-rated health | 0.27 [0.26, 0.28] | 0.34 [0.33, 0.36] | 0.07 [0.05, 0.09] | <0.001 |
| Limiting long-standing illness | 0.35 [0.34, 0.36] | 0.37 [0.36, 0.39] | 0.03 [0.01, 0.04] | 0.010 |
| Any pain | 0.37 [0.36, 0.38] | 0.47 [0.46, 0.49] | 0.10 [0.08, 0.12] | <0.001 |
| Moderate/severe pain | 0.27 [0.26, 0.28] | 0.33 [0.31, 0.34] | 0.06 [0.04, 0.08] | <0.001 |
| Any ADL limitations | 0.20 [0.19, 0.21] | 0.20 [0.19, 0.21] | -0.00 [-0.02, 0.02] | 0.917 |
| Any IADL limitations | 0.22 [0.21, 0.23] | 0.21 [0.20, 0.23] | -0.00 [-0.02, 0.01] | 0.611 |
| Any mobility impairments | 0.58 [0.57, 0.59] | 0.52 [0.50, 0.54] | -0.06 [-0.08, -0.04] | <0.001 |
| ***Panel B. Chronic conditions, obesity, and inflammation*** | | | | |
| Self-reported hypertension | 0.35 [0.34, 0.36] | 0.40 [0.39, 0.42] | 0.05 [0.03, 0.07] | <0.001 |
| Hypertension biomarker | 0.38 [0.36, 0.39] | 0.30 [0.28, 0.31] | -0.08 [-0.10, -0.06] | <0.001 |
| Self-reported diabetes | 0.08 [0.08, 0.09] | 0.13 [0.12, 0.14] | 0.05 [0.03, 0.06] | <0.001 |
| Diabetes biomarker | 0.07 [0.06, 0.07] | 0.11 [0.09, 0.12] | 0.04 [0.02, 0.06] | <0.001 |
| Self-reported high cholesterol | 0.19 [0.18, 0.20] | 0.39 [0.37, 0.41] | 0.20 [0.18, 0.22] | <0.001 |
| High cholesterol biomarker | 0.78 [0.77, 0.79] | 0.54 [0.52, 0.56] | -0.24 [-0.26, -0.21] | <0.001 |
| Self-reported lung disease | 0.05 [0.05, 0.06] | 0.06 [0.05, 0.07] | 0.01 [0.00, 0.02] | 0.014 |
| Self-reported stroke | 0.02 [0.01, 0.02] | 0.01 [0.01, 0.02] | -0.00 [-0.01, 0.00] | 0.347 |
| Self-reported heart diseases | 0.15 [0.14, 0.16] | 0.18 [0.16, 0.19] | 0.02 [0.01, 0.04] | 0.002 |
| Obesity biomarker | 0.28 [0.27, 0.29] | 0.34 [0.32, 0.36] | 0.06 [0.04, 0.09] | <0.001 |
| Elevated C-reactive protein | 0.37 [0.35, 0.38] | 0.24 [0.22, 0.26] | -0.12 [-0.15, -0.10] | <0.001 |
| Self-reported arthritis | 0.36 [0.35, 0.37] | 0.38 [0.36, 0.39] | 0.02 [0.00, 0.04] | 0.027 |
| ***Panel C. Mental health and cognition*** | | | | |
| Self-reported psychiatric problems | 0.06 [0.06, 0.07] | 0.10 [0.09, 0.11] | 0.04 [0.02, 0.05] | <0.001 |
| Depression | 0.15 [0.14, 0.16] | 0.18 [0.17, 0.20] | 0.03 [0.01, 0.05] | <0.001 |
| Depressive symptoms | 1.56 [1.51, 1.60] | 1.69 [1.62, 1.76] | 0.13 [0.05, 0.21] | 0.002 |
| CASP-19 | 41.14 [40.92, 41.38] | 39.12 [38.72, 39.51] | -2.04 [-2.49, -1.58] | <0.001 |
| Memory | 9.99 [9.92, 10.06] | 10.57 [10.45, 10.69] | 0.58 [0.44, 0.72] | <0.001 |

Data source: English Longitudinal Study of Ageing (ELSA) waves 2 (2004–05), 6 (2012–13) and 11 (2023–24). Notes: Estimates are adjusted predicted probabilities from regression models controlling for age groups, sex, education, and wealth quintiles. Absolute change is expressed in percentage points and calculated as the adjusted predicted probability in 2023–24 minus that in 2004–05. Confidence intervals for the change test whether the adjusted predicted probabilities differ between the two survey periods (with relevant p-value).

#### Table S7. Within-age-group adjusted predicted probabilities and absolute percentage-point changes in health outcomes, 2004/05 to 2023/24

| Health outcomes | Predicted Probability 2004–05 | Predicted Probability 2023–24 | Absolute change [95% CI] | p-values for trend |
|---|---|---|---|---|
| ***Panel A. Self-rated health, pain, and functional limitations*** | | | | |
| Poor/fair self-rated health | | | | |
| 50–59 | 0.23 [0.21, 0.25] | 0.29 [0.25, 0.32] | 0.05 [0.02, 0.09] | 0.004 |
| 60–69 | 0.27 [0.25, 0.28] | 0.32 [0.29, 0.34] | 0.05 [0.02, 0.08] | 0.002 |
| 70–79 | 0.30 [0.28, 0.32] | 0.39 [0.37, 0.42] | 0.09 [0.05, 0.13] | <0.001 |
| 80+ | 0.33 [0.31, 0.36] | 0.46 [0.42, 0.49] | 0.12 [0.07, 0.17] | <0.001 |
| Limiting long-standing illness | | | | |
| 50–59 | 0.28 [0.26, 0.30] | 0.29 [0.26, 0.32] | 0.01 [-0.03, 0.04] | 0.775 |
| 60–69 | 0.34 [0.32, 0.35] | 0.35 [0.32, 0.37] | 0.01 [-0.02, 0.04] | 0.570 |
| 70–79 | 0.39 [0.37, 0.42] | 0.43 [0.40, 0.46] | 0.03 [-0.00, 0.07] | 0.051 |
| 80+ | 0.46 [0.43, 0.49] | 0.55 [0.51, 0.58] | 0.08 [0.04, 0.13] | 0.001 |
| Any pain | | | | |
| 50–59 | 0.35 [0.33, 0.37] | 0.44 [0.40, 0.48] | 0.09 [0.05, 0.13] | <0.001 |
| 60–69 | 0.39 [0.37, 0.40] | 0.47 [0.44, 0.50] | 0.09 [0.05, 0.12] | <0.001 |
| 70–79 | 0.39 [0.36, 0.41] | 0.50 [0.47, 0.53] | 0.11 [0.08, 0.15] | <0.001 |
| 80+ | 0.38 [0.35, 0.41] | 0.51 [0.48, 0.55] | 0.13 [0.09, 0.18] | <0.001 |
| Moderate/severe pain | | | | |
| 50–59 | 0.23 [0.22, 0.25] | 0.26 [0.23, 0.30] | 0.03 [-0.01, 0.07] | 0.114 |
| 60–69 | 0.28 [0.27, 0.30] | 0.34 [0.31, 0.36] | 0.05 [0.02, 0.09] | 0.002 |
| 70–79 | 0.29 [0.27, 0.31] | 0.37 [0.34, 0.40] | 0.08 [0.05, 0.11] | <0.001 |
| 80+ | 0.28 [0.25, 0.30] | 0.39 [0.35, 0.42] | 0.11 [0.07, 0.15] | <0.001 |
| Any ADL limitations | | | | |
| 50–59 | 0.13 [0.12, 0.14] | 0.13 [0.11, 0.16] | 0.01 [-0.03, 0.03] | 0.839 |
| 60–69 | 0.18 [0.17, 0.20] | 0.19 [0.16, 0.21] | 0.01 [-0.02, 0.03] | 0.673 |
| 70–79 | 0.25 [0.23, 0.27] | 0.23 [0.20, 0.25] | -0.02 [-0.05, 0.01] | 0.192 |
| 80+ | 0.33 [0.30, 0.36] | 0.34 [0.30, 0.38] | 0.01 [-0.04, 0.05] | 0.734 |
| Any IADL limitations | | | | |
| 50–59 | 0.15 [0.14, 0.16] | 0.15 [0.12, 0.18] | -0.00 [-0.03, 0.03] | 0.925 |
| 60–69 | 0.18 [0.17, 0.20] | 0.18 [0.16, 0.21] | -0.00 [-0.03, 0.03] | 0.875 |
| 70–79 | 0.24 [0.22, 0.26] | 0.23 [0.20, 0.25] | -0.02 [-0.05, 0.01] | 0.281 |
| 80+ | 0.42 [0.39, 0.45] | 0.42 [0.38, 0.46] | 0.00 [-0.05, 0.05] | 0.991 |
| Any mobility impairments | | | | |
| 50–59 | 0.46 [0.44, 0.48] | 0.37 [0.33, 0.40] | -0.09 [-0.13, -0.05] | <0.001 |
| 60–69 | 0.57 [0.55, 0.58] | 0.50 [0.48, 0.53] | -0.06 [-0.09, -0.03] | <0.001 |
| 70–79 | 0.66 [0.64, 0.68] | 0.62 [0.59, 0.64] | -0.04 [-0.07, -0.01] | 0.011 |
| 80+ | 0.75 [0.72, 0.77] | 0.75 [0.72, 0.78] | 0.01 [-0.03, 0.05] | 0.712 |
| ***Panel B. Chronic conditions, obesity, and inflammation*** | | | | |
| Self-reported hypertension | | | | |
| 50–59 | 0.26 [0.25, 0.28] | 0.24 [0.21, 0.27] | -0.02 [-0.06, 0.01] | 0.196 |
| 60–69 | 0.34 [0.33, 0.36] | 0.38 [0.35, 0.41] | 0.04 [0.01, 0.07] | 0.024 |
| 70–79 | 0.43 [0.41, 0.45] | 0.52 [0.49, 0.55] | 0.09 [0.06, 0.13] | <0.001 |
| 80+ | 0.39 [0.36, 0.42] | 0.58 [0.54, 0.61] | 0.19 [0.14, 0.23] | <0.001 |

| | | | | |
|---|---|---|---|---|
| Hypertension biomarker | | | | |
| 50–59 | 0.29 [0.26, 0.31] | 0.23 [0.19, 0.27] | -0.06 [-0.10, -0.01] | 0.015 |
| 60–69 | 0.37 [0.35, 0.39] | 0.29 [0.26, 0.33] | -0.07 [-0.11, -0.04] | <0.001 |
| 70–79 | 0.46 [0.43, 0.48] | 0.37 [0.34, 0.40] | -0.09 [-0.13, -0.05] | <0.001 |
| 80+ | 0.48 [0.44, 0.52] | 0.34 [0.30, 0.39] | -0.14 [-0.19, -0.08] | <0.001 |
| Self-reported diabetes | | | | |
| 50–59 | 0.05 [0.05, 0.06] | 0.08 [0.06, 0.10] | 0.02 [0.00, 0.05] | 0.043 |
| 60–69 | 0.08 [0.07, 0.10] | 0.14 [0.12, 0.16] | 0.06 [0.03, 0.08] | <0.001 |
| 70–79 | 0.12 [0.10, 0.13] | 0.17 [0.15, 0.19] | 0.05 [0.02, 0.08] | <0.001 |
| 80+ | 0.09 [0.07, 0.11] | 0.17 [0.14, 0.20] | 0.07 [0.04, 0.11] | <0.001 |
| Diabetes biomarker | | | | |
| 50–59 | 0.04 [0.03, 0.05] | 0.05 [0.03, 0.07] | 0.01 [-0.02, 0.03] | 0.610 |
| 60–69 | 0.08 [0.07, 0.10] | 0.12 [0.09, 0.15] | 0.04 [0.01, 0.07] | 0.018 |
| 70–79 | 0.08 [0.07, 0.10] | 0.15 [0.12, 0.17] | 0.06 [0.03, 0.09] | <0.001 |
| 80+ | 0.07 [0.04, 0.09] | 0.15 [0.11, 0.19] | 0.08 [0.04, 0.13] | <0.001 |
| Self-reported high cholesterol | | | | |
| 50–59 | 0.16 [0.15, 0.18] | 0.23 [0.20, 0.26] | 0.07 [0.03, 0.10] | <0.001 |
| 60–69 | 0.22 [0.20, 0.23] | 0.41 [0.38, 0.44] | 0.19 [0.16, 0.23] | <0.001 |
| 70–79 | 0.23 [0.21, 0.25] | 0.51 [0.48, 0.54] | 0.28 [0.25, 0.31] | <0.001 |
| 80+ | 0.11 [0.09, 0.13] | 0.46 [0.42, 0.49] | 0.35 [0.31, 0.39] | <0.001 |
| High cholesterol biomarker | | | | |
| 50–59 | 0.84 [0.83, 0.86] | 0.73 [0.68, 0.77] | -0.12 [-0.17, -0.07] | <0.001 |
| 60–69 | 0.81 [0.79, 0.82] | 0.52 [0.48, 0.56] | -0.29 [-0.33, -0.24] | <0.001 |
| 70–79 | 0.72 [0.70, 0.75] | 0.42 [0.38, 0.45] | -0.31 [-0.35, -0.26] | <0.001 |
| 80+ | 0.66 [0.62, 0.70] | 0.36 [0.30, 0.41] | -0.30 [-0.37, -0.23] | <0.001 |
| Self-reported lung disease | | | | |
| 50–59 | 0.03 [0.03, 0.04] | 0.04 [0.02, 0.05] | 0.00 [-0.01, 0.02] | 0.824 |
| 60–69 | 0.05 [0.05, 0.06] | 0.06 [0.05, 0.08] | 0.01 [-0.01, 0.03] | 0.401 |
| 70–79 | 0.06 [0.05, 0.07] | 0.09 [0.07, 0.11] | 0.03 [0.01, 0.05] | 0.003 |
| 80+ | 0.06 [0.04, 0.07] | 0.08 [0.05, 0.10] | 0.02 [-0.01, 0.04] | 0.189 |
| Self-reported stroke | | | | |
| 50–59 | 0.00 [0.00, 0.01] | 0.01 [0.00, 0.01] | 0.00 [-0.00, 0.01] | 0.622 |
| 60–69 | 0.01 [0.01, 0.02] | 0.01 [0.01, 0.02] | -0.00 [-0.01, 0.01] | 0.902 |
| 70–79 | 0.02 [0.02, 0.03] | 0.02 [0.01, 0.02] | -0.01 [-0.02, 0.00] | 0.085 |
| 80+ | 0.04 [0.03, 0.05] | 0.04 [0.02, 0.05] | -0.00 [-0.02, 0.02] | 0.745 |
| Self-reported heart diseases | | | | |
| 50–59 | 0.08 [0.07, 0.09] | 0.09 [0.07, 0.12] | 0.02 [-0.01, 0.04] | 0.142 |
| 60–69 | 0.14 [0.13, 0.16] | 0.15 [0.13, 0.17] | 0.00 [-0.02, 0.03] | 0.788 |
| 70–79 | 0.19 [0.18, 0.21] | 0.21 [0.19, 0.23] | 0.01 [-0.02, 0.04] | 0.394 |
| 80+ | 0.26 [0.23, 0.29] | 0.36 [0.33, 0.40] | 0.10 [0.06, 0.15] | <0.001 |
| Obesity biomarker | | | | |
| 50–59 | 0.31 [0.29, 0.33] | 0.34 [0.30, 0.38] | 0.03 [-0.02, 0.08] | 0.289 |
| 60–69 | 0.30 [0.28, 0.32] | 0.40 [0.37, 0.43] | 0.10 [0.06, 0.14] | <0.001 |
| 70–79 | 0.27 [0.25, 0.29] | 0.35 [0.32, 0.38] | 0.08 [0.04, 0.12] | <0.001 |
| 80+ | 0.17 [0.15, 0.20] | 0.21 [0.17, 0.24] | 0.03 [-0.01, 0.08] | 0.187 |
| Elevated C-reactive protein | | | | |
| 50–59 | 0.33 [0.30, 0.35] | 0.23 [0.19, 0.28] | -0.09 [-0.14, -0.05] | <0.001 |
| 60–69 | 0.36 [0.34, 0.38] | 0.25 [0.21, 0.28] | -0.11 [-0.15, -0.07] | <0.001 |
| 70–79 | 0.43 [0.40, 0.45] | 0.27 [0.23, 0.30] | -0.16 [-0.20, -0.12] | <0.001 |
| 80+ | 0.38 [0.34, 0.42] | 0.23 [0.18, 0.27] | -0.15 [-0.21, -0.09] | <0.001 |

| | | | | |
|---|---|---|---|---|
| Self-reported arthritis | | | | |
| 50–59 | 0.26 [0.25, 0.28] | 0.17 [0.14, 0.19] | -0.10 [-0.13, -0.07] | <0.001 |
| 60–69 | 0.36 [0.34, 0.38] | 0.39 [0.36, 0.42] | 0.03 [-0.00, 0.06] | 0.086 |
| 70–79 | 0.40 [0.38, 0.42] | 0.52 [0.50, 0.55] | 0.12 [0.09, 0.15] | <0.001 |
| 80+ | 0.44 [0.35, 0.37] | 0.54 [0.50, 0.58] | 0.10 [0.06, 0.15] | <0.001 |
| ***Panel C. Mental health and cognition*** | | | | |
| Self-reported psychiatric problems | | | | |
| 50–59 | 0.10 [0.09, 0.11] | 0.12 [0.10, 0.14] | 0.02 [-0.00, 0.05] | 0.096 |
| 60–69 | 0.07 [0.06, 0.08] | 0.13 [0.11, 0.15] | 0.07 [0.04, 0.09] | <0.001 |
| 70–79 | 0.04 [0.03, 0.05] | 0.08 [0.07, 0.10] | 0.05 [0.03, 0.06] | <0.001 |
| 80+ | 0.04 [0.03, 0.05] | 0.04 [0.03, 0.05] | 0.00 [-0.02, 0.02] | 0.836 |
| Depression | | | | |
| 50–59 | 0.15 [0.14, 0.17] | 0.19 [0.16, 0.23] | 0.04 [0.01, 0.07] | 0.021 |
| 60–69 | 0.14 [0.12, 0.15] | 0.19 [0.17, 0.22] | 0.06 [0.03, 0.09] | <0.001 |
| 70–79 | 0.15 [0.14, 0.17] | 0.17 [0.15, 0.19] | 0.02 [-0.01, 0.04] | 0.245 |
| 80+ | 0.18 [0.16, 0.20] | 0.16 [0.13, 0.19] | -0.02 [-0.06, 0.01] | 0.248 |
| Depressive symptoms | | | | |
| 50–59 | 1.56 [1.48, 1.64] | 1.75 [1.59, 1.91] | 0.19 [0.02, 0.37] | 0.002 |
| 60–69 | 1.44 [1.37, 1.51] | 1.71 [1.60, 1.83] | 0.27 [0.14, 0.41] | <0.001 |
| 70–79 | 1.55 [1.46, 1.64] | 1.61 [1.51, 1.71] | 0.06 [-0.07, 0.19] | 0.375 |
| 80+ | 1.87 [1.74, 2.01] | 1.69 [1.56, 1.82] | -0.18 [-0.37, 0.00] | 0.054 |
| CASP-19 | | | | |
| 50–59 | 42.15 [41.78, 42.53] | 39.52 [38.63, 40.41] | -2.63 [-3.61, -1.66] | <0.001 |
| 60–69 | 42.19 [41.82, 42.57] | 39.11 [38.49, 39.74] | -3.08 [-3.81, -2.35] | <0.001 |
| 70–79 | 40.58 [40.09, 41.06] | 39.50 [38.96, 40.04] | -1.08 [-1.81, -0.35] | 0.004 |
| 80+ | 37.13 [36.29, 41.38] | 37.01 [36.29, 37.74] | -0.11 [-1.21, 0.98] | 0.840 |
| Memory | | | | |
| 50–59 | 11.36 [11.24, 11.48] | 11.34 [11.07, 11.61] | -0.02 [-0.32, 0.28] | 0.890 |
| 60–69 | 10.36 [10.24, 10.47] | 10.81 [10.62, 11.00] | 0.45 [0.23, 0.68] | <0.001 |
| 70–79 | 9.10 [8.95, 9.24] | 10.29 [10.13, 10.45] | 1.20 [0.98, 1.42] | <0.001 |
| 80+ | 7.05 [6.82, 7.27] | 8.17 [7.94, 8.41] | 1.13 [0.80, 1.45] | <0.001 |

Data source: English Longitudinal Study of Ageing (ELSA) waves 2 (2004–05), 6 (2012–13) and 11 (2023–24). Notes: Estimates are adjusted predicted probabilities from regression models including age group x time interaction and controlling for age groups, sex, education, and wealth quintiles. Absolute change is expressed in percentage points and calculated as the adjusted predicted probability in 2023–24 minus that in 2004–05. Confidence intervals for the change test whether the adjusted predicted probabilities differ between the two survey periods (with relevant p-value).

#### Table S8. Within-education adjusted predicted probabilities and absolute percentage-point changes in health outcomes, 2004/05 to 2023/24

| Health outcomes | Predicted Probability 2004–05 | Predicted Probability 2023–24 | Absolute change [95% CI] | p-values for trend |
|---|---|---|---|---|
| ***Panel A. Self-rated health, pain, and functional limitations*** | | | | |
| Poor/fair self-rated health | | | | |
| High edu | 0.19 [0.16, 0.22] | 0.22 [0.19, 0.24] | 0.03 [-0.01, 0.07] | 0.121 |
| Medium edu | 0.25 [0.24, 0.27] | 0.32 [0.30, 0.34] | 0.06 [0.04, 0.09] | 0.002 |
| Low edu | 0.34 [0.32, 0.35] | 0.44 [0.40, 0.48] | 0.10 [0.06, 0.14] | <0.001 |
| Limiting long-standing illness | | | | |
| High edu | 0.33 [0.29, 0.36] | 0.28 [0.25, 0.31] | -0.05 [-0.09, -0.01] | 0.027 |
| Medium edu | 0.35 [0.34, 0.37] | 0.38 [0.36, 0.40] | 0.03 [0.00, 0.05] | 0.050 |
| Low edu | 0.37 [0.36, 0.39] | 0.43 [0.39, 0.46] | 0.05 [0.01, 0.10] | 0.009 |
| Any pain | | | | |
| High edu | 0.29 [0.26, 0.32] | 0.38 [0.35, 0.41] | 0.09 [0.05, 0.14] | <0.001 |
| Medium edu | 0.38 [0.36, 0.39] | 0.46 [0.44, 0.48] | 0.08 [0.06, 0.11] | <0.001 |
| Low edu | 0.41 [0.39, 0.43] | 0.56 [0.52, 0.60] | 0.15 [0.11, 0.20] | <0.001 |
| Moderate/severe pain | | | | |
| High edu | 0.19 [0.16, 0.22] | 0.21 [0.18, 0.23] | 0.02 [-0.02, 0.05] | 0.345 |
| Medium edu | 0.27 [0.26, 0.28] | 0.31 [0.29, 0.33] | 0.04 [0.02, 0.07] | <0.001 |
| Low edu | 0.31 [0.29, 0.32] | 0.43 [0.39, 0.47] | 0.13 [0.09, 0.17] | <0.001 |
| Any ADL limitations | | | | |
| High edu | 0.15 [0.13, 0.18] | 0.13 [0.11, 0.15] | -0.03 [-0.06, 0.01] | 0.125 |
| Medium edu | 0.21 [0.19, 0.22] | 0.20 [0.18, 0.21] | -0.01 [-0.03, 0.01] | 0.464 |
| Low edu | 0.22 [0.21, 0.24] | 0.24 [0.21, 0.27] | 0.02 [-0.01, 0.05] | 0.250 |
| Any IADL limitations | | | | |
| High edu | 0.18 [0.15, 0.22] | 0.13 [0.11, 0.15] | -0.05 [-0.09, -0.02] | 0.003 |
| Medium edu | 0.23 [0.21, 0.24] | 0.21 [0.19, 0.22] | -0.02 [-0.04, 0.00] | 0.105 |
| Low edu | 0.24 [0.23, 0.25] | 0.28 [0.24, 0.31] | 0.04 [0.00, 0.07] | 0.039 |
| Any mobility impairments | | | | |
| High edu | 0.50 [0.47, 0.54] | 0.39 [0.36, 0.42] | -0.11 [-0.16, -0.07] | <0.001 |
| Medium edu | 0.60 [0.58, 0.62] | 0.52 [0.50, 0.55] | -0.07 [-0.10, -0.05] | <0.001 |
| Low edu | 0.61 [0.59, 0.62] | 0.62 [0.59, 0.66] | 0.01 [-0.02, 0.05] | 0.446 |
| ***Panel B. Chronic conditions, obesity, and inflammation*** | | | | |
| Self-reported hypertension | | | | |
| High edu | 0.32 [0.29, 0.35] | 0.33 [0.30, 0.36] | 0.01 [-0.03, 0.05] | 0.604 |
| Medium edu | 0.35 [0.34, 0.37] | 0.40 [0.37, 0.42] | 0.04 [0.02, 0.07] | 0.001 |
| Low edu | 0.36 [0.34, 0.37] | 0.46 [0.42, 0.50] | 0.10 [0.06, 0.14] | <0.001 |
| Hypertension biomarker | | | | |
| High edu | 0.35 [0.31, 0.39] | 0.26 [0.23, 0.30] | -0.09 [-0.14, -0.04] | <0.001 |
| Medium edu | 0.38 [0.36, 0.40] | 0.30 [0.28, 0.33] | -0.08 [-0.10, -0.05] | <0.001 |
| Low edu | 0.40 [0.38, 0.42] | 0.31 [0.26, 0.36] | -0.09 [-0.14, -0.04] | <0.001 |
| Self-reported diabetes | | | | |
| High edu | 0.09 [0.07, 0.11] | 0.09 [0.07, 0.11] | 0.00 [-0.03, 0.03] | 0.921 |
| Medium edu | 0.08 [0.07, 0.09] | 0.13 [0.12, 0.15] | 0.05 [0.03, 0.07] | <0.001 |
| Low edu | 0.09 [0.08, 0.10] | 0.15 [0.12, 0.18] | 0.07 [0.04, 0.10] | <0.001 |

| | | | | |
|---|---|---|---|---|
| Diabetes biomarker | | | | |
| High edu | 0.07 [0.04, 0.09] | 0.07 [0.05, 0.09] | 0.01 [-0.02, 0.04] | 0.724 |
| Medium edu | 0.07 [0.06, 0.08] | 0.11 [0.09, 0.12] | 0.04 [0.02, 0.06] | <0.001 |
| Low edu | 0.07 [0.06, 0.08] | 0.13 [0.09, 0.18] | 0.06 [0.02, 0.11] | 0.009 |
| Self-reported high cholesterol | | | | |
| High edu | 0.18 [0.15, 0.21] | 0.34 [0.31, 0.36] | 0.16 [0.12, 0.20] | <0.001 |
| Medium edu | 0.19 [0.18, 0.21] | 0.38 [0.36, 0.40] | 0.19 [0.17, 0.21] | <0.001 |
| Low edu | 0.19 [0.18, 0.21] | 0.44 [0.39, 0.48] | 0.25 [0.21, 0.29] | <0.001 |
| High cholesterol biomarker | | | | |
| High edu | 0.79 [0.76, 0.82] | 0.63 [0.59, 0.66] | -0.16 [-0.21, -0.12] | <0.001 |
| Medium edu | 0.76 [0.75, 0.78] | 0.53 [0.50, 0.56] | -0.23 [-0.27, -0.20] | <0.001 |
| Low edu | 0.77 [0.75, 0.80] | 0.45 [0.38, 0.52] | -0.32 [-0.40, -0.25] | <0.001 |
| Self-reported lung disease | | | | |
| High edu | 0.03 [0.02, 0.04] | 0.03 [0.02, 0.04] | 0.01 [-0.01, 0.02] | 0.403 |
| Medium edu | 0.05 [0.04, 0.06] | 0.06 [0.05, 0.07] | 0.01 [-0.01, 0.02] | 0.248 |
| Low edu | 0.06 [0.05, 0.07] | 0.08 [0.06, 0.10] | 0.02 [0.00, 0.05] | 0.048 |
| Self-reported stroke | | | | |
| High edu | 0.01 [0.00, 0.02] | 0.01 [0.00, 0.02] | -0.00 [-0.01, 0.01] | 0.611 |
| Medium edu | 0.02 [0.01, 0.02] | 0.02 [0.01, 0.02] | -0.00 [-0.01, 0.01] | 0.870 |
| Low edu | 0.02 [0.01, 0.02] | 0.01 [0.01, 0.02] | -0.00 [-0.01, 0.00] | 0.362 |
| Self-reported heart diseases | | | | |
| High edu | 0.15 [0.12, 0.18] | 0.16 [0.14, 0.18] | 0.01 [-0.03, 0.04] | 0.648 |
| Medium edu | 0.16 [0.14, 0.17] | 0.18 [0.16, 0.20] | 0.02 [0.00, 0.04] | 0.022 |
| Low edu | 0.14 [0.13, 0.15] | 0.18 [0.15, 0.21] | 0.04 [0.01, 0.07] | 0.020 |
| Obesity biomarker | | | | |
| High edu | 0.22 [0.19, 0.25] | 0.27 [0.24, 0.30] | 0.05 [0.01, 0.10] | 0.016 |
| Medium edu | 0.27 [0.26, 0.29] | 0.33 [0.31, 0.36] | 0.06 [0.04, 0.09] | <0.001 |
| Low edu | 0.33 [0.31, 0.35] | 0.39 [0.33, 0.45] | 0.06 [0.00, 0.12] | 0.049 |
| Elevated C-reactive protein | | | | |
| High edu | 0.27 [0.24, 0.31] | 0.20 [0.16, 0.23] | -0.08 [-0.13, -0.03] | 0.002 |
| Medium edu | 0.38 [0.35, 0.39] | 0.24 [0.21, 0.26] | -0.13 [-0.16, -0.10] | <0.001 |
| Low edu | 0.41 [0.39, 0.43] | 0.28 [0.22, 0.34] | -0.13 [-0.19, -0.07] | <0.001 |
| Self-reported arthritis | | | | |
| High edu | 0.29 [0.26, 0.32] | 0.31 [0.29, 0.34] | 0.02 [-0.02, 0.06] | 0.264 |
| Medium edu | 0.36 [0.34, 0.38] | 0.37 [0.35, 0.39] | 0.01 [-0.01, 0.04] | 0.279 |
| Low edu | 0.36 [0.35, 0.38] | 0.40 [0.36, 0.44] | 0.04 [-0.00, 0.08] | 0.076 |
| ***Panel C. Mental health and cognition*** | | | | |
| Self-reported psychiatric problems | | | | |
| High edu | 0.08 [0.06, 0.10] | 0.10 [0.08, 0.11] | 0.02 [-0.01, 0.04] | 0.177 |
| Medium edu | 0.07 [0.06, 0.08] | 0.11 [0.09, 0.12] | 0.04 [0.02, 0.05] | <0.001 |
| Low edu | 0.06 [0.05, 0.07] | 0.11 [0.08, 0.14] | 0.05 [0.02, 0.08] | 0.001 |
| Depression | | | | |
| High edu | 0.12 [0.09, 0.14] | 0.12 [0.10, 0.14] | 0.01 [-0.02, 0.04] | 0.641 |
| Medium edu | 0.14 [0.13, 0.15] | 0.17 [0.15, 0.18] | 0.02 [0.00, 0.04] | 0.032 |
| Low edu | 0.19 [0.17, 0.20] | 0.24 [0.20, 0.27] | 0.05 [0.01, 0.09] | 0.010 |
| Depressive symptoms | | | | |
| High edu | 1.35 [1.25, 1.46] | 1.35 [1.25, 1.44] | -0.01 [-0.14, 0.13] | 0.927 |
| Medium edu | 1.51 [1.45, 1.57] | 1.61 [1.52, 1.70] | 0.10 [-0.01, 0.20] | 0.073 |
| Low edu | 1.80 [1.73, 1.88] | 2.15 [1.92, 2.39] | 0.35 [0.11, 0.59] | 0.005 |

| | | | | |
|---|---|---|---|---|
| CASP-19 | | | | |
| High edu | 42.58 [42.05, 43.10] | 41.20 [40.61, 41.79] | -1.38 [-2.12, -0.63] | <0.001 |
| Medium edu | 41.68 [41.37, 41.98] | 39.29 [38.79, 39.79] | -2.38 [-2.96, -1.81] | <0.001 |
| Low edu | 39.30 [38.88, 39.71] | 37.24 [36.03, 38.45] | -2.06 [-3.32, -0.80] | 0.001 |
| Memory | | | | |
| High edu | 10.97 [10.78, 11.17] | 11.71 [11.48, 11.95] | 0.74 [0.45, 1.03] | <0.001 |
| Medium edu | 10.20 [10.10, 10.30] | 10.75 [10.60, 10.90] | 0.56 [0.38, 0.73] | <0.001 |
| Low edu | 8.83 [8.71, 8.94] | 9.40 [9.08, 9.72] | 0.57 [0.24, 0.90] | <0.001 |

Data source: English Longitudinal Study of Ageing (ELSA) waves 2 (2004–05), 6 (2012–13) and 11 (2023–24). Notes: Estimates are adjusted predicted probabilities from regression models including education x time interaction and controlling for age groups, sex, education, and wealth quintiles. Absolute change is expressed in percentage points and calculated as the adjusted predicted probability in 2023–24 minus that in 2004–05. Confidence intervals for the change test whether the adjusted predicted probabilities differ between the two survey periods (with relevant p-value).
