## Supplementary File 2 for "Diverging trends in health at older ages in England, 2004–2024: evidence from the English Longitudinal Study of Ageing"

```
* Health trends, ELSA
* code version: 20260614
* waves 2 (2004/05), 6 (2012/13), 11 (2023/24)

cd "C:\Users\Data\ELSA Health trends" // for replication, replace this with the location of the folder with all ELSA datasets
clear all
cls

** To ensure loops work correctly, rename files before running this .do file
** if re-run code, do not start from here

    foreach j in 2 6 11 {
    use "wave_`j'_elsa_data.dta", clear

    order _all, alphabetic
            rename *, lower
            foreach var of varlist _all {
                    rename `var' `var'_w`j'
            }
    generate inWave`j'=1
    rename idauniq_w`j' idauniq
    save, replace
    }

    foreach j in 2 6 11 {
    use "wave_`j'_financial_derived_variables.dta", clear

    order _all, alphabetic
            rename *, lower
            foreach var of varlist _all {
                    rename `var' `var'_w`j'
            }
    generate inWave`j'=1
    rename idauniq_w`j' idauniq
    save, replace
    }


    foreach j in 2 6 11 {
    use "wave_`j'_ifs_derived_variables.dta", clear

    order _all, alphabetic
            rename *, lower
            foreach var of varlist _all {
                    rename `var' `var'_w`j'
            }
    generate inWave`j'=1
    rename idauniq_w`j' idauniq
    save, replace
    }

    foreach j in 2 6 11 {
    use "wave_`j'_nurse_data.dta", clear

    order _all, alphabetic
            rename *, lower
            foreach var of varlist _all {
                    rename `var' `var'_w`j'
            }
    generate inWave`j'=1
    rename idauniq_w`j' idauniq
    save, replace
```

```
    }


// for each wave, create a master dataset
    foreach j in 2 6 11 {
    use "wave_`j'_elsa_data.dta", clear
    sort idauniq
    merge 1:1 idauniq using "wave_`j'_financial_derived_variables"
    drop if _merge == 2
    drop _merge

    sort idauniq
    merge 1:1 idauniq using "wave_`j'_ifs_derived_variables"
    drop if _merge == 2
    drop _merge

    sort idauniq
    merge 1:1 idauniq using "wave_`j'_nurse_data"
    drop if _merge == 2
    drop _merge

    save "temp`j'", replace
}

**********************
**********************

* We now combine these three datasets to create 1 grand dataset for Waves 2 6 11
    clear all
    cls
    set maxvar 120000, perma
    use temp2.dta, clear
    foreach i in 6 11 {
            merge 1:1 idauniq using "temp`i'"
            drop _merge
            }

    count  // 17938 respondents - Note that some are in multiple waves

    egen ELSA_inwave2_6_11 = concat(inWave2 inWave6 inWave11)
    fre ELSA_inwave2_6_11 // 2079 R (11.6%) interviewed in w2, w6, and w11

* ###############################################################################
* AAA. SAMPLE SELECTION (ELSA core members, aged 50+, no proxy, valid cross-sectional weight (i.e. >0))

* Age --> indager
        foreach j in 2 6 11 {
            recode indager_w`j' 50/59=1 60/69=2 70/79=3 80/max=4 -7=4 -8=. min/49=. , into(age_group_4cat_w`j') // -7 means aged 90+
        }
        label define age_group_4cat_w 1"50/59" 2"60/69" 3"70/79" 4"80+", replace
        label values age_group_4cat_w* age_group_4cat_w

* core members
    drop if inWave2 == 1 & sampsta_w2 !=1
    drop if inWave6 == 1 & (finstatw6_w6 != 1 & finstatw6_w6 != 7 & finstatw6_w6 != 14 & finstatw6_w6 != 25)
    drop if inWave11 == 1 & corepartner_w11 == 0
* age 50+
    drop if (age_group_4cat_w2==. & inWave2==1) | (age_group_4cat_w6==. & inWave6==1) | (age_group_4cat_w11==. & inWave11==1)

```

```
* We get rid of PROXY respondents
    drop if askpx1_w2==1 | askpx_w6==1 | askpx_w11==1

* Rename weights (main interview, self completion, nurse visit, blood) and only retain those with valid cross-sectional weight (i.e. >0)
    rename (w2wgt_w2 w6xwgt_w6 w11xwgt_w11) (x_weight_w2 x_weight_w6 x_weight_w11)
    rename (w6scwt_w6 w11scwt_w11) (sc_weight_w6 sc_weight_w11)
    rename scw2wgt_w2 sc_weight_w2
    rename (w2wtnur_w2 w6nurwt_w6 w11nurwt_w11) (nurse_weight_w2 nurse_weight_w6 nurse_weight_w11)
    rename w2wtbld_w2 blood_weight_w2
    rename (w6bldwt_w6 w11bldwt_w11) (blood_weight_w6 blood_weight_w11)

    count // 14,664 respondents

    drop if (x_weight_w2 <=0 & inWave2==1) | (x_weight_w6 <=0 & inWave6==1) | (x_weight_w11 <=0 & inWave11==1)
    // 62 R are dropped
    count // 14,602 R

save "C:\Users\Data\ELSA Health trends\ELSA_temp2611.dta", replace
***
use "C:\Users\Data\ELSA Health trends\ELSA_temp2611.dta", clear

********* BBB Variables
* Sex --> indsex
        foreach k in 2 6 11 {
            generate female_w`k'=(indsex_w`k'==2) if indsex_w`k'!=.
        }

* Education
    foreach k in 2 6 11 {
        recode edqual_w`k' (1=1)    /// Degree, teaching qual., other higher qual., nursing qual.
        (3/5=2)         /// A-Level, City & Guilds Ordinary National Certificate, O-Level
        (6=2)           /// Commercial qual., apprenticeship, other qual.
        (7=3)           /// No quals, never went to school.
        (-9/-1=.)       /// Incomplete, DK
        , gen(education_w`k')
}
    label define education 1"1 Post-secondary" 2"2 A-Level, O-Level, CSE, technical equiv" 3"3 No academic/technical qualifications", replace
    label values education* education

* Wealth quintiles
    foreach k in 2 6 11 {
    rename tnhwq5_bu_s_w`k' wealth_q_w`k'
}
    label define  wealth_q 1 "1st quintile lowest" 2"2nd" 3"3rd" 4"4th" 5 "5th quintile highest", replace
    label values wealth_q* wealth_q

********* HEALTH OUTCOMES
*** Panel A. Self-rated health, pain, and functional limitations
*** Self-rated health, Limiting long-standing illness,
*** Any pain, Severe pain, ADL, IADL, Mobility
**# Bookmark #1

* Self-rated health --> hehelf
    foreach j in 2 6 11 {
            recode hehelf_w`j' 1/3=0 4/5=1 -9/-1=., into(srh_w`j')
        }
        label define srh_w 0">=Good" 1"Fair or Poor", replace
        label values srh_w* srh_w
        fre srh_w*
```

```
* Limiting long-standing illness
        recode heill_w* helim* (-9/-1=.)
        foreach j in 2 6 11 {
            gen gali_w`j'=.
            replace gali_w`j'=0 if heill_w`j'==2
            replace gali_w`j'=1 if heill_w`j'==1 & helim_w`j'==2
            replace gali_w`j'=2 if heill_w`j'==1 & helim_w`j'==1
            }
        label define gali_w 0"no illness" 1"illness not limiting" 2"limiting illness", replace
        label values gali_w* gali_w
        fre gali_w*

        foreach j in 2 6 11 {
            recode gali_w`j' 0/1=0 2=1, into(longill_w`j')
            }
        label define longill_w 0"no" 1"limiting longstanding illness", replace
        label values longill_w* longill_w
        fre longill_w*

* Any pain
        recode hepain* (-9/-1=.)
        foreach j in 2 6 11 {
            recode hepain_w`j' (1=1 "Yes") (2=0 "No"), gen(pain_w`j')
        }
        fre pain_w*

* Severe pain
        foreach j in 2 6 11 {
            gen severe_pain_w`j' = 0
            replace severe_pain_w`j' = 1 if inrange(hepaa_w`j',2,3)
            replace severe_pain_w`j' = . if hepain_w`j' < 0
            replace severe_pain_w`j' = . if hepaa_w`j' == -8
            label define severe_pain_w`j' 1 "severe/moderate pain" 0 "mild pain or no pain", replace
            label values severe_pain_w`j' severe_pain_w`j'
            replace severe_pain_w`j' = . if inWave`j' != 1
        }
        fre severe_pain_w*

* ADL (for consistency, we use SIX items: dressing, walking, bathing, eating, getting out of bed,
toilet)
        egen adl_w2=anycount(headb*_w2), v(1 2 3 4 5 6)
        replace adl_w2=. if headb01_w2<=-1
        replace adl_w2=. if inWave2!=1

        foreach j in 6 11 {
            egen adl_w`j'=anycount(headldr_w`j' headlwa_w`j' headlba_w`j' headlea_w`j' headlbe_w`j'
headlwc_w`j'), v(1)
            replace adl_w`j'=. if headl96_w`j'<-1
            replace adl_w`j'=. if inWave`j'!=1
        }
        foreach j in 2 6 11 {
            recode adl_w`j' (0=0 "No ADL") (1/6=1 "1+ ADL"), gen(adl_bin_w`j')
        }
        fre adl_w*
        fre adl_bin_w*

* IADL (for consistency, we use SEVEN items: map, meal, shop, phone, medicine, house, money)
        egen iadl_w2=anycount(headb*_w2), v(7 8 9 10 11 12 13)
        replace iadl_w2=. if headb01_w2<=-1
        replace iadl_w2=. if inWave2!=1
        foreach j in 6 11 {
            egen iadl_w`j'=anycount(headlma_w`j' headlpr_w`j' headlsh_w`j' ///
```

```
            headlph_w`j' headlme_w`j' headlho_w`j' headlmo_w`j'), v(1)
            replace iadl_w`j'=. if headl96_w`j'<-1
            replace iadl_w`j'=. if inWave`j'!=1
        }
        foreach j in 2 6 11 {
            recode iadl_w`j' (0=0 "No IADL") (1/7=1 "1+ IADL"), gen(iadl_bin_w`j')
        }
        fre iadl_w*
        fre iadl_bin_w*

* mobility impairments
    egen mobiwalk_w2 = anycount(heada01 heada02 heada03 heada04 heada05 heada06 heada07 heada08 heada09 heada10), v(1)
    egen mobisit_w2 = anycount(heada01 heada02 heada03 heada04 heada05 heada06 heada07 heada08 heada09 heada10), v(2)
    egen mobigetup_w2 = anycount(heada01 heada02 heada03 heada04 heada05 heada06 heada07 heada08 heada09 heada10), v(3)
    egen mobimultiflight_w2 = anycount(heada01 heada02 heada03 heada04 heada05 heada06 heada07 heada08 heada09 heada10), v(4)
    egen mobioneflight_w2 = anycount(heada01 heada02 heada03 heada04 heada05 heada06 heada07 heada08 heada09 heada10), v(5)
    egen mobistoop_w2 = anycount(heada01 heada02 heada03 heada04 heada05 heada06 heada07 heada08 heada09 heada10), v(6)
    egen mobiarm_w2 = anycount(heada01 heada02 heada03 heada04 heada05 heada06 heada07 heada08 heada09 heada10), v(7)
    egen mobipull_w2 = anycount(heada01 heada02 heada03 heada04 heada05 heada06 heada07 heada08 heada09 heada10), v(8)
    egen mobi10pound_w2 = anycount(heada01 heada02 heada03 heada04 heada05 heada06 heada07 heada08 heada09 heada10), v(9)
    egen mobicoin_w2 = anycount(heada01 heada02 heada03 heada04 heada05 heada06 heada07 heada08 heada09 heada10), v(10)

    egen mobi_number_w2 = rowtotal(mobiwalk_w2 mobisit_w2 mobigetup_w2 mobimultiflight_w2 mobioneflight_w2 ///
    mobistoop_w2 mobiarm_w2 mobipull_w2 mobi10pound_w2 mobicoin_w2)
    replace mobi_number_w2 = . if heada01 == -8 & heada02 == -1 & heada03 == -1 ///
    & heada04 == -1 & heada05 == -1 & heada06 == -1 & ///
    heada07 == -1 & heada08 == -1 & heada09 == -1 & heada10 == -1
    recode mobi_number_w2 (1/10 = 1 "yes, >=1 mobility impairments") (0 = 0 "no mobility impairment"), gen(mobi_if_w2)
    tab mobi_number_w2 mobi_if_w2
    fre mobi_if_w2 if inWave2==1
    sum mobi_number_w2
    replace mobi_number_w2 = . if inWave2 !=1
    replace mobi_if_w2 = . if inWave2 !=1
    sum mobi_number_w2

    egen mobi_number_w6 = anycount(hemobwa_w6 hemobsi_w6 hemobch_w6 hemobcs_w6 hemobcl_w6 hemobst_w6 ///
                                   hemobre_w6 hemobpu_w6 hemobli_w6 hemobpi_w6), v(1)
    replace mobi_number_w6 = . if hemobwa_w6 <0 & hemobsi_w6 <0 & hemobch_w6 <0 & hemobcs_w6 <0 & hemobcl_w6 < 0 & ///
                                  hemobst_w6 <0 & hemobre_w6 <0 & hemobpu_w6 <0 & hemobli_w6 <0 & hemobpi_w6 < 0
    recode mobi_number_w6 (1/10 = 1 "yes, >=1 mobility impairments") (0 = 0 "no mobility impairments"), gen(mobi_if_w6)
    tab mobi_number_w6 mobi_if_w6
    fre mobi_if_w6
    sum mobi_number_w6
    replace mobi_number_w6 = . if inWave6 !=1
    replace mobi_if_w6 = . if inWave6 !=1
```

```
    egen mobi_number_w11 = anycount(hemobwa_w11 hemobsi_w11 hemobch_w11 hemobcs_w11 hemobcl_w11 hemobst_w11 ///
                                   hemobre_w11 hemobpu_w11 hemobli_w11 hemobpi_w11), v(1)
    replace mobi_number_w11 = . if hemobwa_w11 <0 & hemobsi_w11 <0 & hemobch_w11 <0 & hemobcs_w11 <0 & hemobcl_w11 < 0 & ///
                                  hemobst_w11 <0 & hemobre_w11 <0 & hemobpu_w11 <0 & hemobli_w11 <0 & hemobpi_w11 < 0
    recode mobi_number_w11 (1/11 = 1 "yes, >=1 mobility impairments") (0 = 0 "no mobility impairments"), gen(mobi_if_w11)
    tab mobi_number_w11 mobi_if_w11
    fre mobi_if_w11
    sum mobi_number_w11
    replace mobi_number_w11 = . if inWave11 !=1
    replace mobi_if_w11 = . if inWave11 !=1

    fre mobi_if_w*

**# Bookmark #2
* Panel B. Chronic conditions (SELF-REPORTED)
* CHRONIC CONDITIONS
    * Hypertension, diabetes, high cholesterol
    * Lung disease
    * Stroke
    * Other heart disease [angina, heart attack, heart failure, heart murmur, abnormal heart rhythm]

* HYPERTENSION

    * In Wave 2  // 1786 people with feedforward issue - we deal with this issue by assigning randomly values based on observed prevalence.
        egen hedim_hypertension_w2 = anycount(hedim*_w2), v(1)    // doctor ever told you ...
        // use hedim*_w2 rather than hedia*_w2 to account for R's other answer that is codeable to hypertension (1 more obs in this case)
        tab hediac1_w2 hedim_hypertension_w2

        count if hediac1_w2 == 1 // 1330 R confirmed W1 diagnosis of hypertension
        count if hediac1_w2 == 1 & hedias1_w2 == 1 // Among these 1330 R, 1020 R still have it in W2
        di 1020/1330 // About 77%
            * Based on real data w1-w2, we know that 77% R who confirmed W1 diagnosis of hypertension still have it in W2
            * We therfore randomly select 77% among those who reported having it in W1 but have feedforward issue in W2
        set seed 24284
        generate random_hyper = runiform() if hediac1_w2==-3

        gen hypertension_w2=0 if inWave2==1
        replace hypertension_w2=1 if hediac1_w2==1 & hedias1_w2==1 // people who had it at w1, did not dispute it, and still have condition
        replace hypertension_w2=1 if hediac1_w2!=1 & hedian1_w2==3 // not had it at w1 but have it now
        replace hypertension_w2=1 if hediac1_w2==-1 & hedim_hypertension_w2==1 // We assume those HyperT is current if condition not reported at w1 but YES at w2
        replace hypertension_w2=1 if hediac1_w2==-3 & random_hyper<0.77 // About 1367 added as having hypertension (77% of those who had it in wave 1 but were not asked the follow-up question for a feed forward problem)
        replace hypertension_w2 = . if hediac1_w2 == -8 // Confirm W1 diagnosis? Don't know
        replace hypertension_w2 = . if hedias1_w2 == -8 // Still have it in W2? Don't know
        replace hypertension_w2 = . if hedim01_w2 == -1 // -1 means not applicable which means "refusal" or "DK" on hedia01_w2 (newly report)
        ci mean hypertension_w2

    * In Wave 6 - We do not know if refreshment sample still have this chronic condition because they are not asked to answer this Q
        count if finstatw6_w6 == 25 // 796 Cohort-6 core members, i.e., refreshment samples added in W6
```

```
        generate hypertension_w6=0 if inWave6==1 & finstatw6_w6 != 25
        replace hypertension_w6=1 if hedacbp_w6==1 & hedasbp_w6==1 // people who had it at previous waves, did not dispute it, and still have condition
        replace hypertension_w6=1 if hedanbp_w6==3                 // not had before but R has it now
        replace hypertension_w6=1 if hedimbp_w6==1 & finstatw6_w6 != 25    // for non-refreshment samples, Q is about "since we last interviewed you"
        replace hypertension_w6=. if hedacbp_w6 == -8 // Confirm previous diagnosis? Don't know
        replace hypertension_w6=. if hedasbp_w6 == -8 // Still have it in W6? Don't know
        replace hypertension_w6=. if hedimbp_w6 <=-8 // newly reported in W6? Refusal or don't know

    * In Wave 11 ** Simple routing in this wave - Do you still have it if you reported it before, if new report, and new among refreshment!
        gen hypertension_w11=0 if inWave11==1
        replace hypertension_w11=1 if (hehavebp_w11==1 | hehavebp_w11==2)   // 1 = have hypertension but does not take medication
                                                                            // 2 = have hypertension and take medication
        replace hypertension_w11=. if hehavebp_w11<-1 // Don't know
    ci mean hypertension_w*

// DIABETES
    * * In Wave 2  // 355 people with feedforward issue
    egen hedim_diabetes_w2 = anycount(hedim*_w2), v(7)    // doctor ever told you ...
        // use hedim*_w2 rather than hedia*_w2 to account for R's other answer that is codeable to diabetes (1 more obs in this case)
        tab hediac7_w2 hedim_diabetes_w2

        count if hediac7_w2 == 1 // 235 R confirmed W1 diagnosis of diabetes
        count if hediac7_w2 == 1 & hedias7_w2 == 1 // Among these 235 R, 230 R still have it in W2
        di 230/235 // 98%
        * so based on real data w1-w2, we know that 98% R who confirmed W1 diagnosis of diabetes still have it in W2
        * We could randomly select 98% among those who reported having it in W1 but have feedforward issue in W2
        set seed 24284
        generate random_diabetes_w2 = runiform() if hediac7_w2==-3
        gen diabetes_w2=0 if inWave2==1
        replace diabetes_w2=1 if hediac7_w2==1 & hedias7_w2==1 // people who had it at w1, did not dispute it, and still have condition
        replace diabetes_w2=1 if hediac7_w2!=1 & hedian7_w2==3 // not had it at w1 but have it now
        replace diabetes_w2=1 if hediac7_w2==-1 & hedim_diabetes_w2==1 // We assume their diabetes is current if condition not reported at w1 but YES at w2
        replace diabetes_w2=1 if hediac7_w2==-3 & random_diabetes_w2 <0.98 // this code did not necessarily turn 98%x235 R with value -3 on hediac7_w2 to "having diabetes" in W2 because some R may have already got value 1 on diabetes_w2

        replace diabetes_w2 = . if hediac7_w2 == -8 // Confirm W1 diagnosis? Don't know
        replace diabetes_w2 = . if hedias7_w2 == -8 // Still have it in W2? Don't know
        replace diabetes_w2 = . if hedim01_w2 == -1 // -1 means not applicable which means "refusal" or "DK" on hedia01_w2 (newly report)

    * In Wave 6
        count if finstatw6_w6 == 25 // 796 Cohort-6 core members, i.e., refreshment samples added in W6
        generate diabetes_w6=0 if inWave6==1 & finstatw6_w6 != 25
    // no question "do you still have diabetes" in W6, but we can use random numbers for those who confirmed previous diagnosis of diabetes. We take the 98% from wave 2 (those who had diabetes in previous wave and still have it now).
        set seed 24284
        generate random_diabetes_w6 = runiform() if hedacdi_w6 == 1
        replace diabetes_w6=1 if hedandi_w6==3                 // not had before but R has diabetes now
        replace diabetes_w6=1 if hedimdi_w6==1 & finstatw6_w6 != 25    // for non-refreshment
```

```
samples, Q is about "since we last interviewed you"
        replace diabetes_w6=1 if hedacdi_w6 == 1 & random_diabetes_w6 <0.98 // we assume 98% of R who confirmed previous diagnosis still have it
        replace diabetes_w6=. if hedacdi_w6 == -8 // Confirm previous diagnosis? Don't know
        replace diabetes_w6=. if hedimdi_w6 <=-8 // newly reported in W6? Refusal or don't know

    * In Wave 11 ** much more simplified - Do you still have it if you reported it before, if new report, and new among refreshment!!
        gen diabetes_w11=0 if inWave11==1
        replace diabetes_w11=1 if (hehavedi_w11==1 | hehavedi_w11==2)   // 1 = have diabetes but does not take medication
                                                                        // 2 = have diabetes and take medication
        replace diabetes_w11=. if hehavedi_w11<-1 // Don't know
    ci means diabetes_w*

// HIGH CHOLESTEROL
  * * In Wave 2. No question to confirm previous diagnosis so feedforward error is not applicable.
    egen hedim_cholesterol_w2 = anycount(hedim*_w2), v(9)    // doctor ever told you ...
        // use hedim*_w2 rather than hedia*_w2 to account for R's other answer that is codeable to heart rhythm (1 more obs in this case)
        * There is no HEDIAC9 to confirm W1 diagnosis of high cholesterol
        * There is no question "Do you still have high cholesterol?"
        gen cholesterol_w2=0 if inWave2==1
        replace cholesterol_w2=1 if hedim_cholesterol_w2==1 // We assume their high cholesterol is current otherwise the % is underestimated
        replace cholesterol_w2 = . if hedim01_w2 == -1 // -1 means not applicable which means "refusal" or "DK" on hedia01_w2 (newly report)

    * In Wave 6 - Again, we assign missing values to all refreshment samples
        count if finstatw6_w6 == 25 // 796 Cohort-6 core members, i.e., refreshment samples added in W6
        generate cholesterol_w6=0 if inWave6==1 & finstatw6_w6 != 25
        replace cholesterol_w6=1 if hedacch_w6==1 & hedasch_w6==1 // people who had it at previous waves, did not dispute it, and still have condition
        replace cholesterol_w6=1 if hedanch_w6==3                 // not had before but R has it now
        replace cholesterol_w6=1 if hedimch_w6==1 & finstatw6_w6 != 25    // for non-refreshment samples, Q is about "since we last interviewed you"
        replace cholesterol_w6 = 1 if hedimch_w6 == 1 & hechmd_w6 == 1 // newly reported high cholesterol and take medication.
        replace cholesterol_w6=. if hedacch_w6 <= -8 // Confirm previous diagnosis? Don't know
        replace cholesterol_w6=. if hedasch_w6 <= -8 // Still have it in W6? Don't know
        replace cholesterol_w6=. if hedimch_w6 <=-8 // newly reported in W6? Refusal or don't know

    * In Wave 11 ** much more simplified - Do you still have it if you reported it before, if new report, and new among refreshment!!
        gen cholesterol_w11=0 if inWave11==1
        replace cholesterol_w11=1 if (hehavehc_w11==1 | hehavehc_w11==2)    // 1 = have high cholesterol but does not take medication
                                                                            // 2 = have high cholesterol  and take medication
        replace cholesterol_w11=. if hehavehc_w11<-1 // Don't know
   ci mean cholesterol_w*

// LUNG DISEASE
  ** In Wave 2. 1 observation with feedforward problem so we can ignore it
    egen hedib_lung_w2 = anycount(hedib01_w2 hedib02_w2 hedib03_w2 hedib04_w2), v(1)
        gen lung_w2=0 if inWave2==1
        replace lung_w2=1 if hediad1_w2 ==1 & hedids1_w2==1 // people who had it at w1, did not dispute it, and still have condition
        replace lung_w2=1 if hediad1_w2!=1 & hediam1_w2==3 // not had it at w1 but have it now
        replace lung_w2=1 if hediad1_w2==-1 & hedib_lung_w2==1 // We assume their lung disease is current if condition not reported at w1 but YES at w2
```

```
        replace lung_w2 = . if hediad1_w2 == -8 // Confirm W1 diagnosis? Don't know
        replace lung_w2 = . if hedids1_w2 == -8 // Still have it in W2? Don't know
        replace lung_w2 = . if hedib01_w2 <= -8 //  "refusal" or "DK" on hedib01_w2 (newly report)

    * In Wave 6
        count if finstatw6_w6 == 25 // 796 Cohort-6 core members, i.e., refreshment samples added in W6
        generate lung_w6=0 if inWave6==1 & finstatw6_w6 != 25
        replace lung_w6=1 if hedbdlu_w6==1 & hedblu_w6==1 // people who had it at previous waves, did not dispute it, and still have condition
        replace lung_w6=1 if hedbmlu_w6==3                 // not had before but R has it now
        replace lung_w6=1 if hediblu_w6==1 & finstatw6_w6 != 25    // for non-refreshment samples, Q is about "since we last interviewed you"
        replace lung_w6=. if hedbdlu_w6 <= -8 // Confirm previous diagnosis? Don't know
        replace lung_w6=. if hedblu_w6 <= -8 // Still have it in W6? Don't know
        replace lung_w6=. if hediblu_w6 <=-8 // newly reported in W6? Refusal or don't know

    * In Wave 11 ** much more simplified - Do you still have it if you reported it before, if new report, and new among refreshment!!
        gen lung_w11=0 if inWave11==1
        replace lung_w11=1 if (hehavecl_w11==1 | hehavecl_w11==2)   // 1 = have lung disease but does not take medication
                                                                     // 2 = have lung disease and take medication
        replace lung_w11=. if hehavecl_w11<-1 // Don't know
  ci means lung_w*

// STROKE
  * Wave 2. 183 R with feedforward errors
      * no question "Do you still have stroke?"
      * no question "Do you take medication for stroke?"
      egen hedim_stroke_w2 = anycount(hedim*_w2), v(8)    // doctor ever told you that you have stroke
        // use hedim*_w2 rather than hedia*_w2 to account for R's other answer that is codeable to stroke (4 more obs in this case)
        tab hediac8_w2 hedim_stroke_w2
        count if hediac8_w2 == 1 // 130 R confirmed W1 diagnosis of stroke
        count if hediac8_w2 == 1 & inrange(henmst_w2,1,3) // 6 R. The routing for henmst (number of strokes since last visit) is confirming W1 stroke OR nelwy reporting stroke in W2
        // those who confirmed W1 diagnosis of stroke may not newly report hedia* in W2 so did not answer henmst
        * so it is not possible to tell the proportion of R who confirmed W1 diagnosis of stroke and had it in last 2 years prior to W2
        gen stroke_w2=0 if inWave2==1
        replace stroke_w2=1 if hediac8_w2!=1 & hedian8_w2==3 // not had it at w1 but have it now
        replace stroke_w2=1 if hediac8_w2==-1 & hedim_stroke_w2==1 // We assume their stroke is current if condition not reported at w1 but YES at w2
        replace stroke_w2=1 if inrange(henmst_w2,1,3) // if had >=1 stroke since last visit

        replace stroke_w2 = . if hediac8_w2 == -8 // Confirm W1 diagnosis? Don't know
        replace stroke_w2 = . if hedim01_w2 == -1 // -1 means not applicable which means "refusal" or "DK" on hedia01_w2 (newly report)
        replace stroke_w2 = . if henmst_w2 == -8 // number of heart attacks since last visit? DK

    * In Wave 6
        count if finstatw6_w6 == 25 // 796 Cohort-6 core members, i.e., refreshment samples added in W6
        generate stroke_w6=0 if inWave6==1 & finstatw6_w6 != 25
        * no question "do you still have stroke?"
        replace stroke_w6=1 if hedanst_w6==3      // not had before but R has it now
        replace stroke_w6=1 if hedimst_w6==1 & finstatw6_w6 != 25    // for non-refreshment samples, Q is about "since we last interviewed you"
        replace stroke_w6 = 1 if inrange(henmst_w6,1,3) // number of stroke in the past 2 years
        replace stroke_w6=. if hedacst_w6 == -8 // Confirm previous diagnosis? Don't know
```

```
        replace stroke_w6=. if hedimst_w6 <=-8 // newly reported in W6? Refusal or don't know
        replace stroke_w6 = . if henmst_w6 == -8 // number of stroke in last 2 years? DK
        replace stroke_w6 = . if hemda1_w6 <= -8 // taking medication? DK

    * In Wave 11 ** no simple variable "hehavest"
        gen stroke_w11=0 if inWave11==1
        replace stroke_w11 = 1 if hediacst_w11 == 1 & heothst_w11 == 1 // had it before, not dispute and had another stroke since last diagnosis
        // the routing for heothst is feedforward = yes and not dispute, so heothst is similar to question "still have it"
        replace stroke_w11 = 1 if hediacst_w11==-1 & hediia7_w11 ==1 // We assume their stroke is current if condition not reported previously but YES in W11
        replace stroke_w11 = 1 if hemda1_w11 == 1 // taking medication for stroke
        replace stroke_w11=. if hediacst_w11 < -1 // confirm previous diagnosis of heart attack? Don't know
        replace stroke_w11=. if hediia7_w11 < -1 // newly reported heart attack? DK, refusal
        replace stroke_w11=. if heothst_w11 < -1 // Had a heart attack since last diagnosis? Don't know
        replace stroke_w11=. if hemda1_w11 < -1 // taking medication? DK
    ci means stroke_w*

// OTHER HEART DISEASES
// IF R had one of the following: heart failure, heart murmur, heart attack, abnormal heart rhythm, or angina.

** HEART FAILURE
  * * In Wave 2  // 34 people with feedforward issue
    egen hedim_hfailure_w2 = anycount(hedim*_w2), v(4)    // doctor ever told you ...
        // use hedim*_w2 rather than hedia*_w2 to account for R's other answer that is codeable to heart failure (6 more obs in this case)
        tab hediac4_w2 hedim_hfailure_w2
        count if hediac4_w2 == 1 // 14 R confirmed W1 diagnosis of hfailure
        count if hediac4_w2 == 1 & hedias4_w2 == 1 // Among these 14 R, 11 R still have it in W2
        di 11/14 // 79%
        * so based on real data w1-w2, we know that 79% R who confirmed W1 diagnosis of heart failure still have it in W2
        * We could randomly select 79% among those who reported having it in W1 but have feedforward issue in W2
        set seed 24284
        generate random_hfailure_w2 = runiform() if hediac4_w2==-3
        gen hfailure_w2=0 if inWave2==1
        replace hfailure_w2=1 if hediac4_w2==1 & hedias4_w2==1 // people who had it at w1, did not dispute it, and still have condition
        replace hfailure_w2=1 if hediac4_w2!=1 & hedian4_w2==3 // not had it at w1 but have it now
        replace hfailure_w2=1 if hediac4_w2==-1 & hedim_hfailure_w2==1 // We assume their heart failure is current if condition not reported at w1 but YES at w2
        replace hfailure_w2=1 if hediac4_w2==-3 & random_hfailure_w2 <0.79 // this code did not necessarily turn 79%x34 R with value -3 on hediac4_w2 to "having hfailure" in W2 because some R may have already got value 1 on hfailure_w2
        replace hfailure_w2 = . if hediac4_w2 == -8 // Confirm W1 diagnosis? Don't know
        replace hfailure_w2 = . if hedias4_w2 == -8 // Still have it in W2? Don't know
        replace hfailure_w2 = . if hedim01_w2 == -1 // -1 means not applicable which means "refusal" or "DK" on hedia01_w2 (newly report)

    * In Wave 6
        count if finstatw6_w6 == 25 // 796 Cohort-6 core members, i.e., refreshment samples added in W6
        generate hfailure_w6=0 if inWave6==1 & finstatw6_w6 != 25
        replace hfailure_w6=1 if hedachf_w6==1 & hedashf_w6==1 // people who had it at previous waves, did not dispute it, and still have condition
        replace hfailure_w6=1 if hedanhf_w6==3                 // not had before but R has it now
        replace hfailure_w6=1 if hedimhf_w6==1 & finstatw6_w6 != 25    // for non-refreshment samples, Q is about "since we last interviewed you"
```

```
        replace hfailure_w6=. if hedachf_w6 == -8 // Confirm previous diagnosis? Don't know
        replace hfailure_w6=. if hedashf_w6 == -8 // Still have it in W6? Don't know
        replace hfailure_w6=. if hedimhf_w6 <=-8 // newly reported in W6? Refusal or don't know

    * In Wave 11 ** much more simplified - Do you still have it if you reported it before, if new report, and new among refreshment!!
        gen hfailure_w11=0 if inWave11==1
        replace hfailure_w11=1 if (hehavehf_w11==1 | hehavehf_w11==2)   // 1 = have heart failure but does not take medication
                                                                                   // 2 = have heart failure and take medication
        replace hfailure_w11=. if hehavehf_w11<-1 // Don't know
    ci mean hfailure_w*


** HEART MURMUR
  * * In Wave 2  // 191 people with feedforward issue
    egen hedim_hmurmur_w2 = anycount(hedim*_w2), v(5)    // doctor ever told you ...
        // use hedim*_w2 rather than hedia*_w2 to account for R's other answer that is codeable to heart murmur (6 more obs in this case)
        tab hediac5_w2 hedim_hmurmur_w2
        count if hediac5_w2 == 1 // 152 R confirmed W1 diagnosis of hmurmur
        count if hediac5_w2 == 1 & hedias5_w2 == 1 // Among these 152 R, 100 R still have it in W2
        di 100/152 // 66%
        * so based on real data w1-w2, we know that 66% R who confirmed W1 diagnosis of heart murmur still have it in W2
        * We could randomly select 66% among those who reported having it in W1 but have feedforward issue in W2
        set seed 24284
        generate random_hmurmur_w2 = runiform() if hediac5_w2==-3
        gen hmurmur_w2=0 if inWave2==1
        replace hmurmur_w2=1 if hediac5_w2==1 & hedias5_w2==1 // people who had it at w1, did not dispute it, and still have condition
        replace hmurmur_w2=1 if hediac5_w2!=1 & hedian5_w2==3 // not had it at w1 but have it now
        replace hmurmur_w2=1 if hediac5_w2==-1 & hedim_hmurmur_w2==1 // We assume their heart murmur is current if condition not reported at w1 but YES at w2
        replace hmurmur_w2=1 if hediac5_w2==-3 & random_hmurmur_w2 <0.66 // this code did not necessarily turn 66%x191 R with value -3 on hediac5_w2 to "having hmurmur" in W2 because some R may have already got value 1 on hmurmur_w2
        replace hmurmur_w2 = . if hediac5_w2 == -8 // Confirm W1 diagnosis? Don't know
        replace hmurmur_w2 = . if hedias5_w2 == -8 // Still have it in W2? Don't know
        replace hmurmur_w2 = . if hedim01_w2 == -1 // -1 means not applicable which means "refusal" or "DK" on hedia01_w2 (newly report)

    * In Wave 6
        count if finstatw6_w6 == 25 // 796 Cohort-6 core members, i.e., refreshment samples added in W6
        generate hmurmur_w6=0 if inWave6==1 & finstatw6_w6 != 25
        replace hmurmur_w6=1 if hedachm_w6==1 & hedashm_w6==1 // people who had it at previous waves, did not dispute it, and still have condition
        replace hmurmur_w6=1 if hedanhm_w6==3                 // not had before but R has it now
        replace hmurmur_w6=1 if hedimhm_w6==1 & finstatw6_w6 != 25    // for non-refreshment samples, Q is about "since we last interviewed you"
        replace hmurmur_w6=. if hedachm_w6 == -8 // Confirm previous diagnosis? Don't know
        replace hmurmur_w6=. if hedashm_w6 == -8 // Still have it in W6? Don't know
        replace hmurmur_w6=. if hedimhm_w6 <=-8 // newly reported in W6? Refusal or don't know

    * In Wave 11 ** much more simplified - Do you still have it if you reported it before, if new report, and new among refreshment!!
        gen hmurmur_w11=0 if inWave11==1
        replace hmurmur_w11=1 if (hehavehm_w11==1 | hehavehm_w11==2)    // 1 = have heart murmur but does not take medication
                                                                                   // 2 = have heart murmur
```

```
and take medication
        replace hmurmur_w11=. if hehavehm_w11<-1 // Don't know
    ci mean hmurmur_w*

// HEART ATTACK
    * Wave 2. 260 R with feedforward errors
  egen hedim_hattack_w2 = anycount(hedim*_w2), v(3)    // doctor ever told you that you have heart attack
        // use hedim*_w2 rather than hedia*_w2 to account for R's other answer that is codeable to heart attack (8 more obs in this case)
        tab hediac3_w2 hedim_hattack_w2
        count if hediac3_w2 == 1 // 193 R confirmed W1 diagnosis of heart attack
        * There is NO question "do you still have a heart attack in W2".
        count if hediac3_w2 == 1 & inrange(henmmi_w2,1,3) // 5 R. The routing for henmmi (number of heart attacks since last visit) is having responded to W1 CVD questions AND reported heart attack in W2
        // those who confirmed W1 diagnosis of heart attack may not newly report hedia* in W2 so did not answer henmmi
        * It is not possible to determine the proportion of R who confirmed W1 diagnosis of heart attack and had it in last 2 years prior to W2

        gen hattack_w2=0 if inWave2==1
        replace hattack_w2=1 if hediac3_w2!=1 & hedian3_w2==3 // not had it at w1 but have it now
        replace hattack_w2=1 if hediac3_w2==-1 & hedim_hattack_w2==1 // We assume their heart attack is current if condition not reported at w1 but YES at w2
        replace hattack_w2=1 if hedim_hattack_w2 == 1 & hehrtb_w2 == 1 // newly reported heart attack and take medication. hehrtb is for either angina or heart attack so we also need to specify hedim_hattack_w2(newly reported)
        replace hattack_w2=1 if inrange(henmmi_w2,1,3) // if had >=1 heart attack since last visit
        replace hattack_w2 = . if hediac3_w2 == -8 // Confirm W1 diagnosis? Don't know
        replace hattack_w2 = . if hedim01_w2 == -1 // -1 means not applicable which means "refusal" or "DK" on hedia01_w2 (newly report)
        replace hattack_w2 = . if henmmi_w2 == -8 // number of heart attacks since last visit? DK
        replace hattack_w2 = . if hehrtb_w2 == -8 // taking medication? DK

    * In Wave 6
        count if finstatw6_w6 == 25 // 796 Cohort-6 core members, i.e., refreshment samples added in W6
        generate hattack_w6=0 if inWave6==1 & finstatw6_w6 != 25
        replace hattack_w6=1 if hedanmi_w6==3      // not had before but R has it now
        replace hattack_w6=1 if hedimmi_w6==1 & finstatw6_w6 != 25    // for non-refreshment samples, Q is about "since we last interviewed you"
        replace hattack_w6 = 1 if inrange(henmmi_w6,1,3) // number of heart attacks in the past 2 years
        replace hattack_w6 = 1 if hedimmi_w6 == 1 & hehrtb_w6 == 1 // newly reported heart attack and take medication. hehrtb is for either angina or heart attack so we also need to specify hedimmi_w2 (newly reported)

        replace hattack_w6=. if hedacmi_w6 == -8 // Confirm previous diagnosis? Don't know
        replace hattack_w6=. if hedimmi_w6 <=-8 // newly reported in W6? Refusal or don't know
        replace hattack_w6 = . if henmmi_w6 == -8 // number of heart attacks in last 2 years? DK
        replace hattack_w6 = . if hehrtb_w6 == -8 // taking medication? DK

    * In Wave 11 ** no simple variable "hehavemi"
        gen hattack_w11=0 if inWave11==1
        replace hattack_w11 = 1 if hediacmi_w11 == 1 & heothha_w11 == 1 // had it before, not dispute and had another heart attack since last diagnosis
        // the routing for heothha is feedforward = yes and not dispute, so heothha is similar to question "still have it"
        replace hattack_w11 = 1 if hediacmi_w11==-1 & hediia3_w11 ==1 // We assume their heart attack is current if condition not reported previously but YES in W11
        replace hattack_w11 = 1 if hemedha_w11 == 1 // taking medication for attack heart

```

```
        replace hattack_w11=. if hediacmi_w11 < -1 // confirm previous diagnosis of heart attack? Don't know
        replace hattack_w11=. if hediia3_w11 < -1 // newly reported heart attack? DK, refusal
        replace hattack_w11=. if heothha_w11 < -1 // Had a heart attack since last diagnosis? Don't know
        replace hattack_w11=. if hemedha_w11 < -1 // taking medication? DK
    ci mean hattack_w*

** ABNORMAL HEART RHYTHM
    * * In Wave 2
    egen hedim_hrhythm_w2 = anycount(hedim*_w2), v(6)    // doctor ever told you ...
        // use hedim*_w2 rather than hedia*_w2 to account for R's other answer that is codeable to heart rhythm (26 more obs in this case)
        tab hediac6_w2 hedim_hrhythm_w2
        gen hrhythm_w2=0 if inWave2==1
        replace hrhythm_w2=1 if hediac6_w2==1 & hedias6_w2==1 // people who had it at w1, did not dispute it, and still have condition
        replace hrhythm_w2=1 if hediac6_w2!=1 & hedian6_w2==3 // not had it at w1 but have it now
        replace hrhythm_w2=1 if hediac6_w2==-1 & hedim_hrhythm_w2==1 // We assume their heart rhythm is current if condition not reported at w1 but YES at w2
        replace hrhythm_w2 = . if hediac6_w2 == -8 // Confirm W1 diagnosis? Don't know
        replace hrhythm_w2 = . if hedias6_w2 == -8 // Still have it in W2? Don't know
        replace hrhythm_w2 = . if hedim01_w2 == -1 // -1 means not applicable which means "refusal" or "DK" on hedia01_w2 (newly report)

    * In Wave 6 - shall we get rid of refreshment sample for this? meaning: assign missing values to all refreshment samples
        count if finstatw6_w6 == 25 // 796 Cohort-6 core members, i.e., refreshment samples added in W6
        generate hrhythm_w6=0 if inWave6==1 & finstatw6_w6 != 25
        replace hrhythm_w6=1 if hedacar_w6==1 & hedasar_w6==1 // people who had it at previous waves, did not dispute it, and still have condition
        replace hrhythm_w6=1 if hedanar_w6==3   // not had before but R has it now
        replace hrhythm_w6=1 if hedimar_w6==1 & finstatw6_w6 != 25    // for non-refreshment samples, Q is about "since we last interviewed you"
        replace hrhythm_w6=. if hedacar_w6 <= -8 // Confirm previous diagnosis? Don't know
        replace hrhythm_w6=. if hedasar_w6 <= -8 // Still have it in W6? Don't know
        replace hrhythm_w6=. if hedimar_w6 <=-8 // newly reported in W6? Refusal or don't know

    * In Wave 11 ** much more simplified - Do you still have it if you reported it before, if new report, and new among refreshment!!
        gen hrhythm_w11=0 if inWave11==1
        replace hrhythm_w11=1 if (hehaveah_w11==1 | hehaveah_w11==2)    // 1 = have heart rhythm but does not take medication
     // 2 = have heart rhythm and take medication
        replace hrhythm_w11=. if hehaveah_w11<-1 // Don't know

// ANGINA
  // In wave 2, 481 obs with feedforward issue. Strategy adopted for angina is similar to hypertension
        egen hedim_angina_w2 = anycount(hedim*_w2), v(2)    // doctor ever told you that you have angina
            // use hedim*_w2 rather than hedia*_w2 to account for R's other answer that is codeable to angina (8 more obs in this case)
        tab hediac2_w2 hedim_angina_w2
        count if hediac2_w2 == 1 // 298 R confirmed W1 diagnosis of angina
        count if hediac2_w2 == 1 & hedias2_w2 == 1 // Among these 298 R, 211 R still have it in W2
        di 211/298 // 71%
        * Based on data w1-w2, we know that 71% R who confirmed W1 diagnosis of angina still have it in W2
        * We could randomly select 71% among those who reported having it in W1 but have feedforward issue in W2
        set seed 24284
        generate random_angina = runiform() if hediac2_w2==-3
```

```
        gen angina_w2=0 if inWave2==1
        replace angina_w2=1 if hediac2_w2==1 & hedias2_w2==1 // people who had it at w1, did not dispute it, and still have condition
        replace angina_w2=1 if hediac2_w2!=1 & hedian2_w2==3 // not had it at w1 but have it now
        replace angina_w2=1 if hediac2_w2==-1 & hedim_angina_w2==1 // We assume their angina is current if condition not reported at w1 but YES at w2
        replace angina_w2=1 if hediac2_w2==-3 & random_angina<0.71 // this code did not necessarily turn 71%x481 R with value -3 on hediac2_w2 to "having angina" in W2 because some R may have already got value 1 on angina_w2
        replace angina_w2 = . if hediac2_w2 == -8 // Confirm W1 diagnosis? Don't know
        replace angina_w2 = . if hedias2_w2 == -8 // Still have it in W2? Don't know
        replace angina_w2 = . if hedim01_w2 == -1 // -1 means not applicable which means "refusal" or "DK" on hedia01_w2 (newly report)

    * In Wave 6
        count if finstatw6_w6 == 25 // 796 Cohort-6 core members, i.e., refreshment samples added in W6
        generate angina_w6=0 if inWave6==1 & finstatw6_w6 != 25
        replace angina_w6=1 if hedacan_w6==1 & hedasan_w6==1 // people who had it at previous waves, did not dispute it, and still have condition
        replace angina_w6=1 if hedanan_w6==3  // not had before but R has it now
        replace angina_w6=1 if hediman_w6==1 & finstatw6_w6 != 25    // for non-refreshment samples, Q is about "since we last interviewed you"
        replace angina_w6=. if hedacan_w6 == -8 // Confirm previous diagnosis? Don't know
        replace angina_w6=. if hedasan_w6 == -8 // Still have it in W6? Don't know
        replace angina_w6=. if hediman_w6 <=-8 // newly reported in W6? Refusal or don't know

    * In Wave 11 ** Do you still have it if you reported it before, if new report, and new among refreshment!!
        gen angina_w11=0 if inWave11==1
        replace angina_w11=1 if (hehavean_w11==1 | hehavean_w11==2)     // 1 = have angina but does not take medication
     // 2 = have angina and take medication
        replace angina_w11=. if hehavean_w11<-1 // Don't know
     ci mean angina_w*

//
 **** OTHER HEART DISEASES
foreach i in 2 6 11 {
         gen heartdisease_w`i' = 0 if inWave`i' == 1
         replace heartdisease_w`i' = 1 if hfailure_w`i' == 1 | hmurmur_w`i' == 1 | hattack_w`i' == 1 | hrhythm_w`i' == 1 ///
                                       | angina_w`i' == 1
        replace heartdisease_w`i' = . if hfailure_w`i' == . & hmurmur_w`i' == . & hattack_w`i' == . & hrhythm_w`i' == . & ///
                                       angina_w`i' == .
        }
    ci means heartdisease_w*

* ##############################################################
* --------------------------------------------------------------
*
// Panel C. Mental health and cognition

** PSYCHIATRIC PROBLEMS
   * In Wave 2
    egen hedib_psychiatric_w2 = anycount(hedib01_w2 hedib02_w2 hedib03_w2 hedib04_w2), v(7)
        gen psychiatric_w2=0 if inWave2==1
        replace psychiatric_w2=1 if hediad7_w2 ==1 & hedids7_w2==1 // people who had it at w1, did not dispute it, and still have condition
        replace psychiatric_w2=1 if hediad7_w2!=1 & hediam7_w2==3 // not had it at w1 but have it now
        replace psychiatric_w2=1 if hediad7_w2==-1 & hedib_psychiatric_w2==1 // We assume their psychiatric is current if condition not reported at w1 but YES at w2
```

```
        replace psychiatric_w2=1 if heyrc_w2 == 1
        replace psychiatric_w2 = . if hediad7_w2 <= -8 // Confirm W1 diagnosis? Don't know
        replace psychiatric_w2 = . if hedids7_w2 <= -8 // Still have it in W2? Don't know
        replace psychiatric_w2 = . if hedib01_w2 <= -8 //  "refusal" or "DK" on hedib01_w2 (newly report)

    * In Wave 6
        count if finstatw6_w6 == 25 // 796 Cohort-6 core members, i.e., refreshment samples added in W6
        generate psychiatric_w6=0 if inWave6==1 & finstatw6_w6 != 25
        replace psychiatric_w6=1 if hedbdps_w6==1 & heyrc_w6==1 // people who had it at previous waves, did not dispute it, and had it in the past 2 years (There is NO question "do you still have psychiatric problems?" in W6). Our approach is the same as what we did for W11 heart attack and W11 stroke: had it in the past 2 years ~~ still have it
        replace psychiatric_w6=1 if hedbmps_w6==3                // not had before but R has it now
        replace psychiatric_w6=1 if hedibps_w6==1 & finstatw6_w6 != 25    // for non-refreshment samples, Q is about "since we last interviewed you"
        replace psychiatric_w6=. if hedbdps_w6 <= -8 // Confirm previous diagnosis? Don't know
        replace psychiatric_w6=. if heyrc_w6 <= -8 // Still have it in W6? Don't know
        replace psychiatric_w6=. if hedibps_w6 <=-8 // newly reported in W6? Refusal or don't know

    * In Wave 11 ** much more simplified - Do you still have it if you reported it before, if new report, and new among refreshment!!
        gen psychiatric_w11=0 if inWave11==1
        replace psychiatric_w11=1 if (hehaveps_w11==1 | hehaveps_w11==2)    // 1 = have psychiatric but does not take medication
                                                                            // 2 = have psychiatric and take medication
        replace psychiatric_w11=. if hehaveps_w11<-1 // Don't know

    ci means psychiatric_w*

// Depression (CESD-8)
        recode pscedd* pscedf* (2=1) (1=0) (-9/-1=.)
        recode psceda* pscedb* pscedc* pscede* pscedg* pscedh* (2=0) (-9/-1=.)
        foreach t in 2 6 11 {
            egen depression_w`t'=rowtotal(psced*_w`t'), missing
            generate depressed_w`t'=(depression_w`t'>=4) if depression_w`t'!=.
        }
    ci means depressed_w*

// QUALITY OF LIFE
* Note - Quality of Life measured with CASP is asked in the Self-completion Questionnaire
* Quality of life (CASP-19; range 0-57; higher==higher quality of life)
        recode scqol* (-9/-1=.)
        recode scqola* scqolb* scqold* scqolf* scqolh* scqoli* (4=3) (3=2) (2=1) (1=0)
        recode scqolc* scqole* scqolg* scqolj* scqolk* scqoll* scqolm* scqoln* scqolo* ///
        scqolp* scqolq* scqolr* scqols* (1=3) (3=1) (4=0)
        foreach t in 2 6 11 {
            egen casp19_w`t'=rowtotal(scqol*_w`t'), missing
        }
    ci means casp19_w*

// MEMORY
        recode cflisen* cflisd* ( -9/-1=.)
        foreach j in 2 6 11 {
            generate memory_w`j'= cflisen_w`j' + cflisd_w`j'
        }
    ci means memory_w*

* ##############################################################
* ==============================================================
*
```

```
// NON-BLOOD BIOMARKERS from nurse visit

 ** Hypertension biomarker.
    * W2
        gen hypertension_bio_w2 = 0
        replace hypertension_bio_w2 = . if nurse_weight_w2 == .
        replace hypertension_bio_w2 = . if sysval_w2 == -1 & diaval_w2 == -1 // in W2, no Q "do you take medication for blood pressure today?"
        replace hypertension_bio_w2 = 1 if inrange(sysval_w2,140,300) | inrange(diaval_w2,90,150)
        label variable hypertension_bio_w2 "hypertension biomarker"

     * W6 Nurses were instructed to give this advice based on the higher of the last two blood pressure readings – the first reading can be high, as people are sometimes nervous about having their blood pressure taken. (ELSA Nurse Visit user guide, p.13, section 4.1)
         gen hypertension_bio_w6 = 0
         replace hypertension_bio_w6 = . if nurse_weight_w6 == .
         replace hypertension_bio_w6 = . if sysval_w6 == -1 & diaval_w6 == -1 // there is a question about taking medication for hypertension, but to be consistent with w2 and w11, we disregard it.
         replace hypertension_bio_w6 = 1 if inrange(sysval_w6,140,300) | inrange(diaval_w6,90,150)
         label variable hypertension_bio_w6 "hypertension biomarker"

    * W11
        gen hypertension_bio_w11 = 0
        replace hypertension_bio_w11 = . if nurse_weight_w11 == .
        replace hypertension_bio_w11 = . if sysval_w11 == -2 & diaval_w11 == -2 // no Q "do you take medication for blood pressure today?"
        replace hypertension_bio_w11 = 1 if inrange(sysval_w11,140,300) | inrange(diaval_w11,90,150)
        label variable hypertension_bio_w11 "hypertension biomarker"

    ci means hypertension_bio_w*

// Weight/BMI
    ** The maximum weight capacity of the scales was 130kg (20½ stone). If the nurse thought the respondent exceeded this limit then they were instructed to code "Weight not attempted" and ask the respondent for an estimate instead. (ELSA Nurse Visit user guide, p.14, section 4.3) R can give an estimated weight and it is considered valid and their BMI can be calculated.
         recode  bmiobe_w2 (-1=.) (1/3 = 0 "no") (4/6=1 "yes"), gen(obesity_bio_w2)
         label variable obesity_bio_w2 "obesity"

         recode  bmiobe_w6 (-1=.) (1/3 = 0 "no") (4=1 "yes"), gen(obesity_bio_w6)
         label variable obesity_bio_w6 "obesity"

         recode  bmiobe_w11 (-9 -1=.) (1/3 = 0 "no") (4=1 "yes"), gen(obesity_bio_w11)
         label variable obesity_bio_w11 "obesity"
    ci means obesity_bio_w*

** BLOOD BIOMARKERS from nurse visit
     * -11:blood sample not taken; -8: cannot measure parameter reliably as TRIG value > 13mmol/L; -7:sample unusable for other reason; -6: sample took more than 5 days to reach the lab; -3: sample received but was haemolysed so not suitable for analysis; -2: sample received but insufficient blood for analysis; -1: sample not received;

// Diabetes biomarker   >= 6.5% or >=48 mmol/mol
     recode hba1c_w2 (-11/-1 =.)  (3/6.4 = 0 "No") (6.5/100 = 1 "Yes"), gen(diabetes_bio_w2)
     label variable diabetes_bio_w2 "diabetes biomarker"
     recode hba1c_w6 (-11/-1 =.)  (3/47 = 0 "No") (48/130 = 1 "Yes"), gen(diabetes_bio_w6)
     label variable diabetes_bio_w6 "diabetes biomarker"
     recode hba1c_w11 (-11/-1 =.)  (3/47 = 0 "No") (48/130 = 1 "Yes"), gen(diabetes_bio_w11)
     label variable diabetes_bio_w11 "diabetes biomarker"
     *  -11:blood sample not taken; -9:sample leaked -8: cannot measure parameter reliably as TRIG value > 13mmol/L; -7:sample unusable for other reason; -6: sample took more than 5 days to reach the lab; -2: sample received but insufficient blood for analysis; -1:sample not received;
    ci means diabetes_bio*
```

```

// High total cholesterol. if taking medication for high cholesterol not asked in w2
   * Cut-off point 5 mmol/L. REF: https://www.nhs.uk/conditions/high-cholesterol/cholesterol-levels/
   * in W2 there is no question "do you take statin to lower your cholesterol levels?"
    foreach j in 2 6 11 {
        recode chol_w`j' (-11/-1 = .)  (0.1/4.9 = 0 "no") (5/20 = 1 "yes"), gen(cholesterol_bio_w`j')
        label variable cholesterol_bio_w`j' "high cholesterol biomarker"
        }
    ci means cholesterol_bio*

// CRP  cutoff point is 3 mg/L
    recode hscrp_w* (-11/-1 = .)
    foreach j in 2 6 11 {
        recode hscrp_w`j' (-11/-1 = .) (0.00000001/2.999999999 = 0) (3/999 = 1 "high-risk CRP cutoff 3"), gen(crp_w`j')
        label variable crp_w`j' "high-risk CRP"
        }
   ci means hscrp_w*


save "C:\Users\Data\ELSA Health trends\all_2611.dta", replace
* use  "C:\Users\Data\ELSA Health trends\all_2611.dta", clear

keep idauniq gor_w* sc_weight_w* x_weight_w* blood_weight_w* nurse_weight_w* inWave* ELSA_inwave2_6_11 ///
     age_group_4cat_w* female_w* wealth_q_w* education_w* ///
     srh_w* gali_w* longill_w*  pain_w* severe_pain_w*  ///
     adl_w* adl_bin_w* iadl_w* iadl_bin_w* mobi_if_w* mobi_number_w* ///
     hypertension_w* angina_w* hfailure_w* hattack_w* hmurmur_w* hrhythm_w* cholesterol_w* diabetes_w* ///
     lung_w* stroke_w* heartdisease_w* ///
     psychiatric_w*  depression_w* depressed_w* casp19_w* ///
     hypertension_bio_w2 hypertension_bio_w6 hypertension_bio_w11 obesity_bio_w2 obesity_bio_w6 obesity_bio_w11 ///
     diabetes_bio_w2 diabetes_bio_w6 diabetes_bio_w11 cholesterol_bio_w2 cholesterol_bio_w6 cholesterol_bio_w11 ///
     crp_w2 crp_w6 crp_w11  ///
     memory_w* ///
     refreshtype_w2 refreshtype_w6 refreshtype_w11 finstat_w6
     // This is a wide format dataset (where multiple observations for the same R are in the same row) for the variables of interest
    ** for simplicity, it can be saved
    save "C:\Users\Data\ELSA Health trends\slim_wide_2611.dta", replace
    rename (gor_w2_w2 gor_w6_w6 gor_w11_w11) (gor_w2 gor_w6 gor_w11)
// we now create a dataset in the "LONG" format
    reshape long gor_w sc_weight_w x_weight_w blood_weight_w nurse_weight_w inWave ///
     age_group_4cat_w female_w wealth_q_w education_w ///
     srh_w gali_w longill_w  pain_w severe_pain_w  ///
     adl_w adl_bin_w iadl_w iadl_bin_w mobi_if_w mobi_number_w ///
     hypertension_w angina_w hfailure_w hattack_w hmurmur_w hrhythm_w cholesterol_w diabetes_w ///
     lung_w stroke_w heartdisease_w ///
     psychiatric_w depression_w depressed_w casp19_w ///
     hypertension_bio_w obesity_bio_w ///
     diabetes_bio_w cholesterol_bio_w ///
     crp_w ///
     memory_w refreshtype_w, ///
     i(idauniq) j(wave)

     drop if inWave==.
     count // 23,028

    * gor is coded differently, we create a new variable
     gen region = .
```

```
        replace region = 1 if gor_w == "A" | gor_w == "E12000001"
        replace region = 2 if gor_w == "B" | gor_w == "E12000002"
        replace region = 3 if gor_w == "D" | gor_w == "E12000003"
        replace region = 4 if gor_w == "E" | gor_w == "E12000004"
        replace region = 5 if gor_w == "F" | gor_w == "E12000005"
        replace region = 6 if gor_w == "G" | gor_w == "E12000006"
        replace region = 7 if gor_w == "H" | gor_w == "E12000007"
        replace region = 8 if gor_w == "J" | gor_w == "E12000008"
        replace region = 9 if gor_w == "K" | gor_w == "E12000009"
        label define region 1 "North East" 2 "North West" 3 "Yorkshire & The Humber" 4 "East Midlands" 5 "West Midlands" ///
                    6 "East of England" 7 "London" 8 "South East" 9 "South West", replace
        label values region region

    ** for simplicity, we also save this long dataset
    save "C:\Users\Data\ELSA Health trends\slim_long_2611.dat", replace

* !!!! Sample size is different for different outcomes
    tabstat x_weight_w, statistics(mean count) by(wave) // to see sample size for main survey
    tabstat sc_weight_w, statistics(mean count) by(wave) // to see sample size for self-completion
    tabstat nurse_weight_w, statistics(mean count) by(wave) // to see sample size for nurse visit
    recode blood_weight_w (-1=.)
    tabstat blood_weight_w, statistics(mean count) by(wave) // to see sample size for venous blood sample assay


// Stata's dtable command only gives SD for continuous variables, but we want to show 95% CI
// For categorical variables, we present freq + %, so command dtable is applicable
// MAIN SURVEY sampling weight
    svyset [pweight=x_weight_w], psu(idauniq) strata(region) vce(robust)
    * categorical variables
    foreach k in 2 6 11 {
        foreach var in i.age_group_4cat_w i.wealth_q_w i.education_w {
            svy: proportion `var' if wave==`k'
            }
            }
    * binary variables
    foreach variable in female_w srh_w  longill_w  pain_w severe_pain_w   adl_bin_w  iadl_bin_w mobi_if_w hypertension_w ///
                        cholesterol_w diabetes_w stroke_w heartdisease_w lung_w psychiatric_w depressed_w depression_w memory_w {
        foreach k in 2 6 11 {
            svy: mean `variable' if wave ==`k'
                }
            }
    * Note -- We re-estimate chronic conditions for w6 making sure we exclude refreshment sample
    foreach variable in hypertension_w cholesterol_w diabetes_w stroke_w heartdisease_w lung_w psychiatric_w {
            svy, subpop(if finstat_w6 != 25): mean `variable' if wave == 6
        }

// NURSE sampling weight
    svyset [pweight=nurse_weight_w], psu(idauniq) strata(region) vce(robust)
    **
        foreach variable in hypertension_bio_w obesity_bio_w  {
        foreach g in 2 6 11 {
            svy: mean `variable' if wave == `g'
            }
        }

// VENOUS BLOOD sampling weight
    svyset [pweight=blood_weight_w], psu(idauniq) strata(region) vce(robust)
    foreach variable in cholesterol_bio_w diabetes_bio_w crp_w {
```

```
        foreach g in 2 6 11 {
        svy: mean `variable' if wave == `g'
        }
    }

// CASP-19 using sc_weight for Table 1
    svyset [pweight=sc_weight_w], psu(idauniq) strata(region) vce(robust)
        foreach g in 2 6 11 {
        svy: mean casp19_w if wave == `g'
        }

********************************************************************************
```
